# Healthy diet metrics for children and adolescents and their suitability for global monitoring: a critical review

**DOI:** 10.1101/2025.07.29.25332348

**Authors:** Alissa M. Pries, Hope Craig, Vrinda Mehra, Edward A. Frongillo, Giles T. Hanley-Cook, Chika Hayashi, Kuntal Saha, Isabela Fleury Sattamini, Teresa R. Schwendler, Jennifer C. Coates, the Healthy Diets Monitoring Initiative

## Abstract

**Background:** Healthy diets during childhood and adolescence are paramount for growth, development, and long-term health. However, there is a lack of low-burden standardized metrics to assess and monitor healthy diets among children and adolescents aged 2-19 years of age on a global scale.

**Objective:** This critical review aimed to identify and evaluate existing metrics for assessing healthy diets in this age group and to determine their suitability for global monitoring based on feasibility and adaptability across different contexts.

**Methods:** A systematic search was conducted across three global databases, encompassing both peer-reviewed and grey literature.

**Results:** A total of 127 distinct healthy diet metrics were identified many of which were developed or adapted based on national dietary guidelines across various geographical contexts. Only five were deemed suitable for global monitoring due to their feasibility and adaptability: the Individual Dietary Diversity Score, 7 food group Minimum Dietary Diversity, 10 food group Minimum Dietary Diversity, Healthy Plate Variety Score, and Adapted ultra-processed food (UPF) Nova Score. Among these metrics, diversity was the most commonly measured sub-construct of a healthy diet, while only the Adapted Nova Score aimed to capture moderation. These five metrics were further evaluated for construct validity, reliability, and cross-context equivalence, which revealed large evidence gaps, particularly regarding sensitivity to change and test-retest reliability.

**Conclusions:** These findings highlight the need for additional research to validate healthy diet metrics globally to ensure their accuracy, sensitivity, and reliability to differentiate populations and changes over time. Developing robust, low-burden metrics is essential for informing effective, timely nutrition policies and interventions aimed at improving the diets of children and adolescents worldwide.

**Statement of significance:** This review is the first to systematically evaluate the feasibility and adaptability of existing healthy diet metrics for global monitoring among children and adolescents, identifying key gaps in their validity and reliability, particularly regarding sensitivity to change and test-retest reliability.

## Introduction

Healthy diets during childhood and adolescence are essential for growth, development and overall well-being (1,2). Optimal nutrition during these periods help to ensure children and adolescents receive adequate nutrients without excess, to prevent both undernutrition and obesity, support cognitive development, and boost immunity, to reduce the risks of infection and disease. Establishing healthy eating habits in early life sets the foundation for health in adulthood by lowering the risk of developing diet-related noncommunicable diseases, such as diabetes and heart disease, and contributing to better health outcomes throughout life (2). Globally, countries increasingly face the triple burden of malnutrition - the coexistence of overweight and obesity, undernutrition, and micronutrient deficiencies, which stem from rapidly changing food environments, unsustainable agrifood systems and evolving dietary choices (3,4). Over the last three decades, the combined prevalence of thinness and obesity among boys and girls has increased in over 130 countries (5).

Healthy diet metrics are essential for the standardized assessment and monitoring of dietary patterns among populations across settings and over time (6). The use of healthy diet metrics allows for the design of evidence-informed nutrition policies and interventions to improve the healthiness of diets and address nutritional gaps. Previous reviews by Marshall et al. (7) in 2014 and the subsequent update by Dalwood et al. (8) in 2020 have catalogued diet quality indices developed for use among children and adolescents. The most recent of these identified 128 unique indices and assessed their content, validity, and associations with health outcomes. These reviews provided an important foundation for understanding the landscape of diet quality metrics used among children and adolescents.

Nevertheless, significant gaps exist in our understanding of which existing measures and indicators are suitable for assessing diets among children and adolescents at scale and at regular intervals, particularly for global monitoring purposes (8). There is a need to i) understand the spectrum of existing metrics; ii) which sub-constructs of a healthy diet they aim to measure; and iii) to evaluate healthy diet metrics’ applicability across diverse contexts and age groups. In addition to feasibility and adaptability, the accuracy, reliability, and cross-context equivalence of healthy diet metrics are crucial for ensuring that they can be validly applied across countries. Current knowledge is limited, however, regarding the extent to which healthy diet metrics for children and adolescents perform in varied cultural and socioeconomic contexts, where dietary patterns and comprehension of questionnaires can differ substantially. (9)

In 2022, the Food and Agriculture Organization of the United Nations (FAO), the United Nations Children’s Fund (UNICEF), and World Health Organization (WHO) initiated the Healthy Diets Monitoring Initiative (HDMI), an effort to strengthen the knowledge base related to healthy diets metrics and their lower-burden data collection methods and to promote their uptake for high-frequency global and national monitoring. The HDMI mission statement is to “enable national and global decision-makers and stakeholders to monitor and achieve healthy diets for people and the planet” (10). HDMI commissioned a background paper, reviewing and comparing seven healthy diet metrics against a set of predefined assessment criteria (11). The report, however, does not compile or assess metrics’ detailed validation findings due to time constraints and only briefly touches on the question of cross-context equivalence. Furthermore, the seven metrics reviewed in this report were applicable to adult populations, and did not include metrics tailored to the specific nutritional requirements and dietary behaviours of children or adolescents. An important next step in evaluating the evidence base is to identify metrics that were designed to capture the various sub-constructs of a healthy diet for children and adolescents and assess their suitability for global monitoring.

The objectives of this review were to: i) identify the existing metrics that have been developed or adapted to assess the sub-constructs of a healthy diet among children and adolescents aged 2-19 years; ii) assess which of these healthy diet metrics are suitable for global monitoring based on their feasibility and adaptability; and iii) summarize the accuracy, test-retest reliability, sensitivity to change, and cross-context equivalence of healthy diet metrics suitable for global monitoring.

## Methods

### Review design

A systematic search and critical review involving a multi-stage method was used. In the first stage, the systematic search was conducted to identify all existing healthy diet metrics and methods used among children and adolescents. Thereafter, a critical review of the identified healthy diet metrics and methods was conducted to assess their suitability for use in global monitoring based on a set of pre-determined criteria. Of this subset of healthy diet metrics, literature pertaining only to these metrics was reviewed to summarize evidence on the accuracy, reliability, sensitivity and cross-context equivalence of metrics and methods suitable for global monitoring. Accuracy included evidence on the construct validity of metrics, reliability included studies assessing the test-retest reliability of metrics, sensitivity to change over time included studies simultaneously comparing changes in a healthy diet metric and a reference metric of dietary intake over time, and cross-context equivalence refers to whether a metric performed equivalently across various contexts, including geography and populations of varying socioeconomic levels (12). This review covered children and adolescents aged 2-19 years, with consideration of metrics developed or adapted across age subgroups: 2-4 years, 5- 9 years, 10-14 years and 15-19 years.

### Search strategy

The systematic search built upon a systematic review conducted by Dalwood et al. (2020) (8), which summarized diet quality indices used among children and adolescents and covered literature published up to January 11 2019. For this review, literature identified by Dalwood et al. on metric development or adaptation was included and a search for literature published since January 12 2019 through June 13, 2024 was conducted to identify additional healthy diet metrics. Databases searched include two used by Dalwood et al. (Embase and MEDLINE) and a third database to capture peer-reviewed and grey literature (Global Health). Search terms were adapted from those used by Dalwood et al. and covered two broad categories: i) the target population (children and adolescents aged 2-19 years) and ii) healthy diet metrics (**Table 1**). Healthy diet metrics were defined as metrics based on the intake of nutrients, foods, or food groups, or combinations, with the aim to measure one or more sub-constructs of a healthy diet. Sub-constructs of a healthy diet included “nutrient adequacy,” “nutrient density,” “macronutrient balance,” “diversity,” “moderation,” “favourable dietary pattern,” and “food safety” (11,13); further sub-constructs of a healthy diet for children and adolescents measured in the identified metrics were also captured. Sub-constructs were extracted whether stated explicitly by the study’s authors or identified implicitly by the full-text reviewers. Operational definitions of these sub-constructs can be found in **Supplemental Table 1**.

**Table 1.** Systematic search terms.

|  |  |
| --- | --- |
| 1 | (child* or adolescent* or teen* or youth* or kid* or pre-teen* or minor*).tw |
| 2 | exp child/ or exp adolescent/ |
| 3 | 1 OR 2 |
| 4 | ("diet quality" or "dietary quality" or "dietary variety" or "diet metric" or "diet measure*" or "dietary measure*" or DQI or DQI-I or "Healthy Eating Index" or HEI or YHEI or "Recommended Food Score" or RFS or "variety score" or "variety ind*" or "diversity score" or "diversity ind*" or DDI or ACARFS or KIDMED or DGI CA or FVI or E-KINDEX or AMQI or DQQ or "diet quality questionnaire" or GDR or "global dietary recommendation*" or "NCD-protect" or "NCD-risk" or "Nova UPF score" or GDQS or "global diet quality score" or MDD or "minimum dietary diversity").tw |
| 5 | 3 AND 4 |
| 6 | Limit to 2019 – present, English language only |

### Inclusion and exclusion criteria

Studies were included if they report the development, adaptation, or validation of a healthy diet metric involving a population of children and adolescents aged 2-19 years (**Table 2**). To ensure that the review focused specifically on metrics developed, adapted or validated for the specific nutritional requirements and dietary behaviours of children and/or adolescents, studies were excluded if they involved a wider age range than 2-19 years and did not disaggregate results for children and adolescents within 2-19 years. All identified literature, both from Dalwood et al. and from the updated systematic search, were subject to the inclusion and exclusion criteria.

**Table 2.** Inclusion and exclusion criteria.

|  | **Inclusion criteria** | **Exclusion criteria** |
| --- | --- | --- |
| Population | Children and adolescents aged 2.0-19.9 years (sample can have a wider age range but must present disaggregated results for an age range within this window) | Samples involving only infants and young children 0-23.9 months of age or adults 20 years or older |
| Language | English | Non-English |
| Scope | Report the development, adaptation or use of a healthy diet metric | Metrics where a substantial component is not related to dietary intake; metrics evaluating intake of a single nutrient, food, beverage, or food group |
| Reference type | Peer-reviewed publications, reports, grey literature (i.e. reports, working papers, white papers) | Abstracts; reviews that did not cover development or adaptation of healthy diet metrics |

### Screening

Covidence – a systematic review management software – was used to identify duplicate references and for the screening process. Following deduplication, titles and abstracts of all citations were screened for eligibility, then full texts of eligible citations were reviewed for final inclusion. Title and abstract screening and full-text review of each paper was conducted by one of two researchers (AP and HC). Double coding of 5% of papers was initially conducted to ensure consistency in screening between the two researchers before moving to single coding. In addition, criteria for inclusion and exclusion were presented in Covidence for each researcher to refer throughout screening. Reference lists of eligible literature were manually searched to identify any further literature for inclusion.

### Data extraction

To facilitate the multi-phased method of this review, during the screening process, included papers were tagged within Covidence to indicate if they contained details on i) metric development, ii) metric adaptation, iii) predictive capacity of a metric, and/or iv) all other forms of validity or accuracy of a metric. There were two phases of data extraction. Data were first extracted from all papers tagged as containing details on metric development or adaptation to identify existing healthy diet metrics for children and adolescents. These metrics were then assessed for suitability for high-frequency global monitoring (details below). In the second phase of data extraction, data were extracted from papers tagged as containing details on predictive capacity or other forms of validity/accuracy of a metric, but only among studies that reported on the predictive capacity, validity, or accuracy of the subset of metrics suitable for global monitoring. For both phases, data were extracted by one researcher (AP).

### Assessment of suitability for global monitoring

All healthy diet metrics for children and adolescents were assessed for suitability for global monitoring based on their feasibility and adaptability (11). The assessment of feasibility considered a metric’s ease of computation and ease of interpretation for the sub-construct measured, while adaptability considered the universality of the evidence underlying a metric and a metric’s application across multiple country contexts (**Table 3**). Metrics rated as medium or high for ease of computation, high for ease of interpretation, and high for adaptability were preliminarily deemed suitable for global monitoring. This subset of suitable metrics was then further evaluated for feasibility of data collection, specifically considering the burden on interviewers and respondents in terms of time, cognitive effort, and invasiveness. Cost is a key component of data collection feasibility, however, cost is highly dependent on the methods used for data collection; therefore, interviewer and respondent burden were considered as proxies for cost, where higher degrees of burden for both are typically associated with higher costs for interviewer training and data collection time. Due to the limited availability of data collection feasibility details in the literature identified through the systematic search, a purposive sample of experts in the field of child and adolescent dietary assessment was contacted to gather additional insights. If a metric within this sub-set was rated as low or medium for both interviewer and respondent burden, it was considered suitable for global monitoring.

**Table 3.** Criteria to assess suitability of healthy diet metrics for global monitoring.

| Criteria | Operationalization |
| --- | --- |
| Feasibility – ease of computation | • High: points assigned per respondent; score or indicator calculated<br>• Medium: food and ingredients assigned to the appropriate metric item; points assigned per respondent; score calculated; indicator tabulated<br>• Low: dietary data processed to estimate quantities consumed of food and ingredients per respondent; food and ingredients assigned to the appropriate metric item; points assigned per respondent; score calculated; indicator tabulated<br>• Very low: food composition data merged; dietary data processed to estimate quantities consumed of food, ingredients, and nutrients per respondent; food, ingredient and nutrient assigned to the appropriate metric item; quantity of consumption per metric item per respondent summed; points assigned per respondent; score calculated; indicator tabulated |
| Feasibility – ease of interpretation of sub-constructs measured | • High: metric captures only one sub-construct or allows for disaggregated sub-metrics for each sub-construct (e.g. a metric has two sub-metrics of ‘unhealthy’ and ‘healthy’ dietary components)<br>• Low: multiple sub-constructs captured within the metric and disaggregated interpretation not possible (e.g. a metric including various unhealthy healthy foods with no disaggregation) |
| Adaptability | • High: For metrics based on dietary recommendations/guidelines, based on regional or international dietary recommendations/guidance (as opposed to national dietary guideline or localized dietary pattern); OR for all metrics, metric developed or adapted for more than one country.<br>• Low: Metric based on national dietary guideline or localized dietary pattern and only developed or adapted for one country. |
| Feasibility – burden of data collection | Interviewer burden: level of expertise required, and perceived physical and cognitive effort<br>• Low: minimal expertise in nutrition/health or minimal training required; minimal physical effort required (e.g. low interview time, no equipment beyond questionnaire); low cognitive effort (e.g. capturing data on food/food groups consumed)<br>• Medium: moderate expertise in nutrition/health or moderate level of training required (e.g. bachelors in topics); moderate physical effort required (e.g. moderate interview time, some additional equipment required); moderate cognitive effort (e.g. portion size estimation required with images)<br>• High: high expertise in nutrition/health or high level of training required (e.g. masters in topics); high physical effort (e.g. high interview time, substantial equipment required); high cognitive effort (e.g. portion size estimation required with food models) |
|  | Respondent burden: degree to which a respondent perceives their participation as difficult, time-consuming or emotionally stressful.<br>• Low: Dietary assessment for metric requires less than 15 minutes; only one assessment is required; interview is minimally invasive<br>• Medium: Dietary assessment for metric requires 15-30 minutes; only 1-2 assessments is required; interview is somewhat invasive<br>• High: Dietary assessment for metric requires 30+ minutes; multiple assessments are required; interview is somewhat invasive |

### Study quality

For studies reporting the validity, reliability, sensitivity to change over time or cross-context equivalence of a metric, critical appraisal was conducted using the Academy of Nutrition and Dietetics Quality Criteria Checklist (QCC) (14). The QCC evaluates risk of bias in participant selection, generalizability, data collection and analysis. Based on this evaluation, a study’s quality was rated as positive, neutral, or negative. In line with the prior review by Dalwood et al. (8), studies were not excluded if their quality was determined to be negative.

## Results

A total of 15,257 papers were identified across the three databases included in the systematic search (**Figure 1**). Following deduplication and title and abstract screening, 1,977 papers were included for full-text review to assess their eligibility, of which a total of 129 were included based on them containing details on development or adaptation of a healthy diet metric for children or adolescents, from which a total of 127 metrics were identified. Fifty of these 127 metrics were additions to those identified by the review by Dalwood et al (8), indicating that over one-third of the healthy diet metrics for children and adolescents aged 2-19 years identified by this present review have been developed or adapted for this population in the past five years. Of these 127 metrics, five were identified as suitable for global monitoring based on their feasibility and adaptability. An additional 15 papers evaluating the validity or reliability of these five metrics were also included in the review.

**Figure 1.**
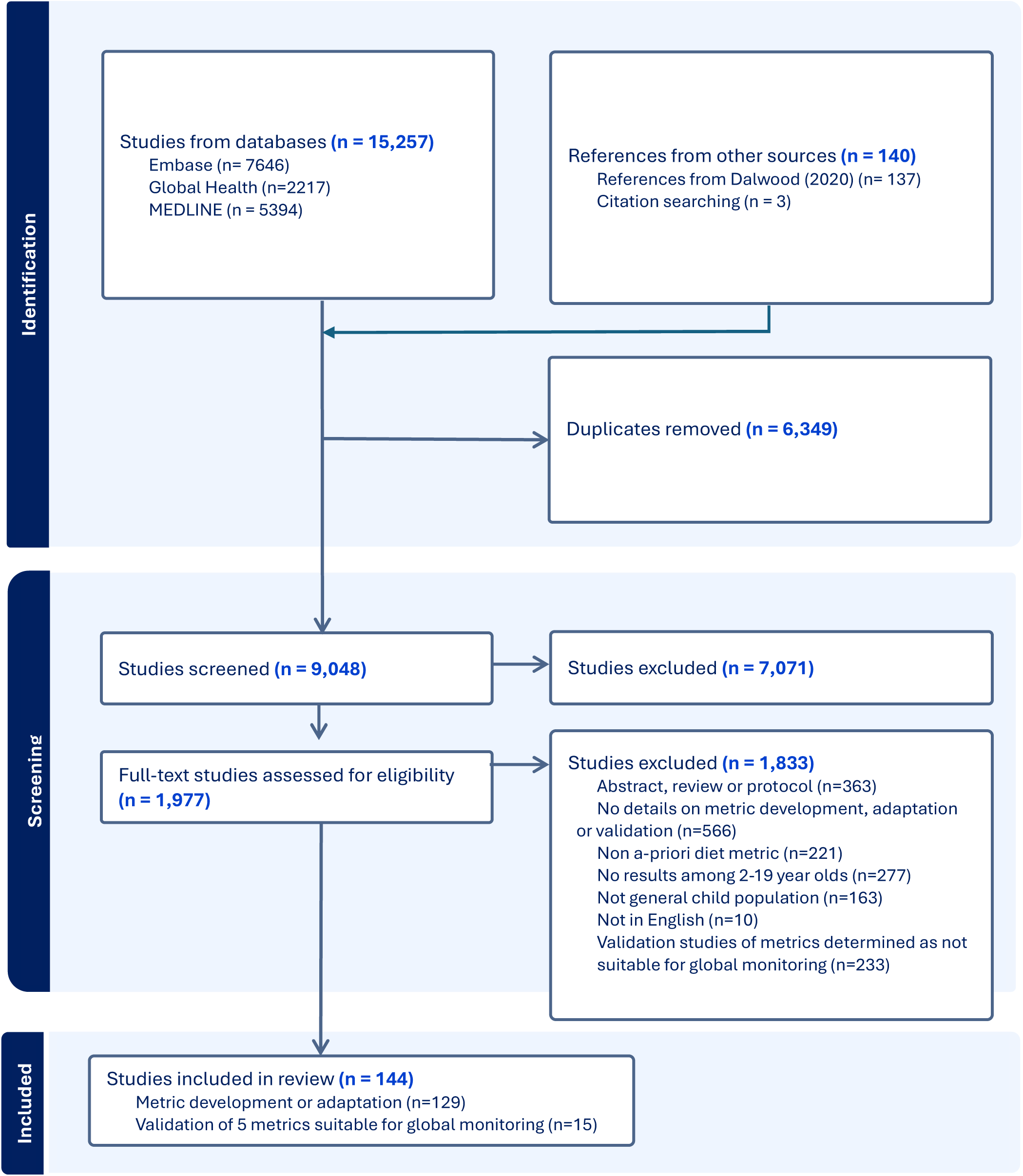
Preferred Reporting Items for Systematic reviews and Meta-Analyses (PRISMA) flow diagram of study selection

### Healthy diet metrics for children and adolescents

Most of the identified metrics (n=103, 81.1%) were based on adherence to national dietary guidelines from a range of countries across different regions, including East Asia, Europe, Latin America, North America, Oceania, South Asia and Southeast Asia (**Table 4**). Additionally, 11 metrics (8.7%) reflected adherence to a national or localized dietary pattern, 12 metrics (9.4%) were based on global dietary guidance, four metrics (3.1%) aligned with regional dietary guidelines, and two metrics (1.6%) considered global evidence on foods associated with health risks or benefits. Most metrics aimed to assess multiple sub-constructs of a healthy diet within a composite score, with diversity (n=119, 93.7%) and moderation (n=105, 82.7%) being the most widely included. Over 80% of metrics combined the sub-constructs of diversity and moderation into a composite measure, representing a favourable dietary pattern. Other sub-constructs of a healthy diet, such as nutrient adequacy (n=10, 7.9%) and macronutrient balance (n=1, 0.8%), were less frequently addressed. Twenty-six metrics (20.5%) measured additional sub-constructs indirectly related to diet, including behaviours that could influence the healthiness of a child or adolescent diets, such as breakfast consumption (n=8, 6.3%) and family meal participation (n=4, 3.1%), as well as lifestyle behaviours such as sedentary activity (n=5, 3.9%) and physical activity (n=4, 3.1%). For most metrics (n=98, 77.2%), the sub-constructs reflected were implicitly identified by the researchers and only 29 metrics explicitly noted purposive measurement of their sub-constructs of a healthy diet.

**Table 4.**
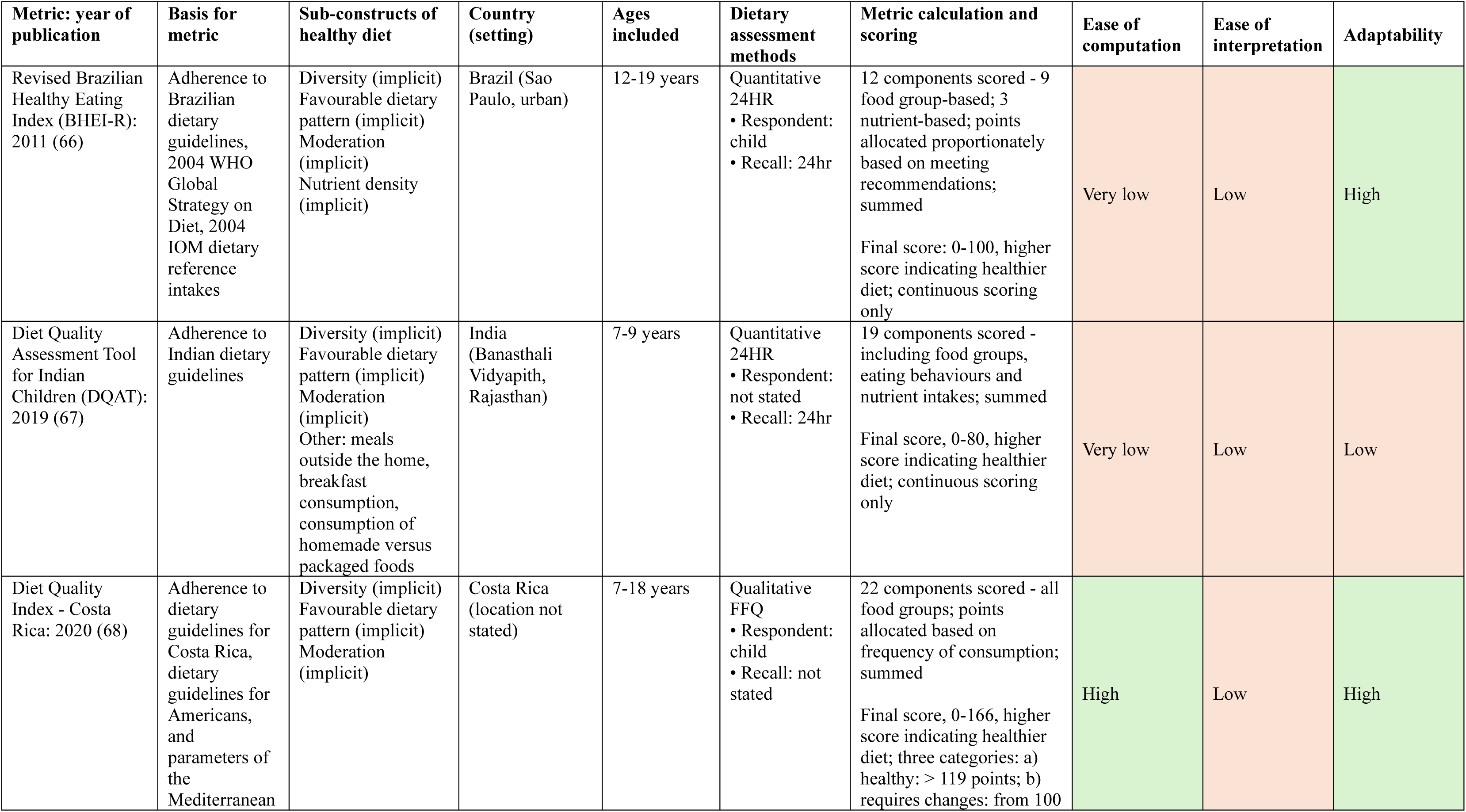
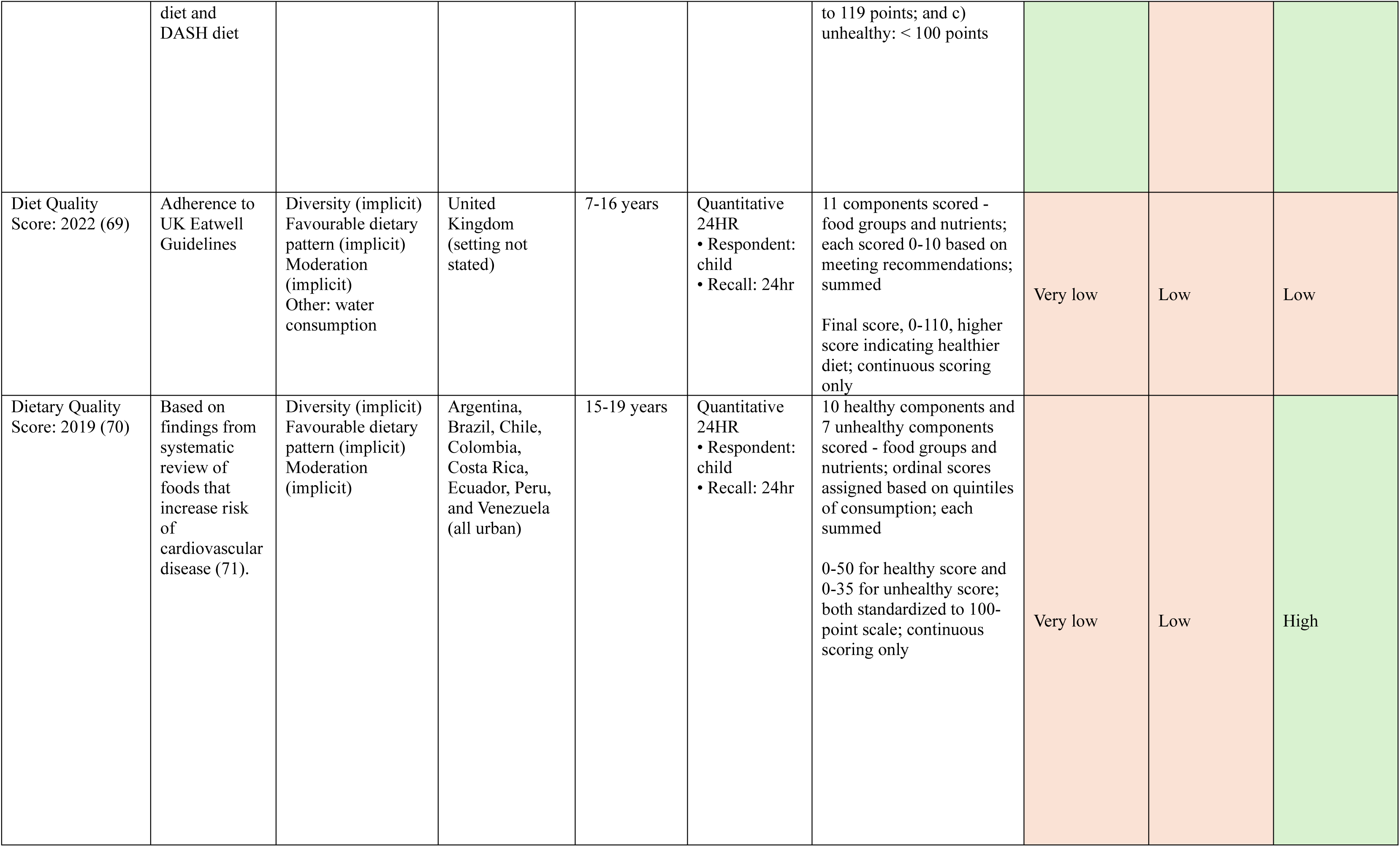
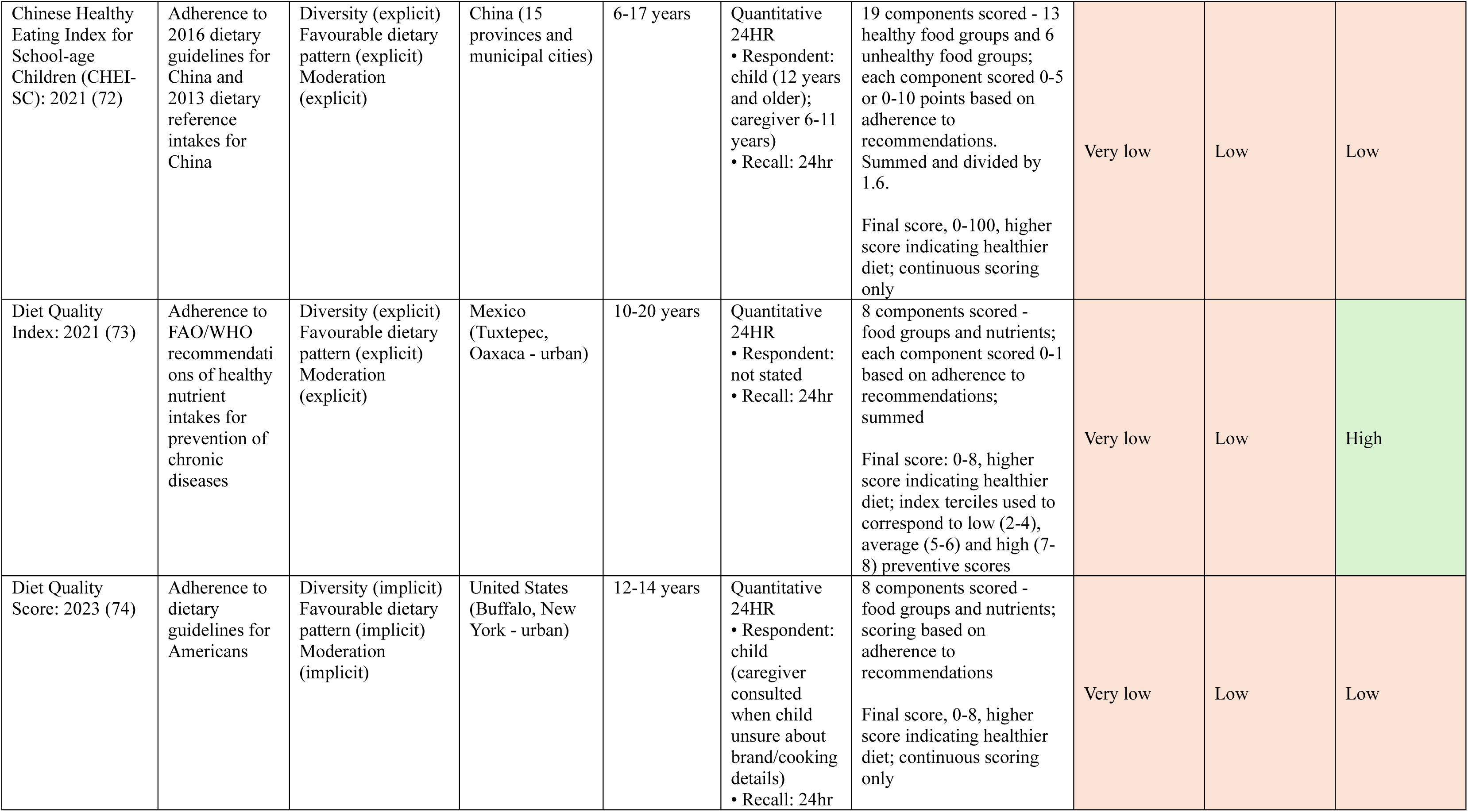
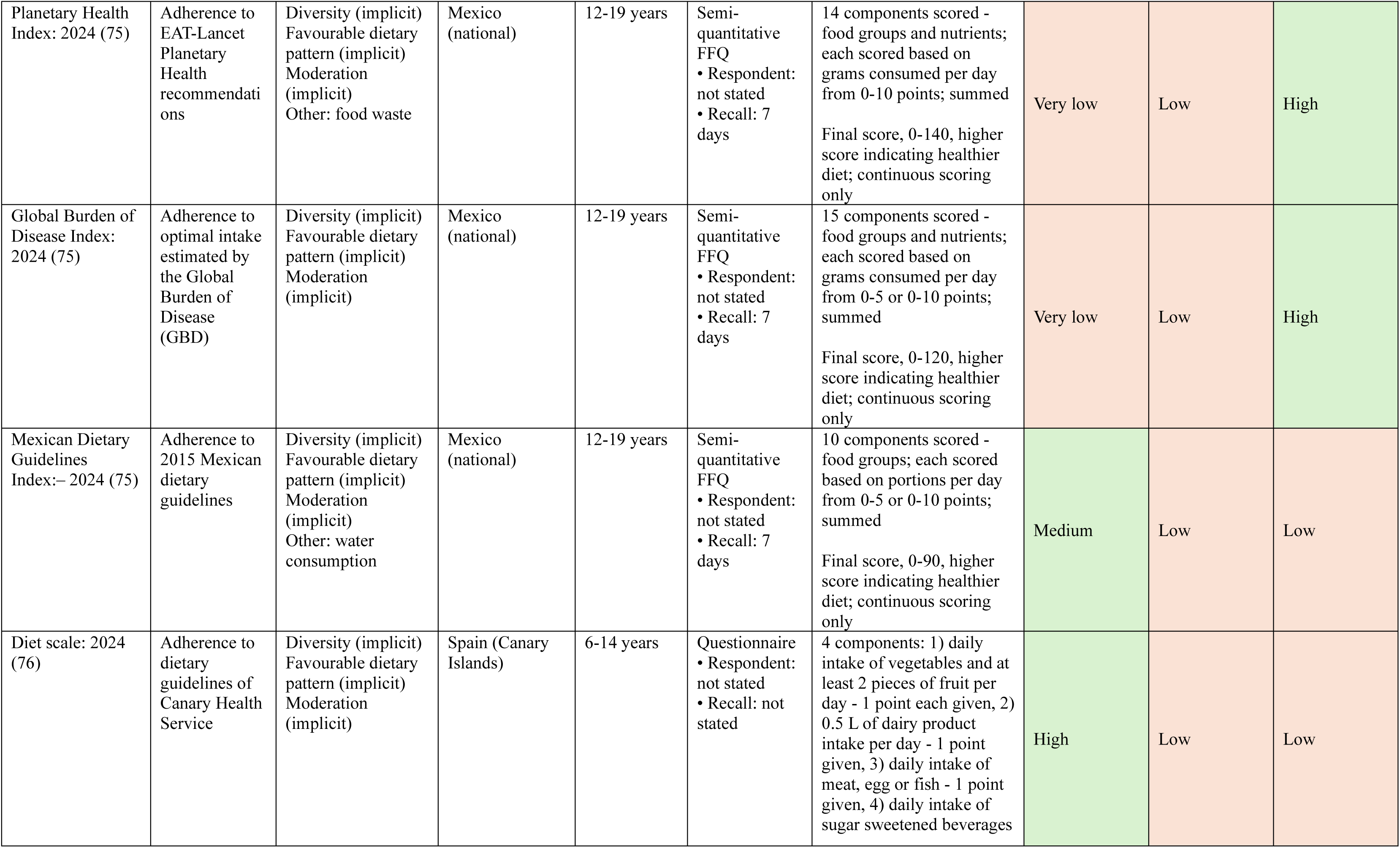
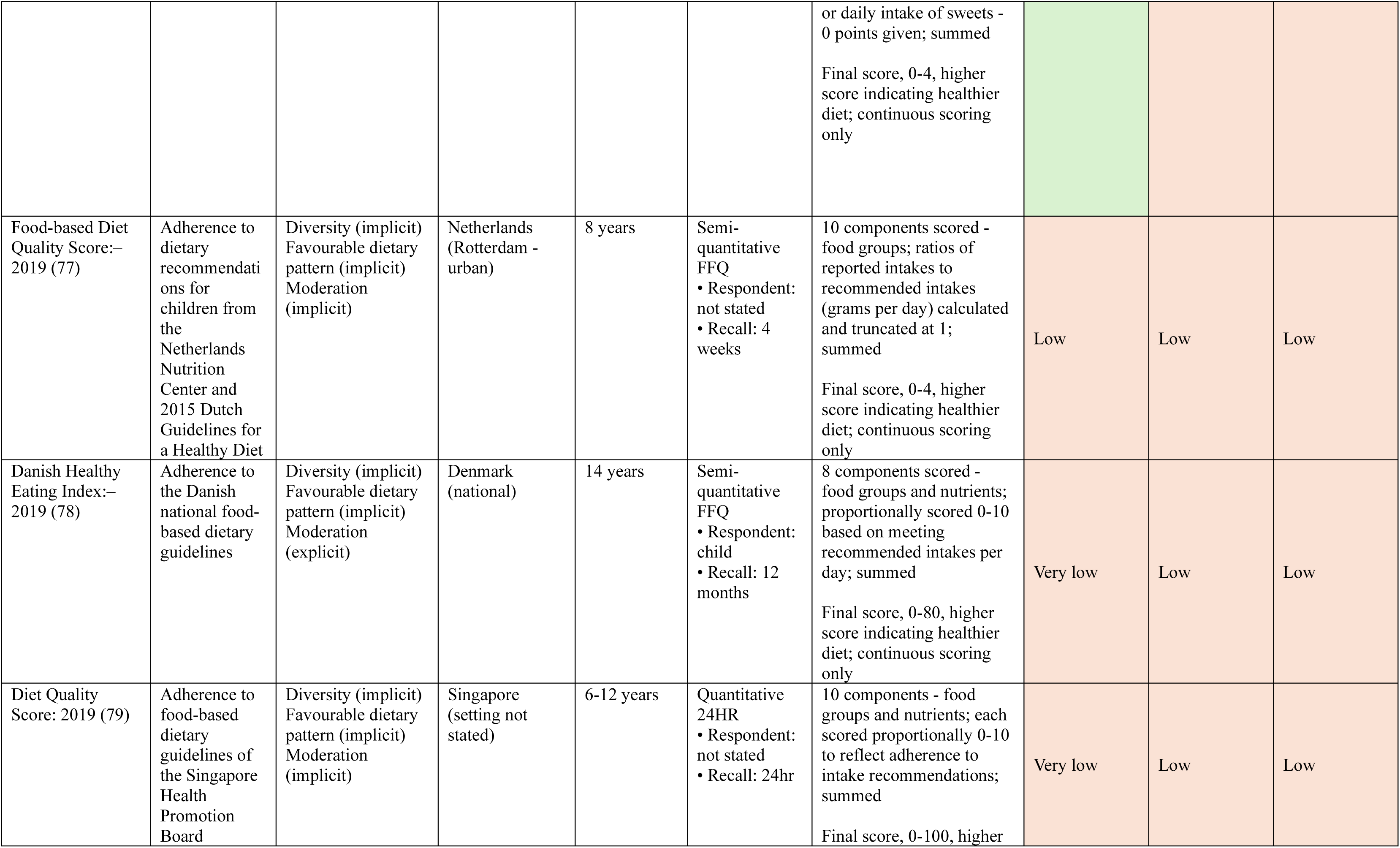
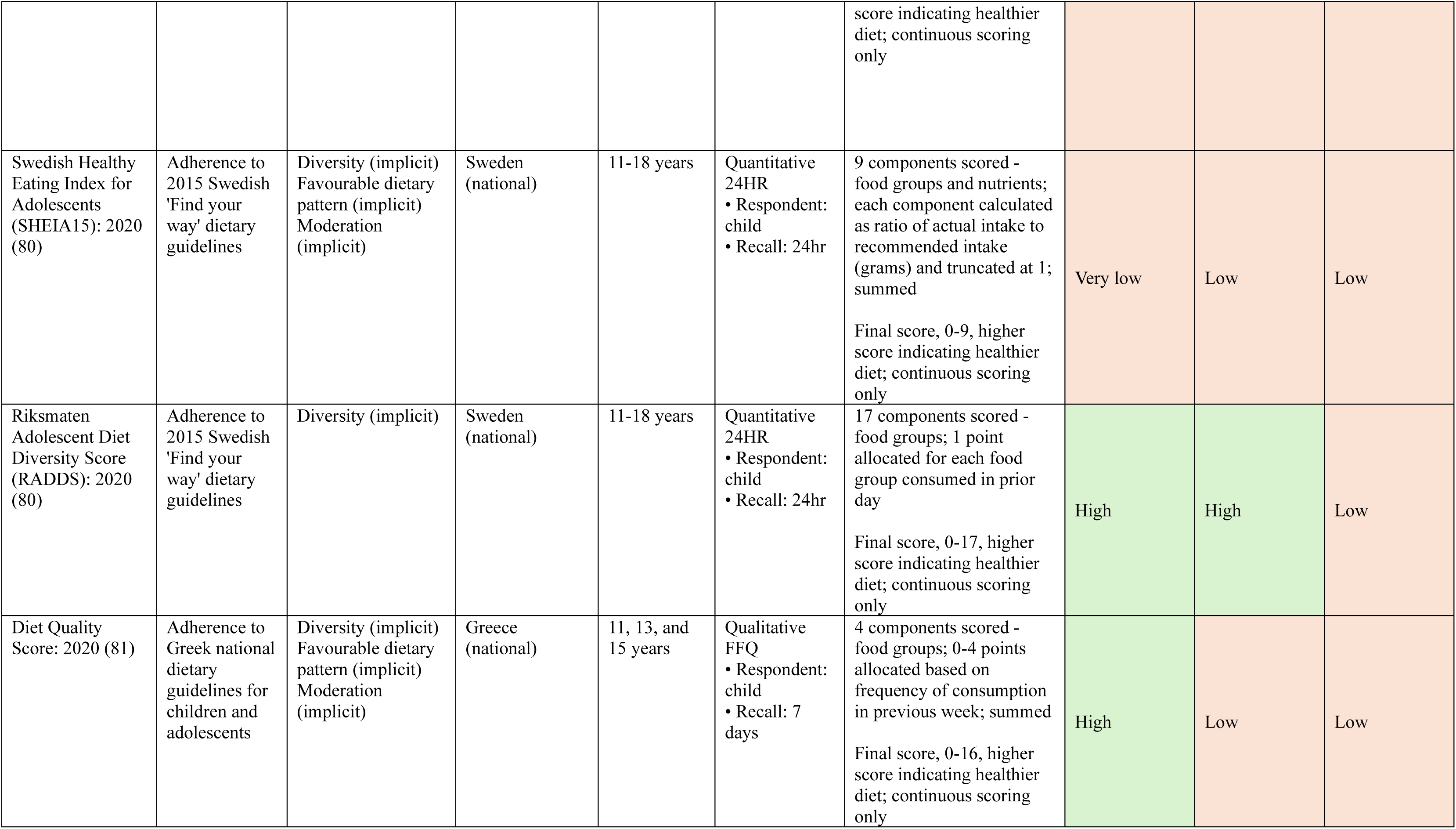
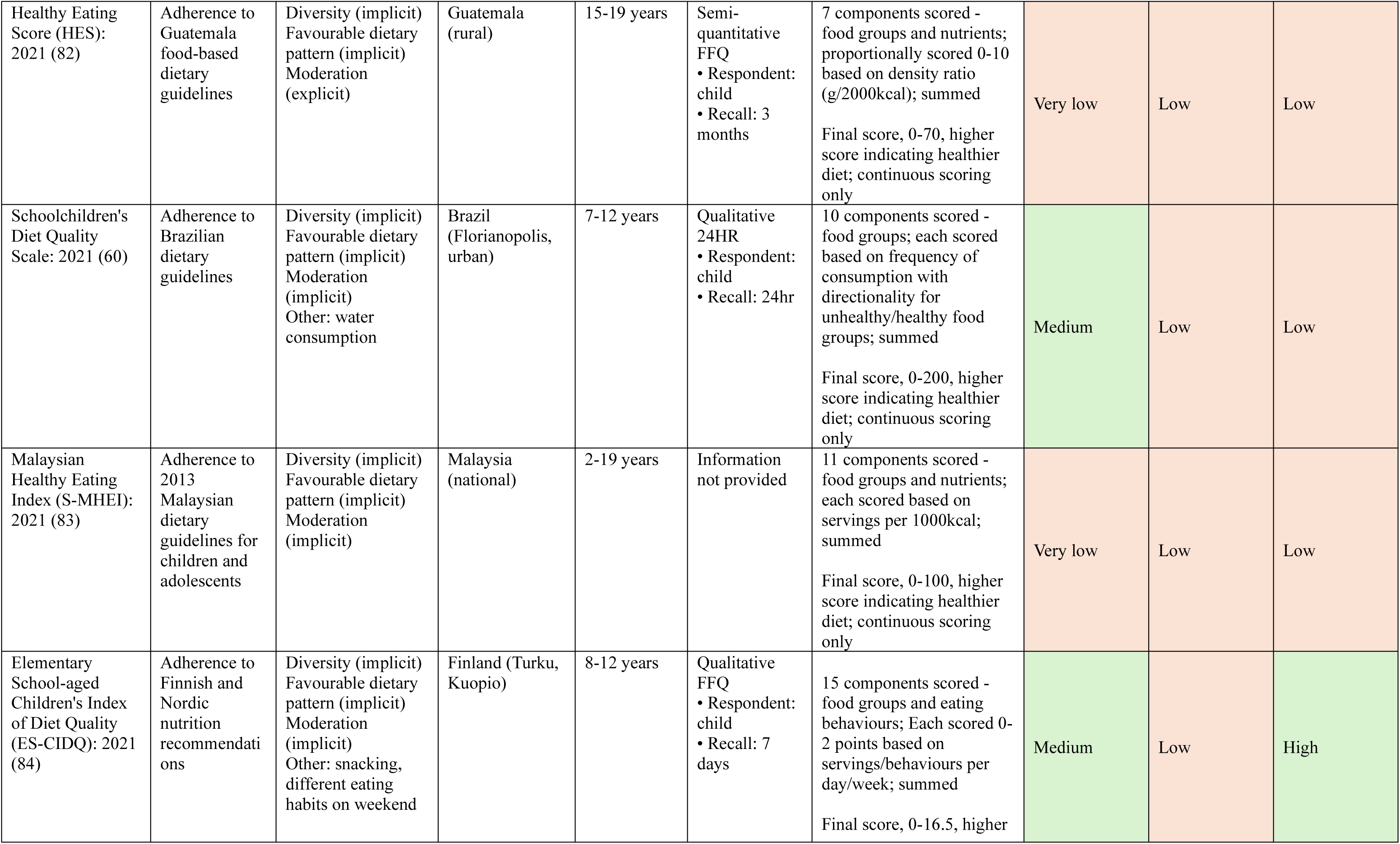
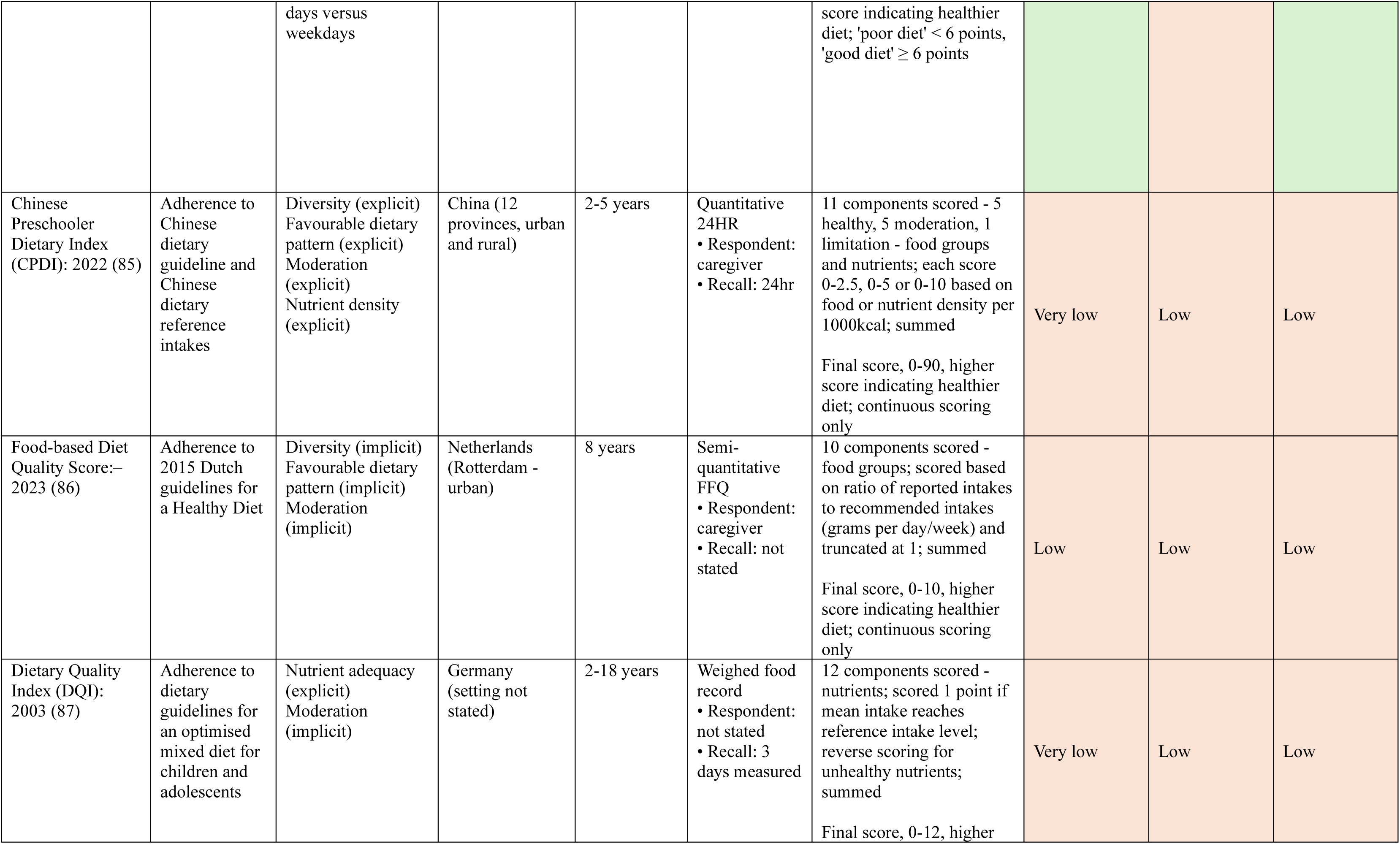
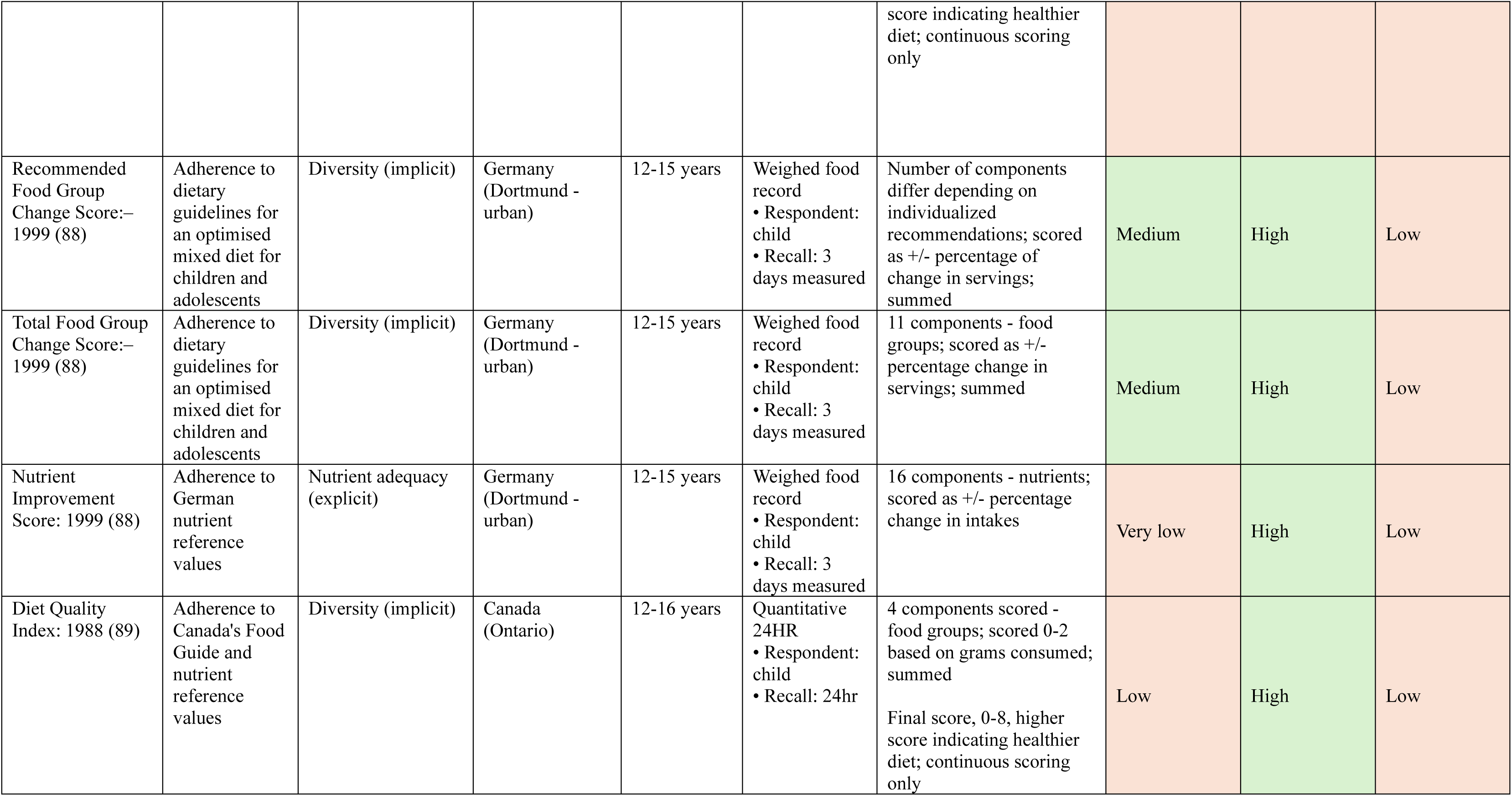
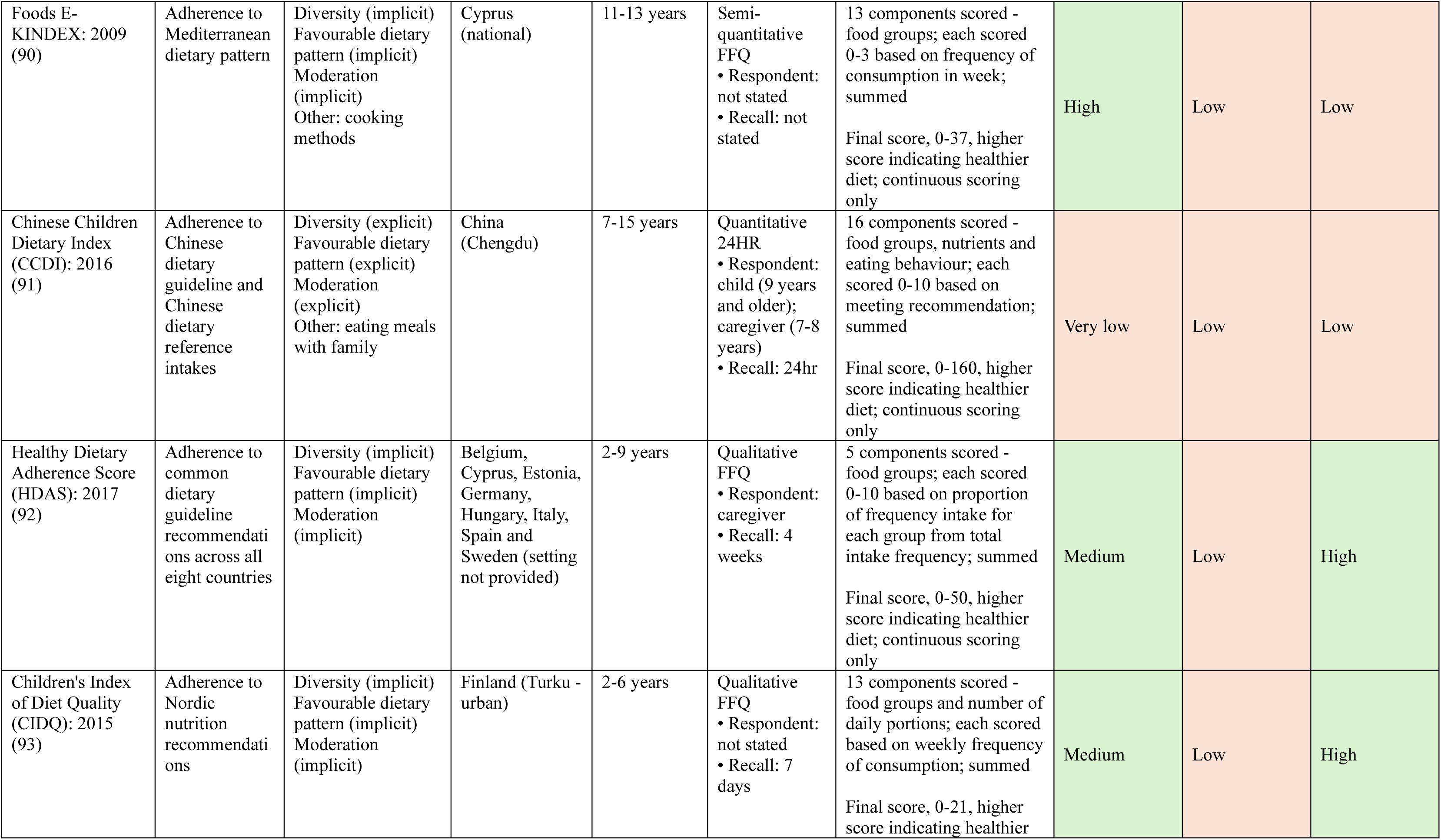
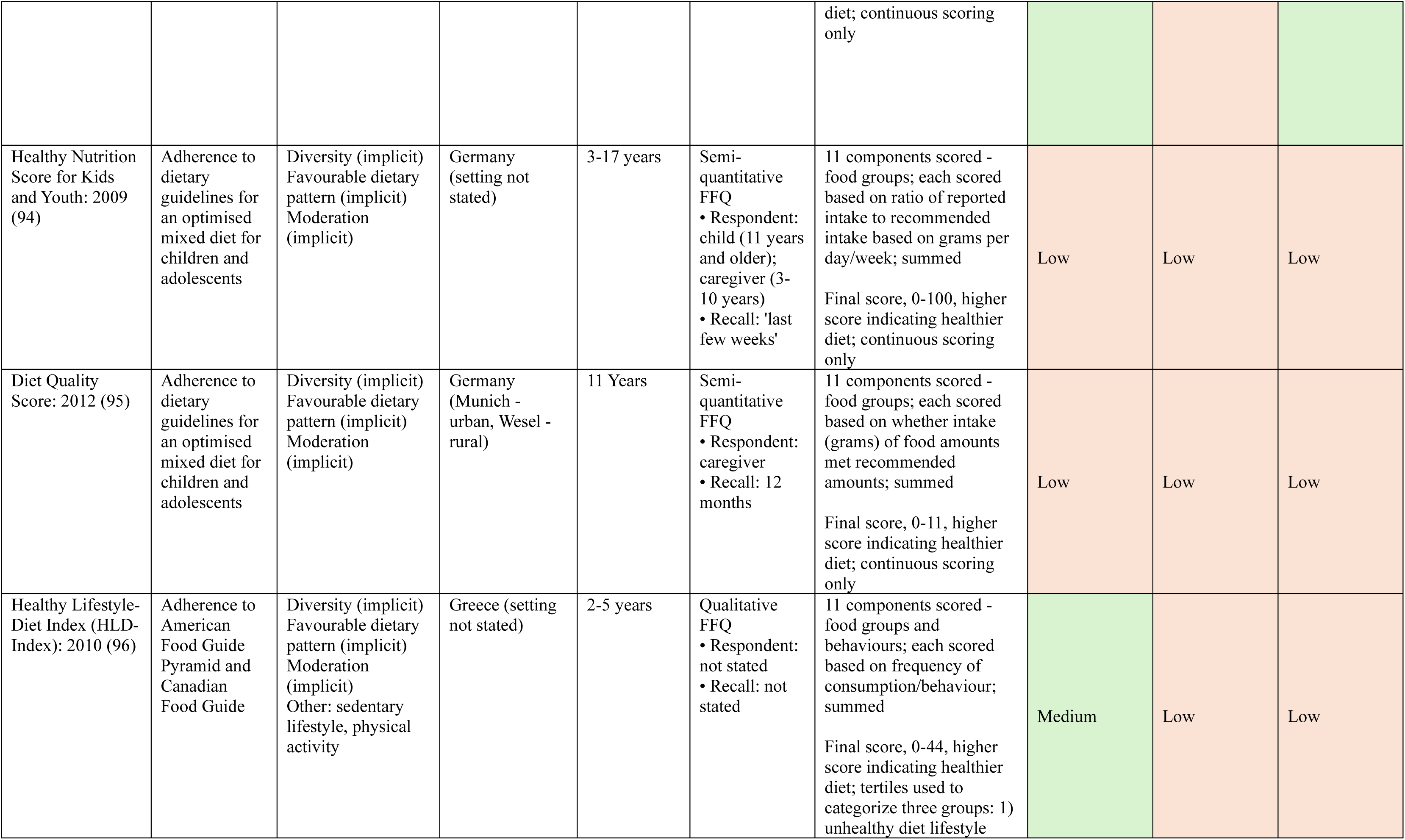
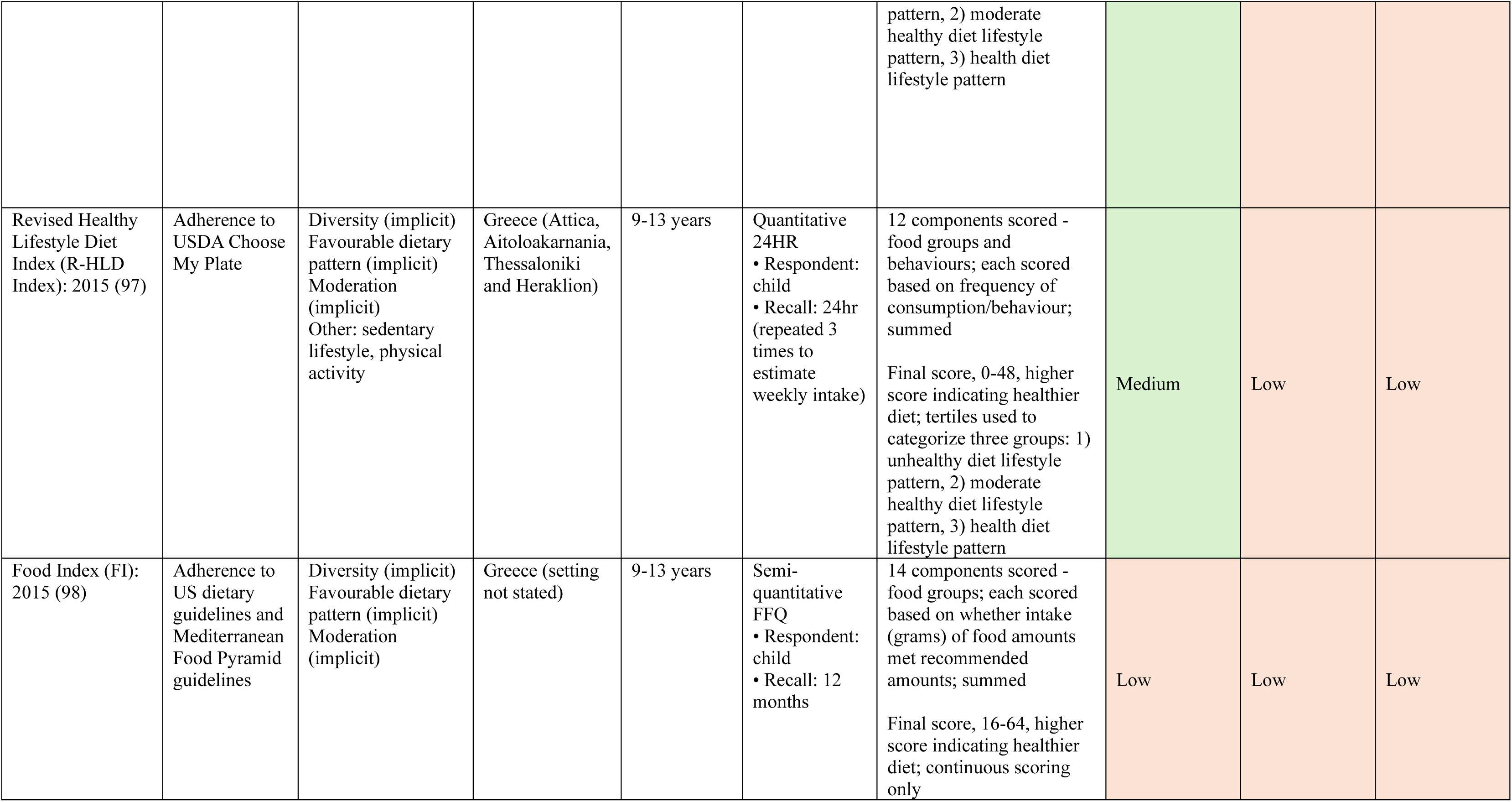
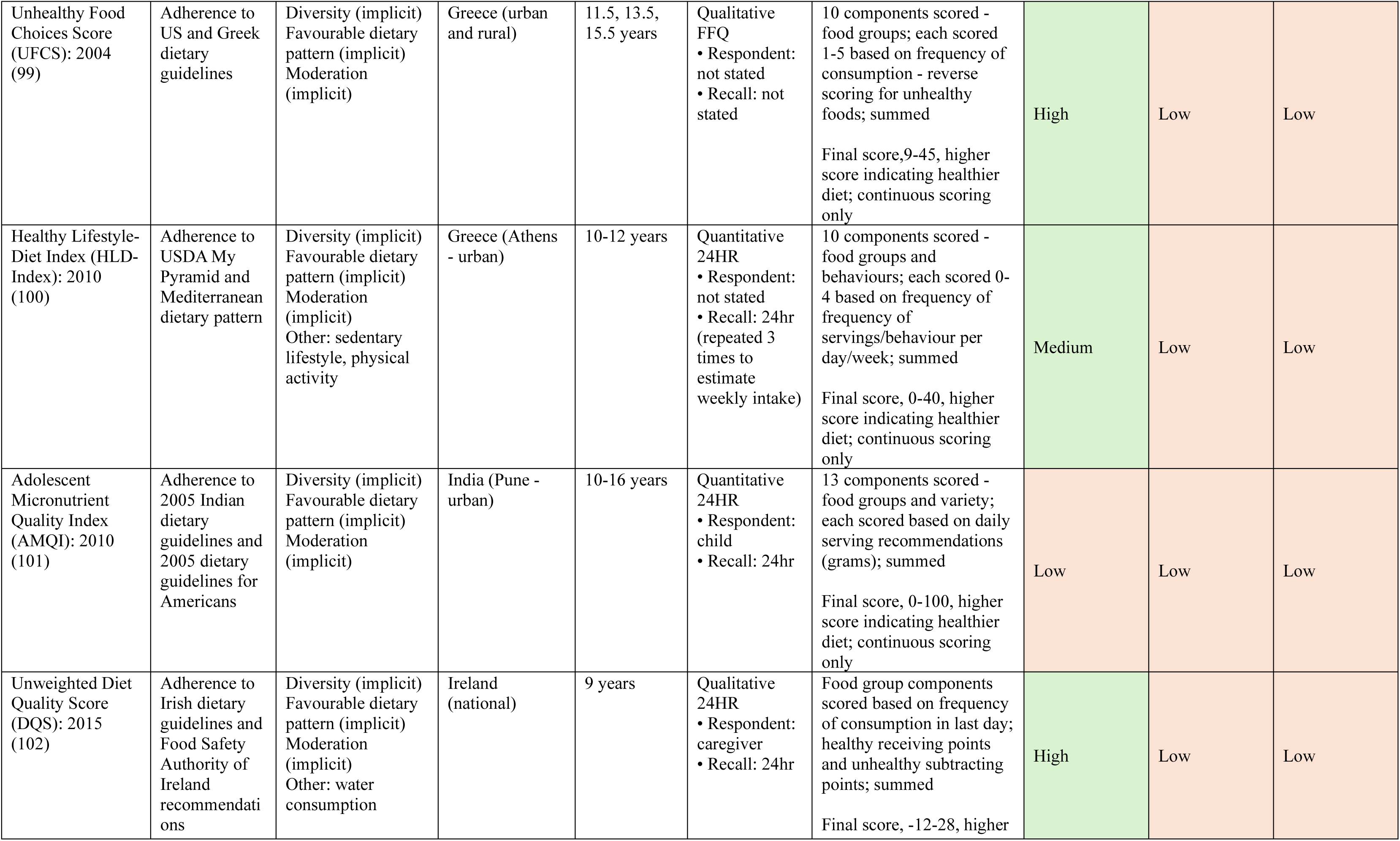
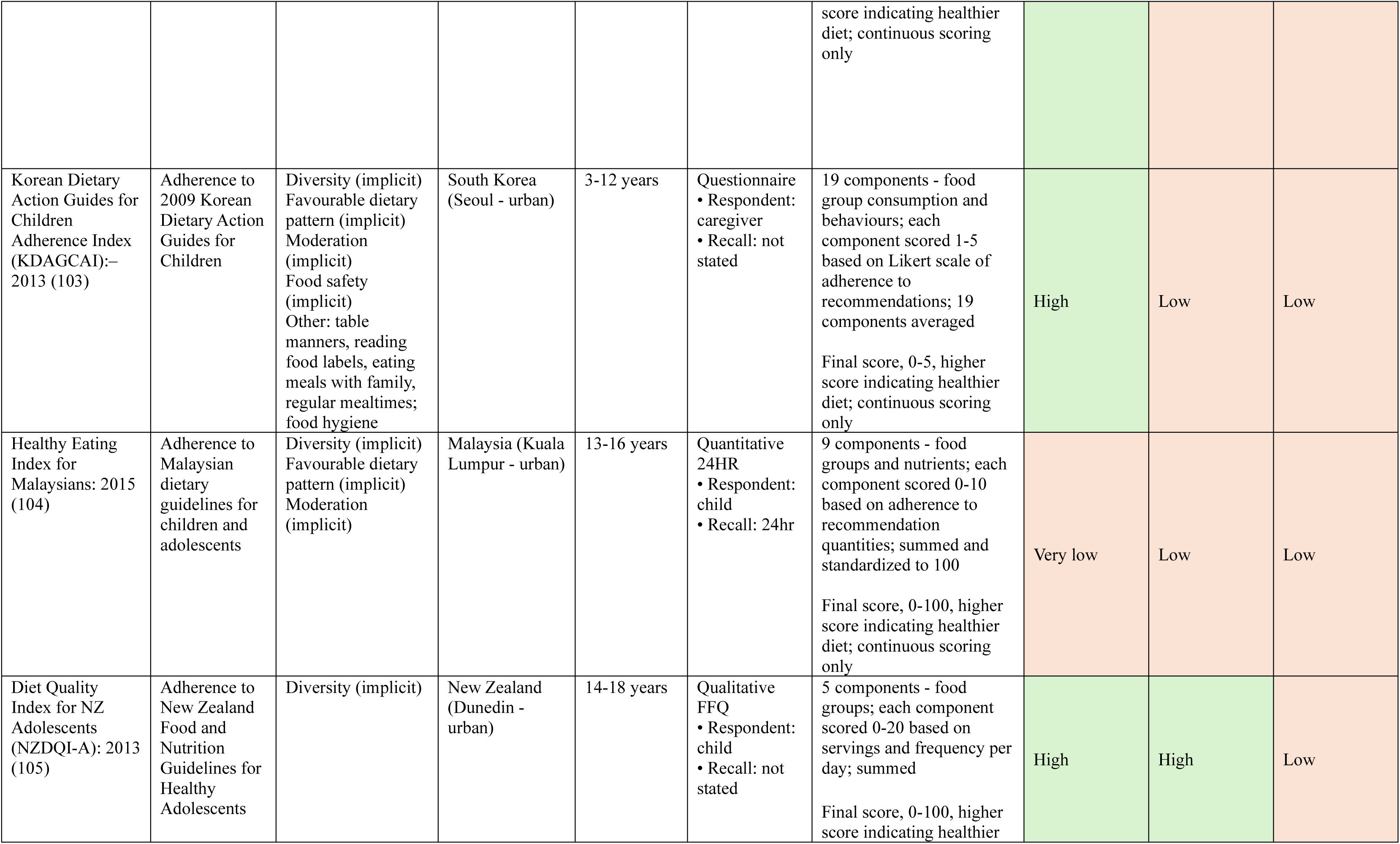
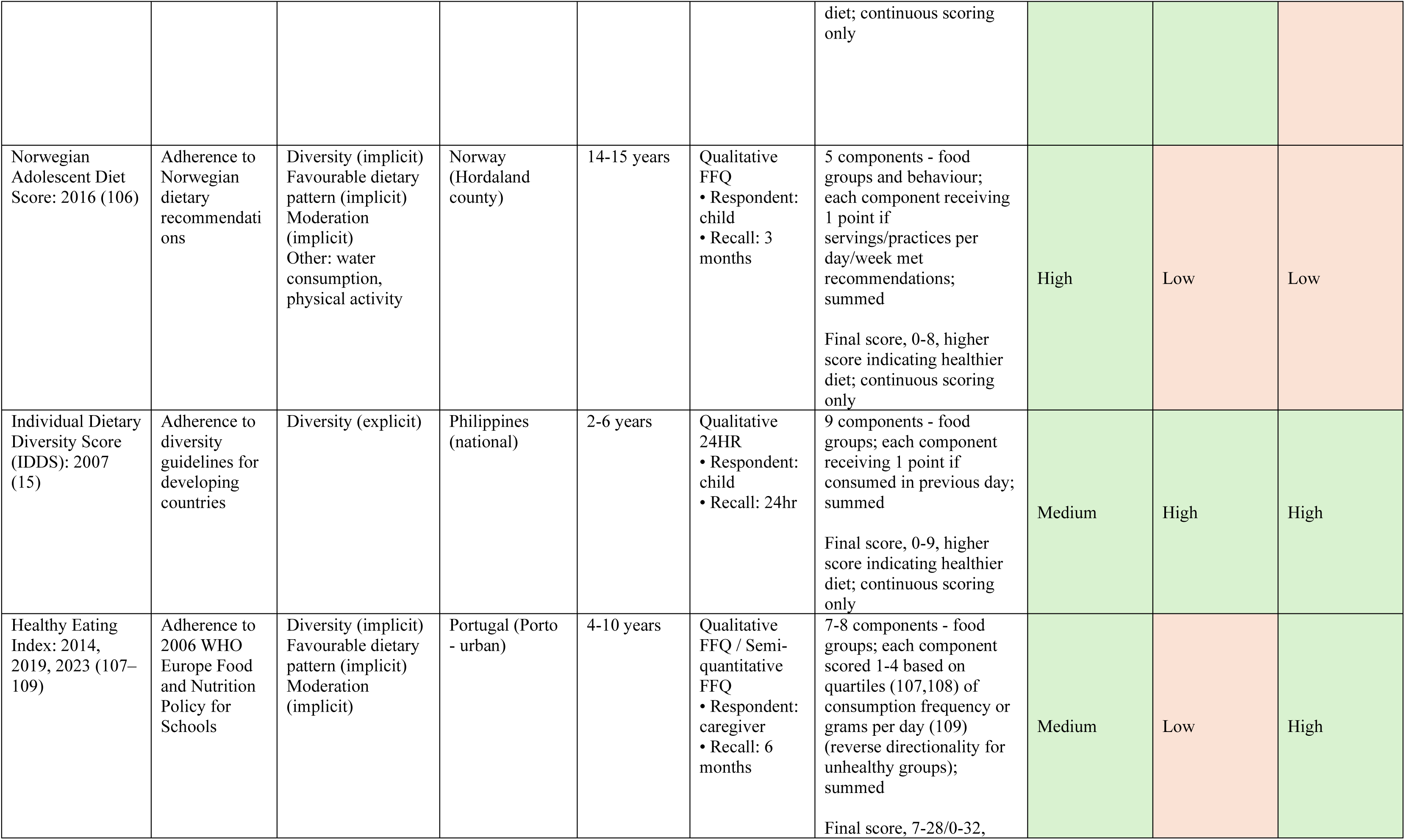
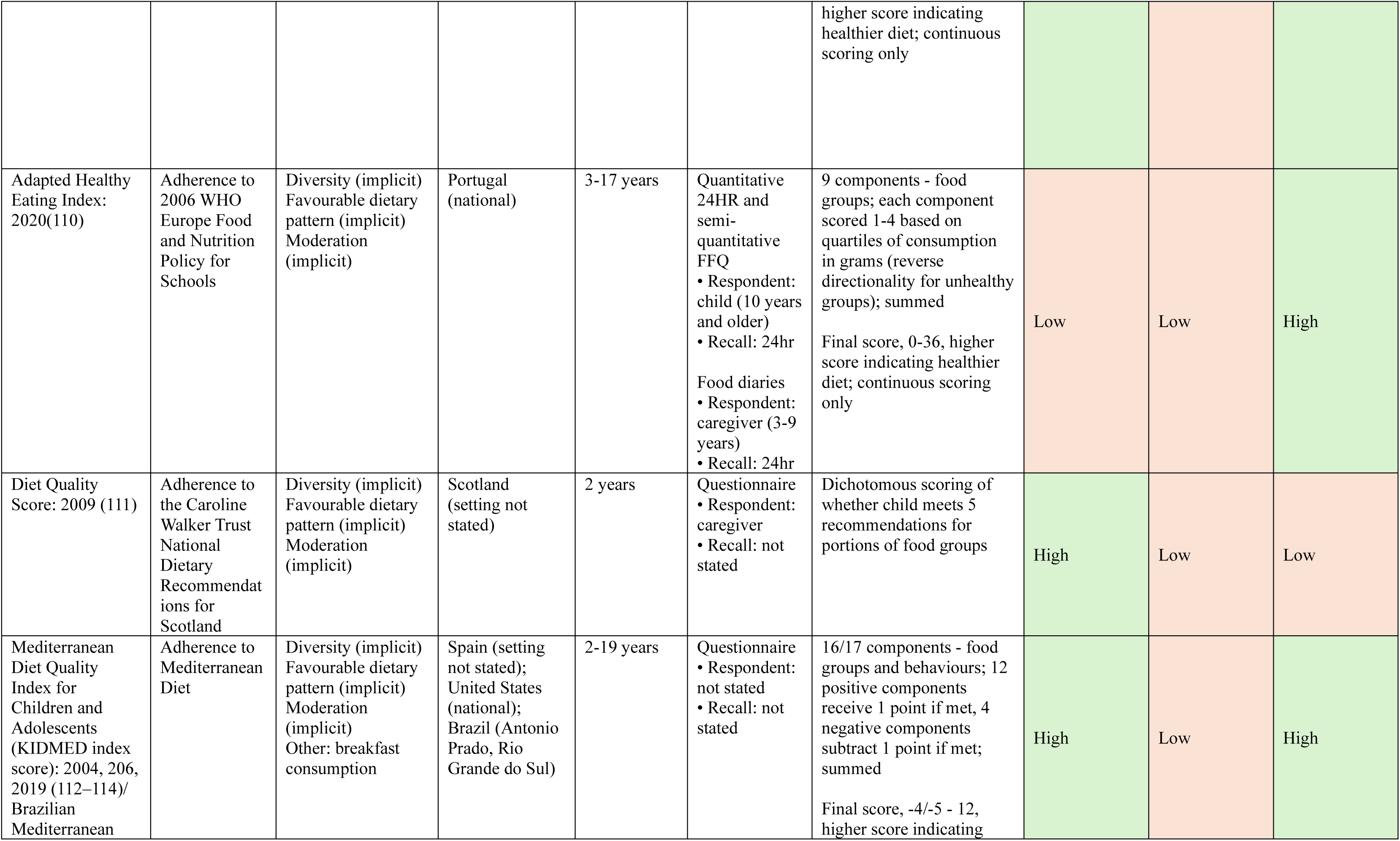
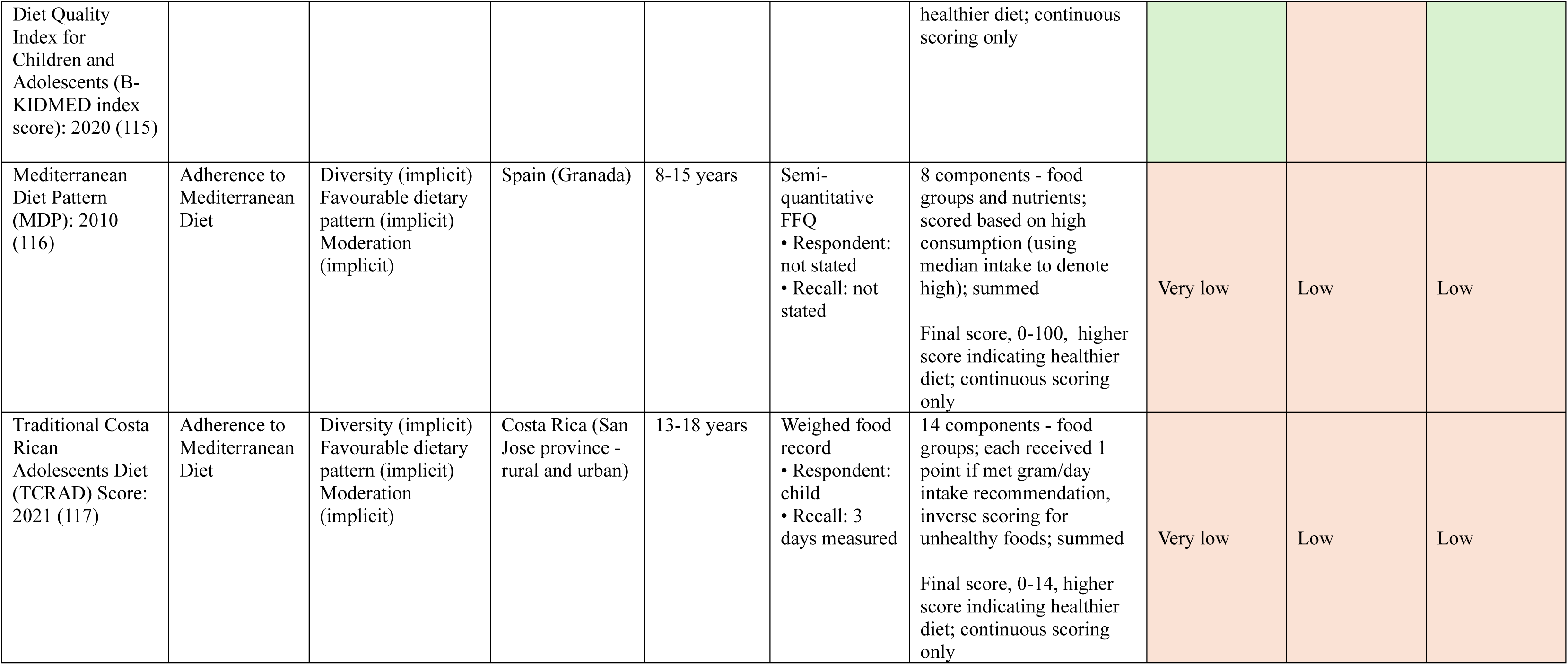
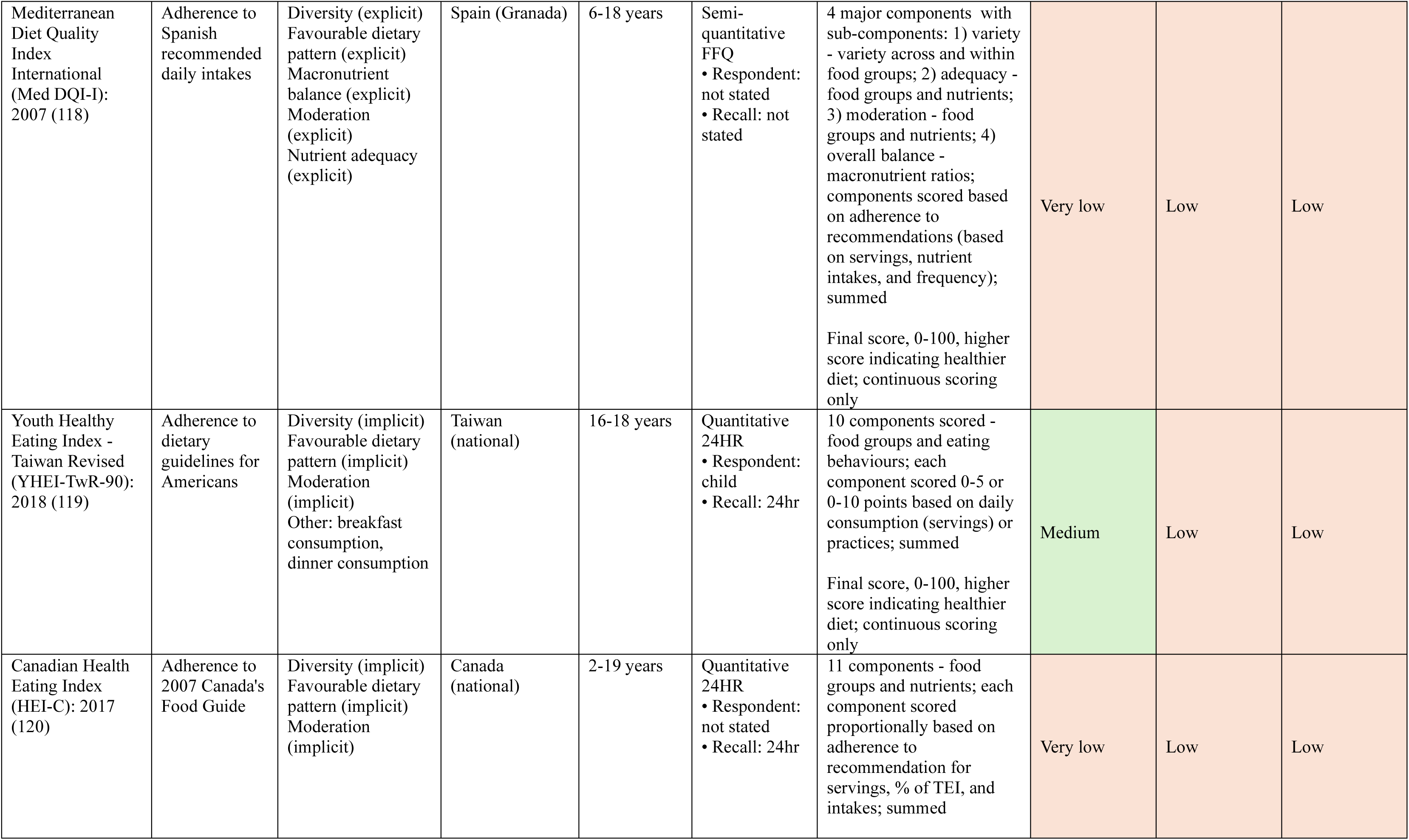
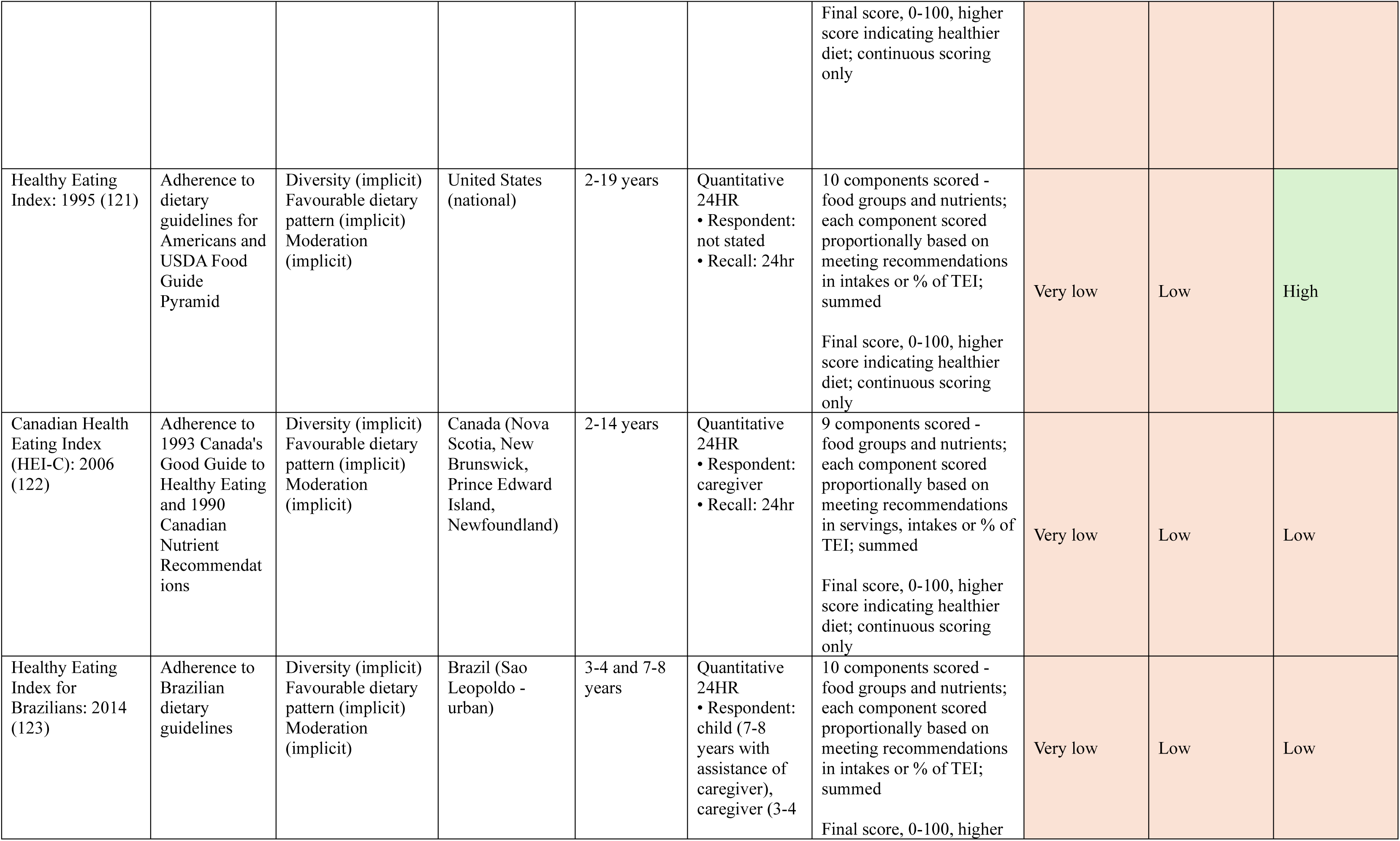
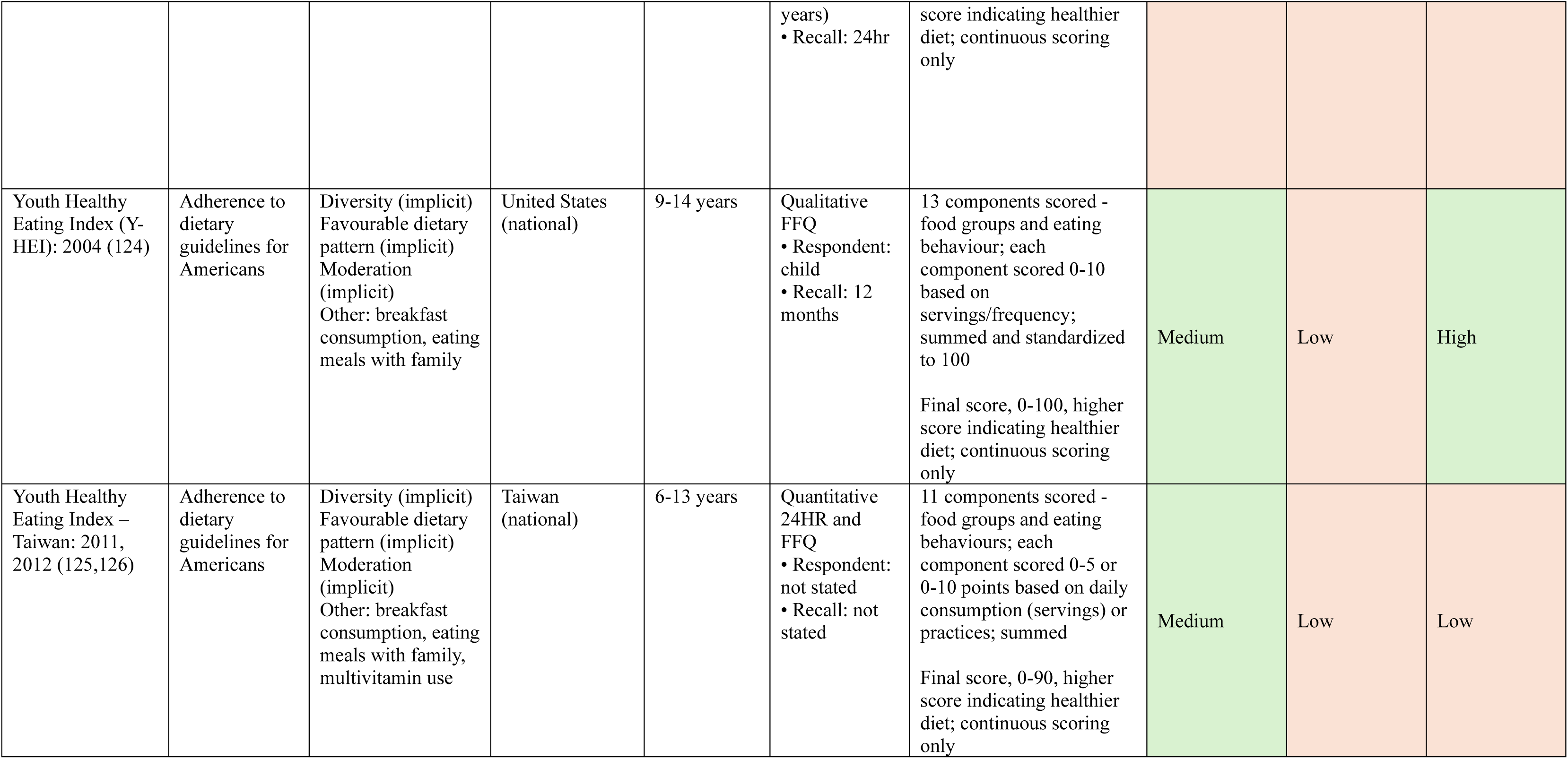
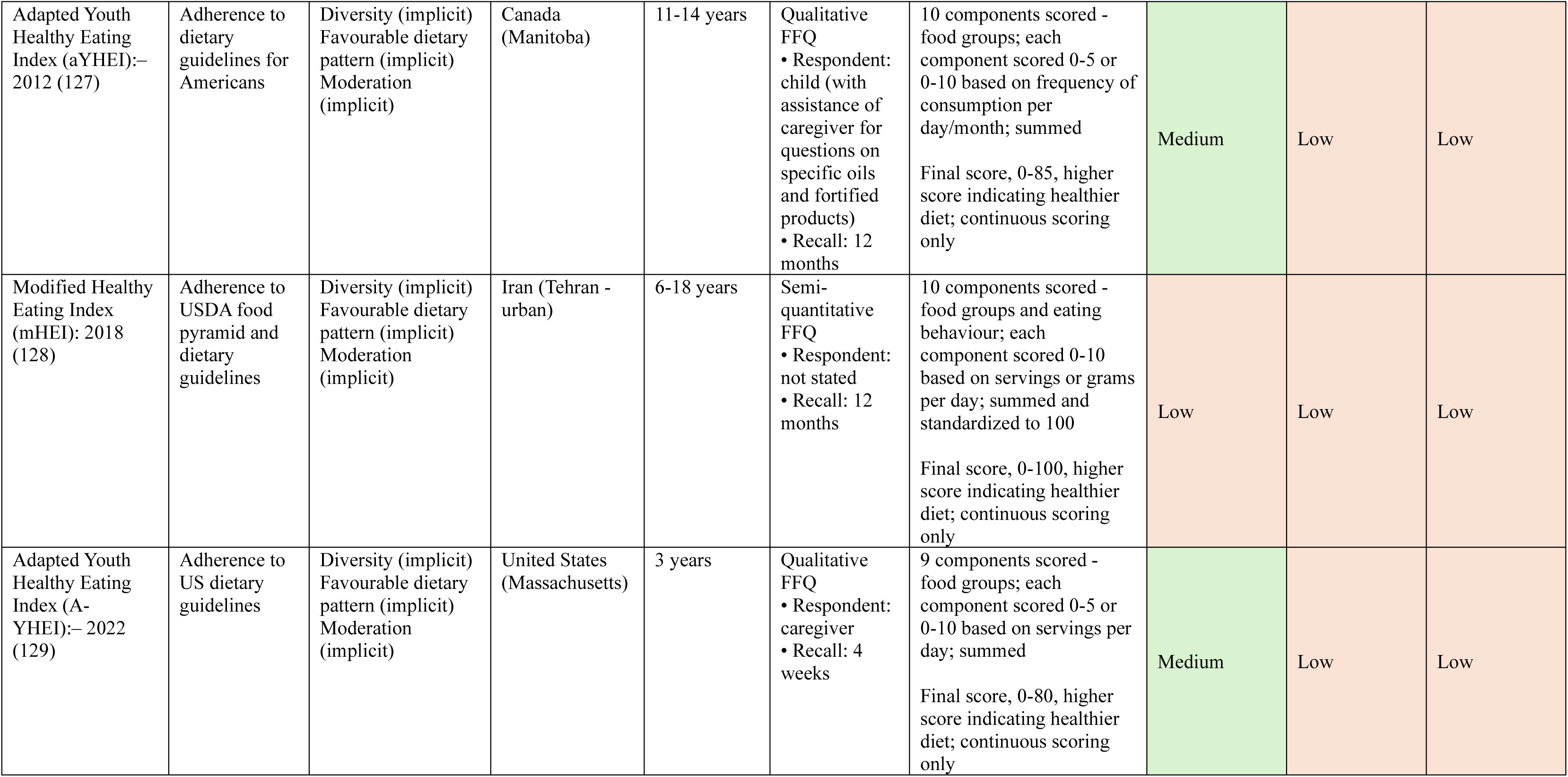
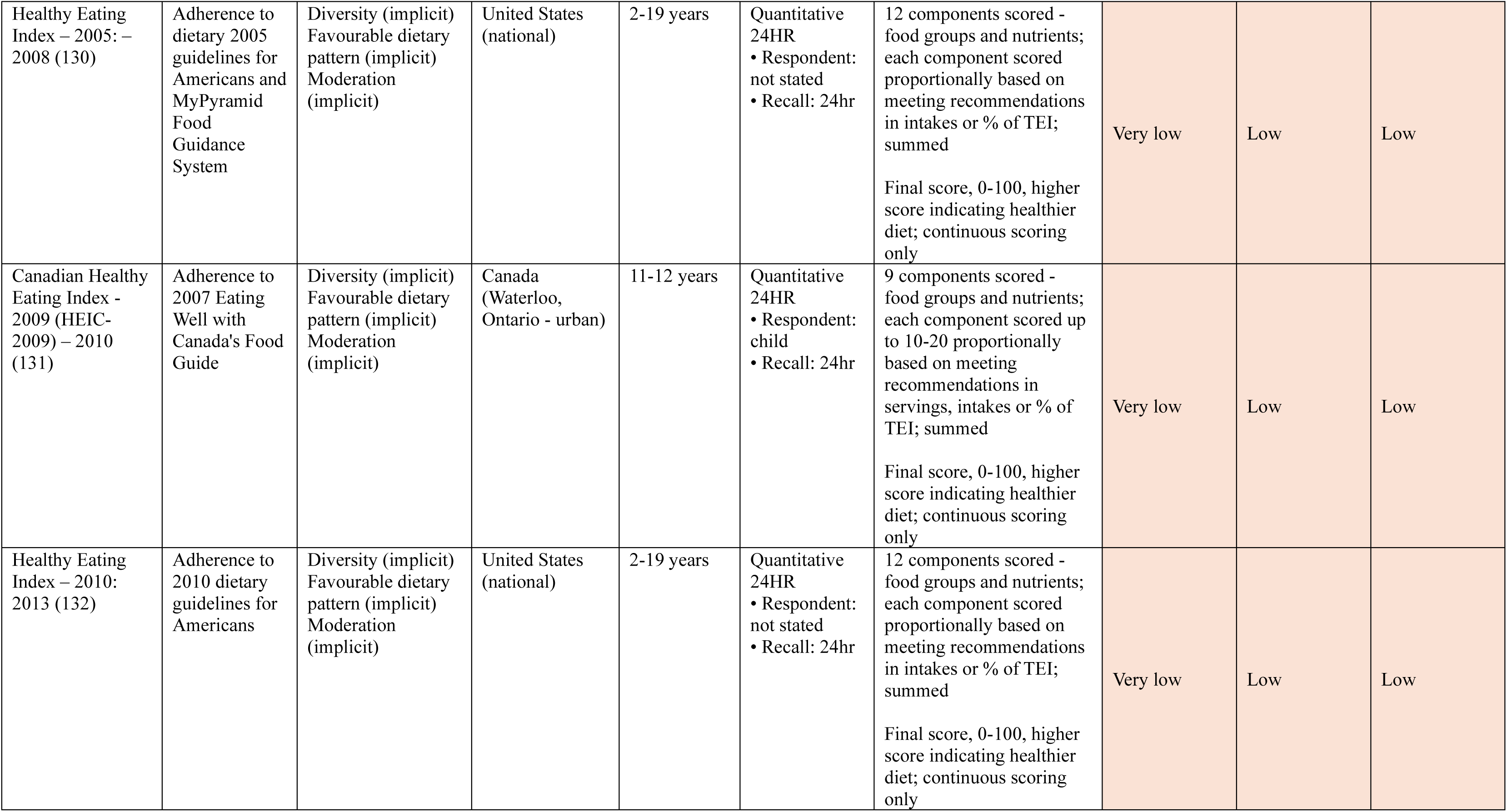
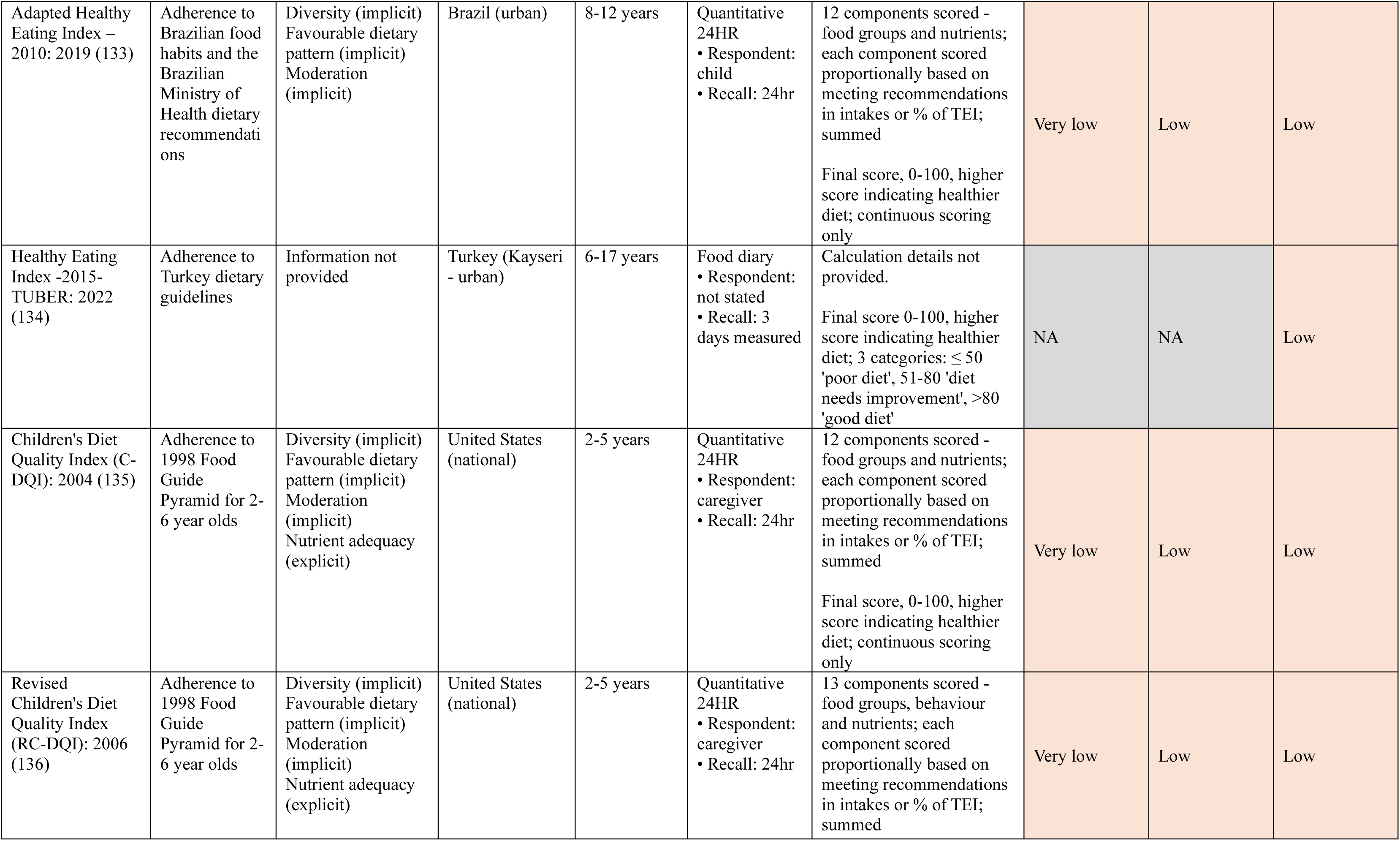
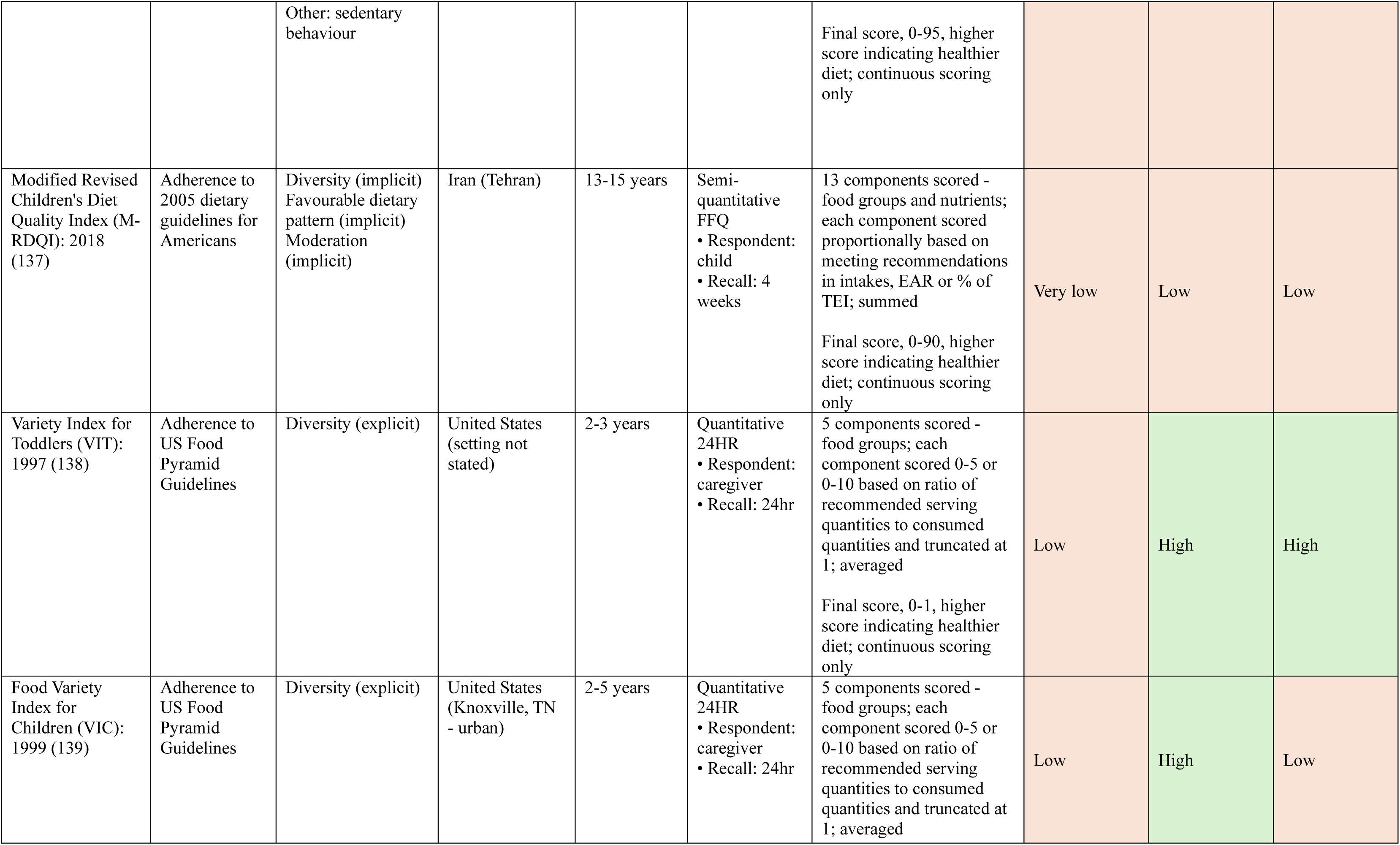
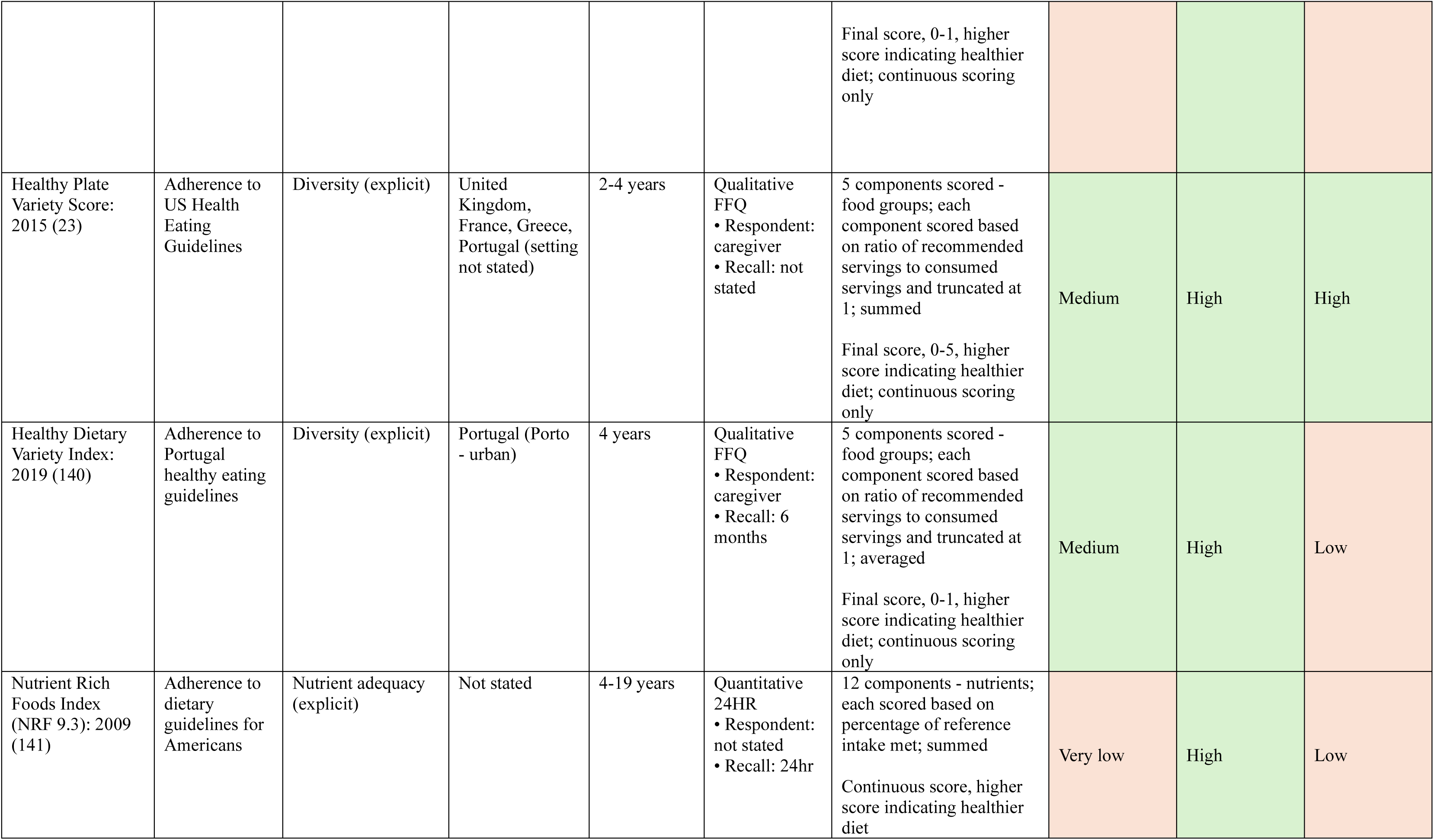
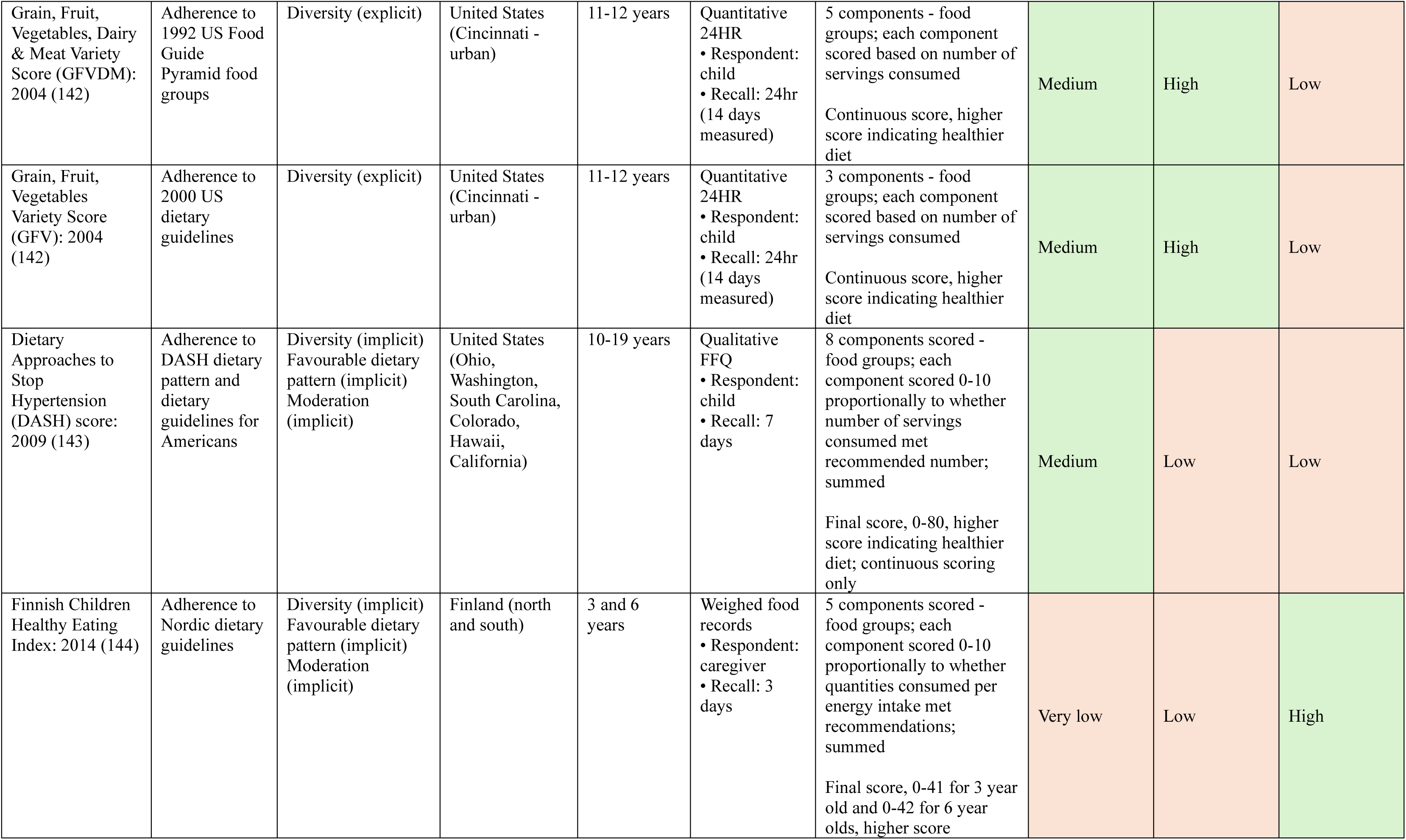
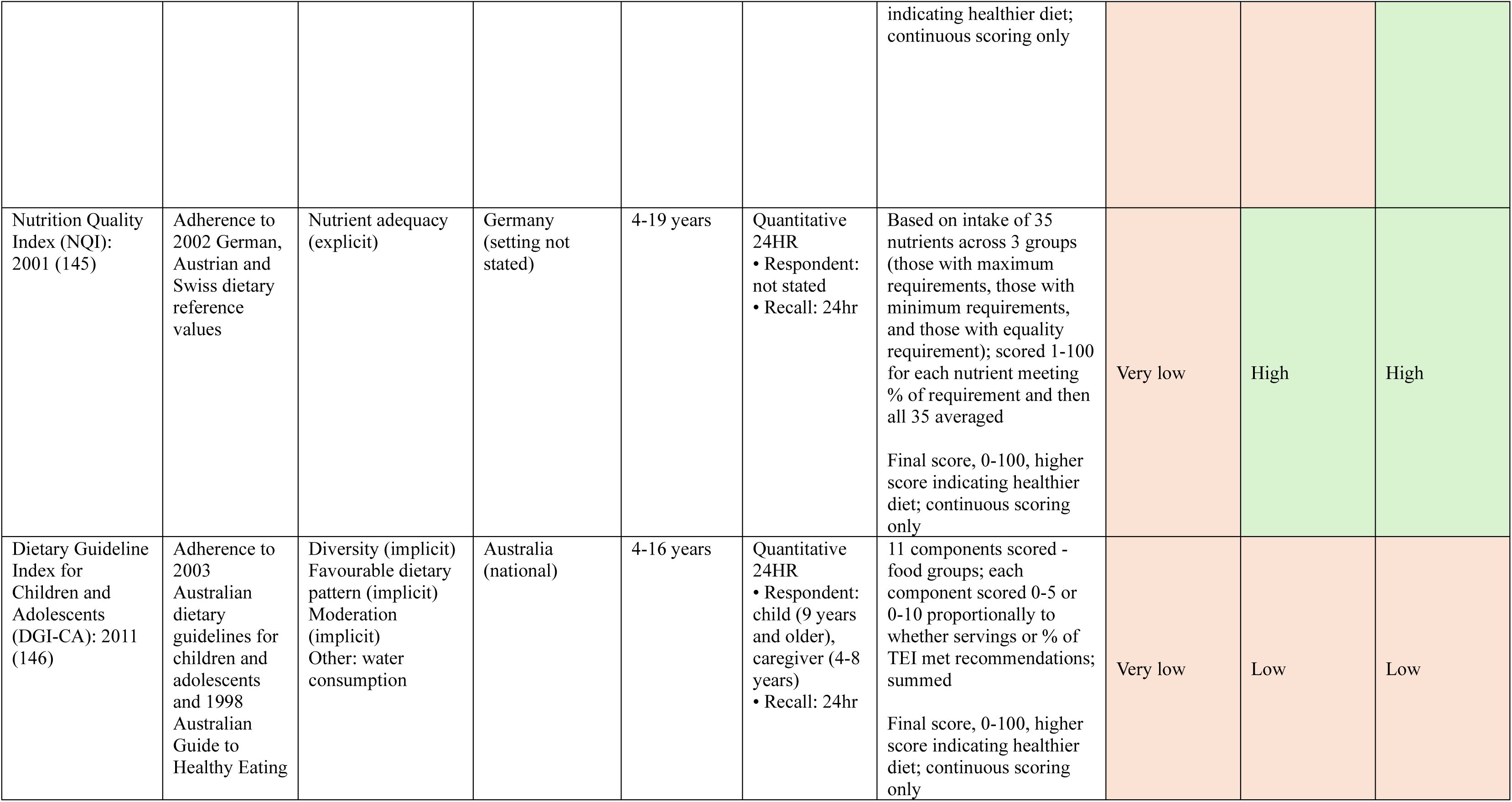
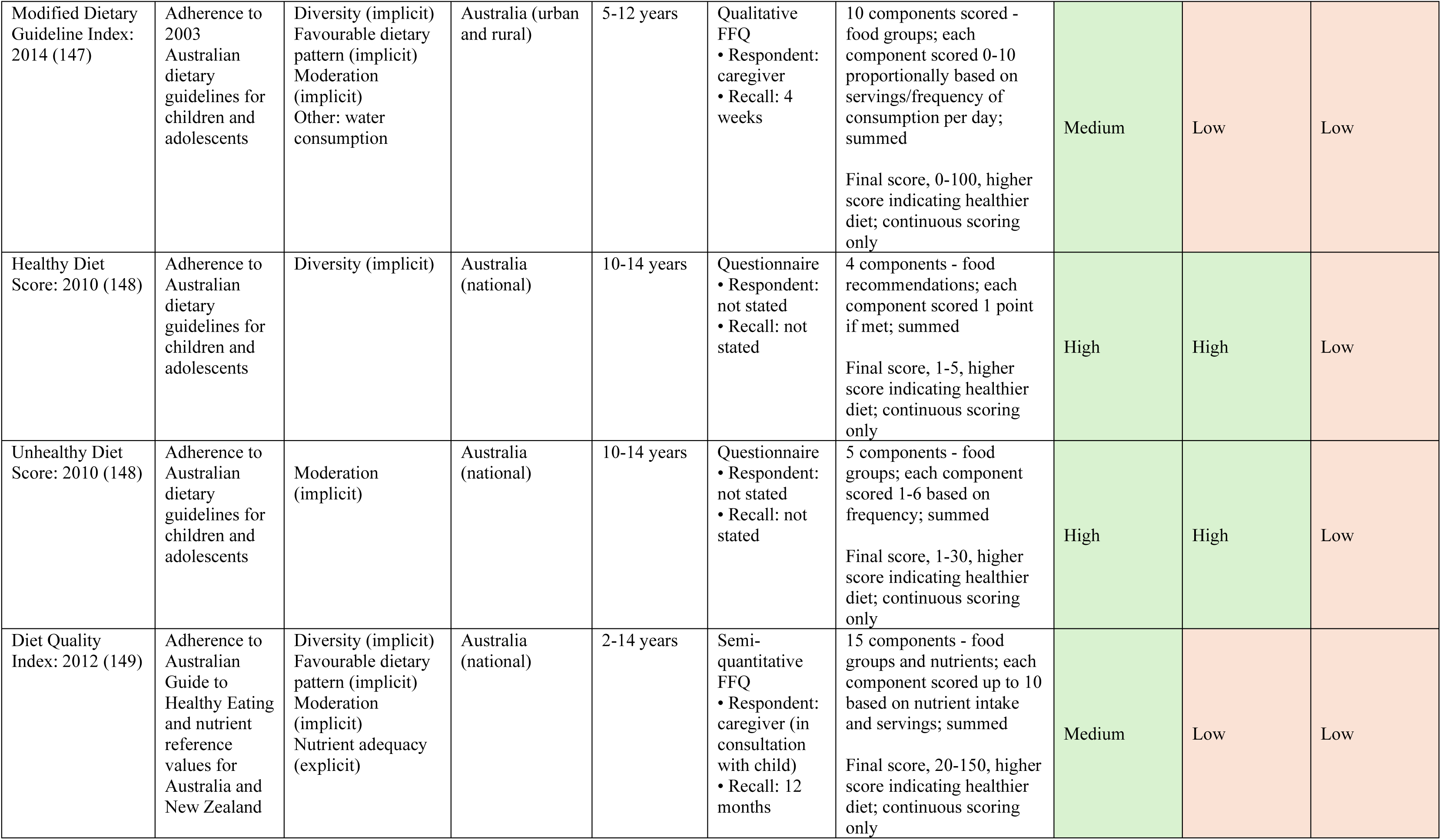
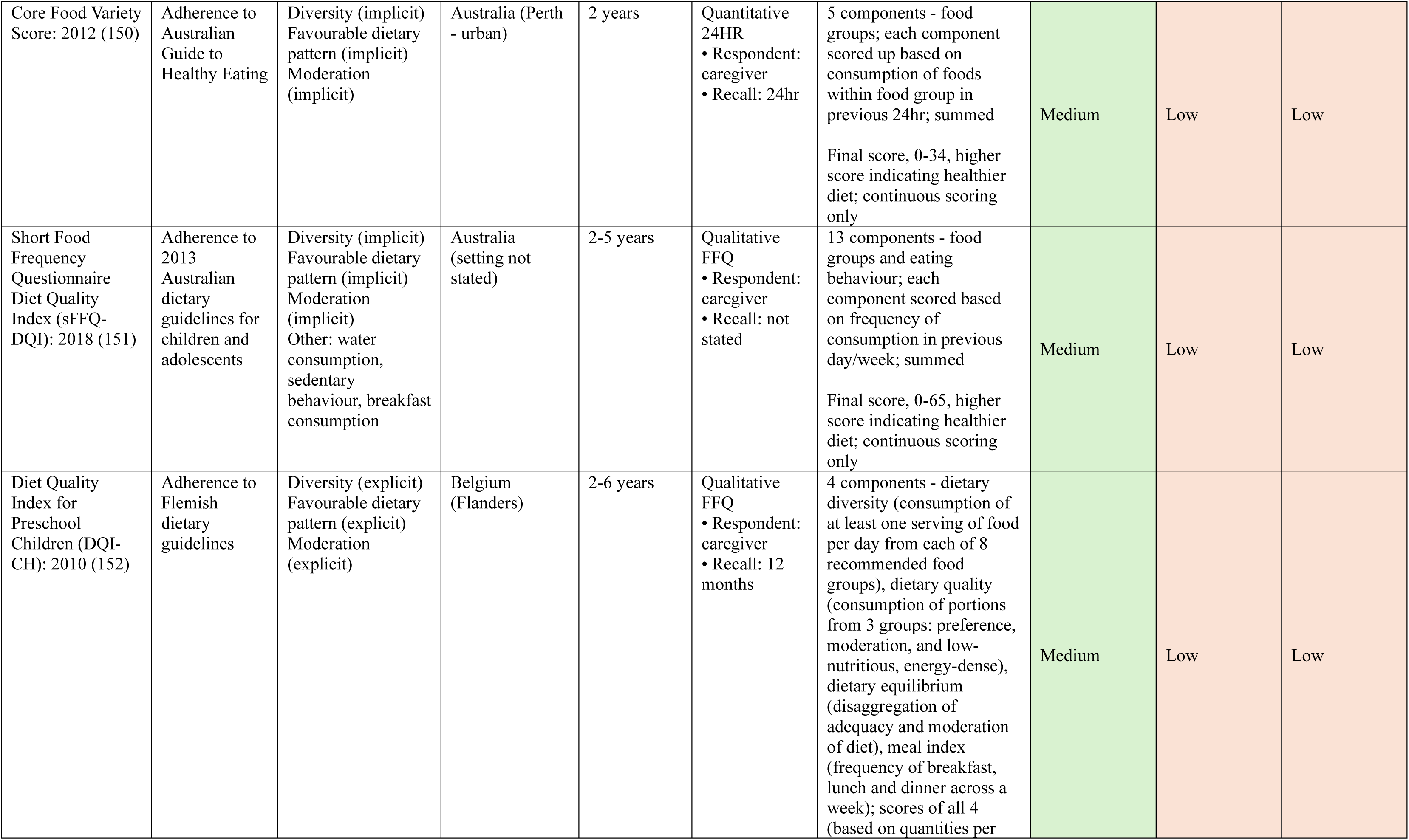
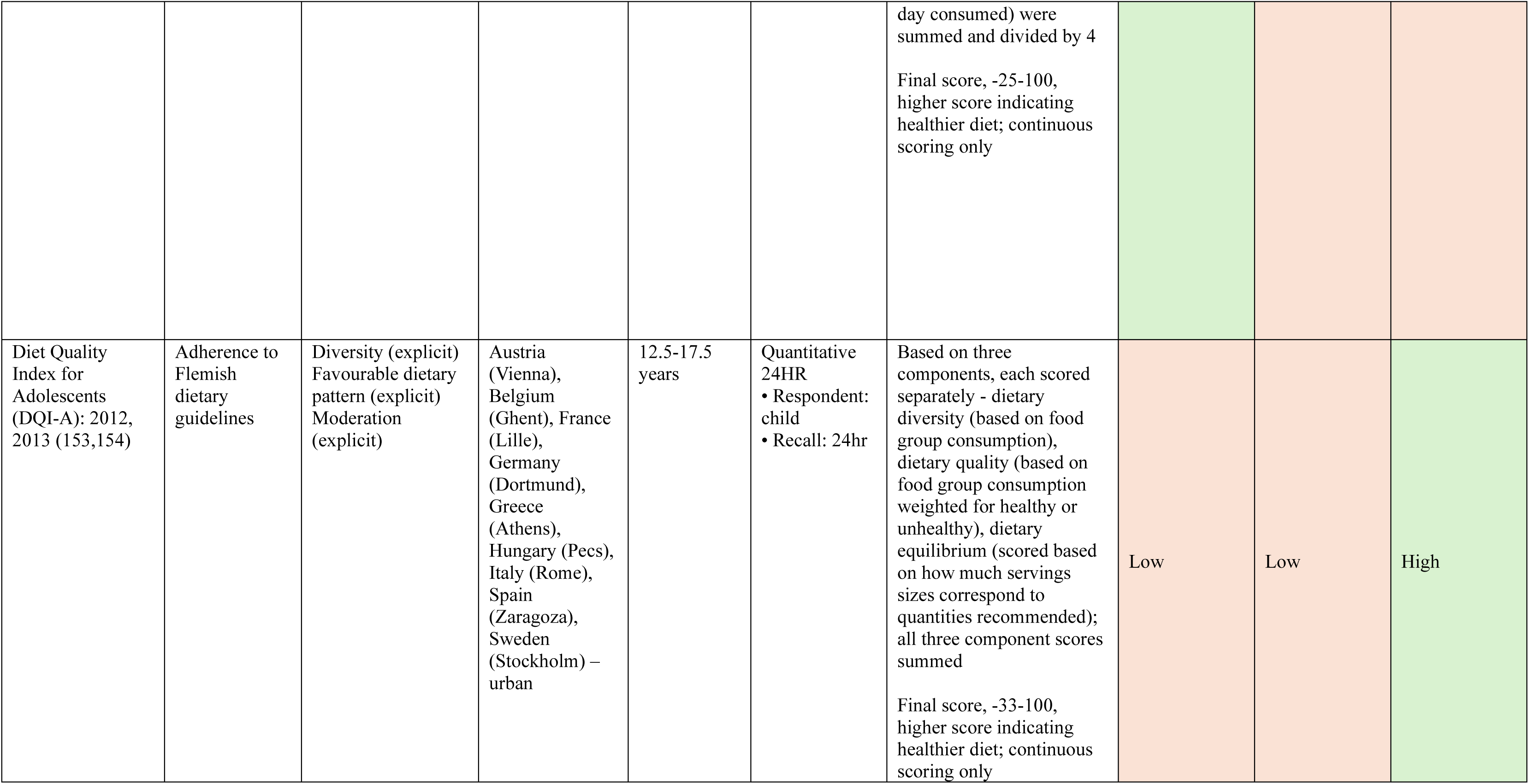
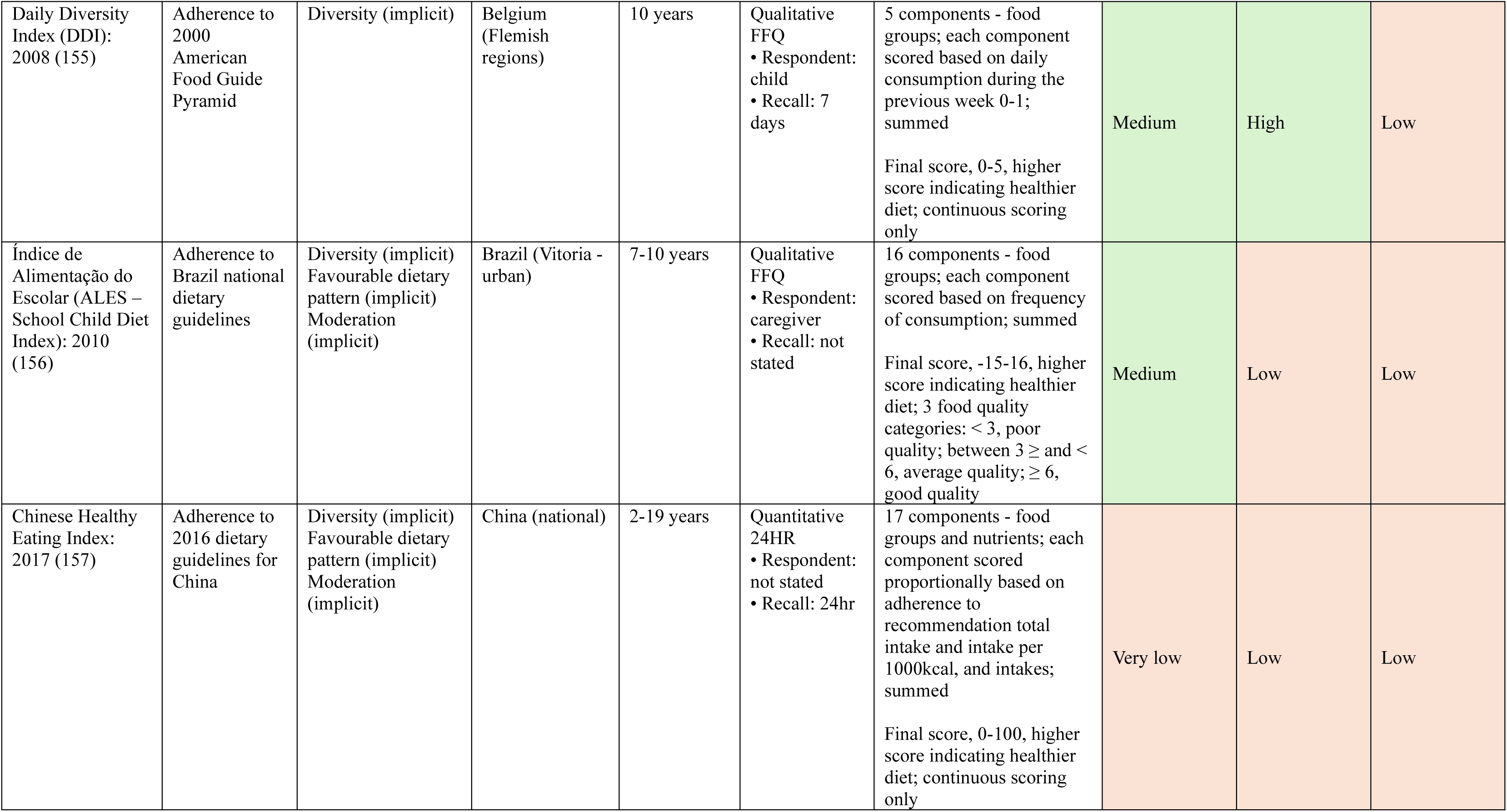
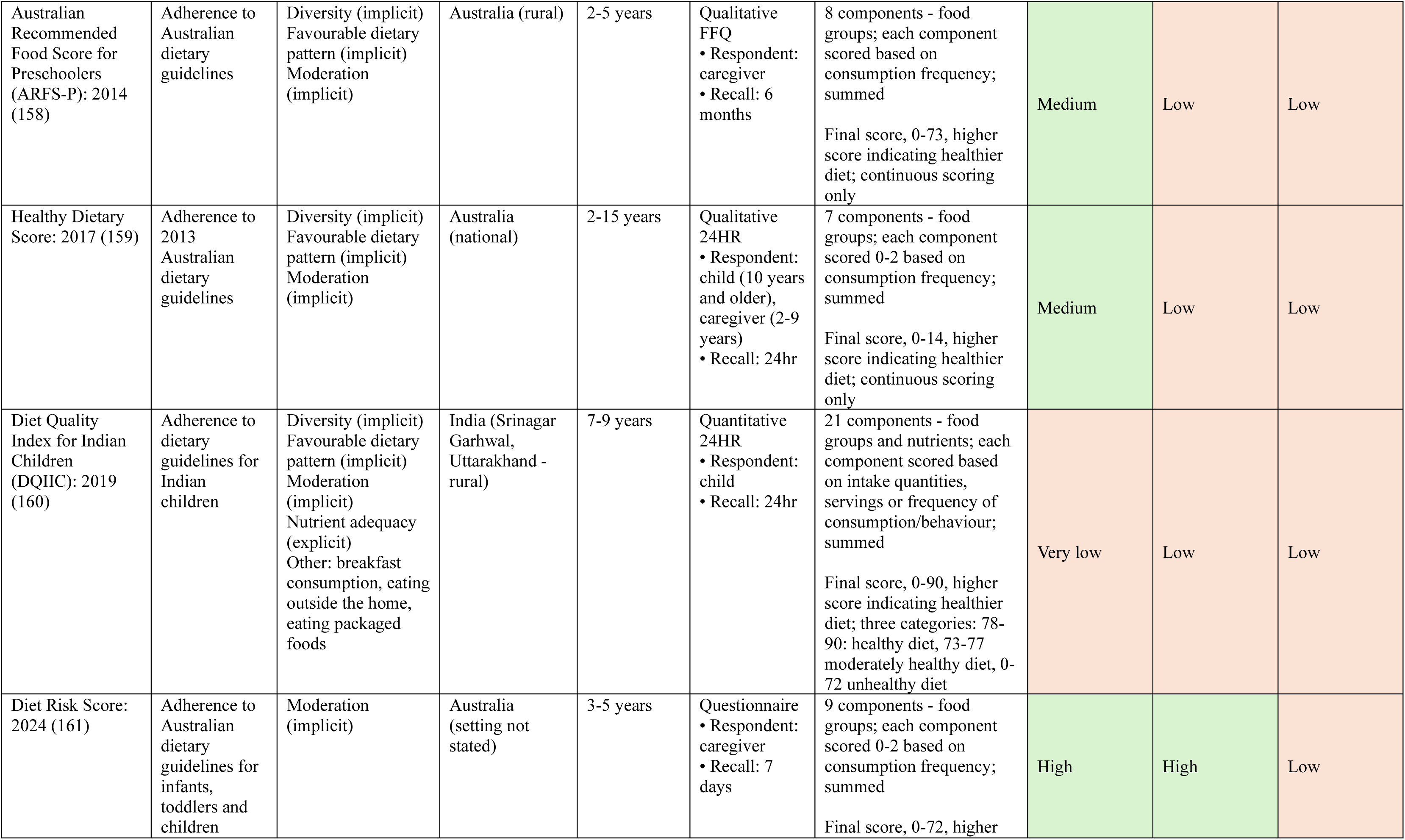
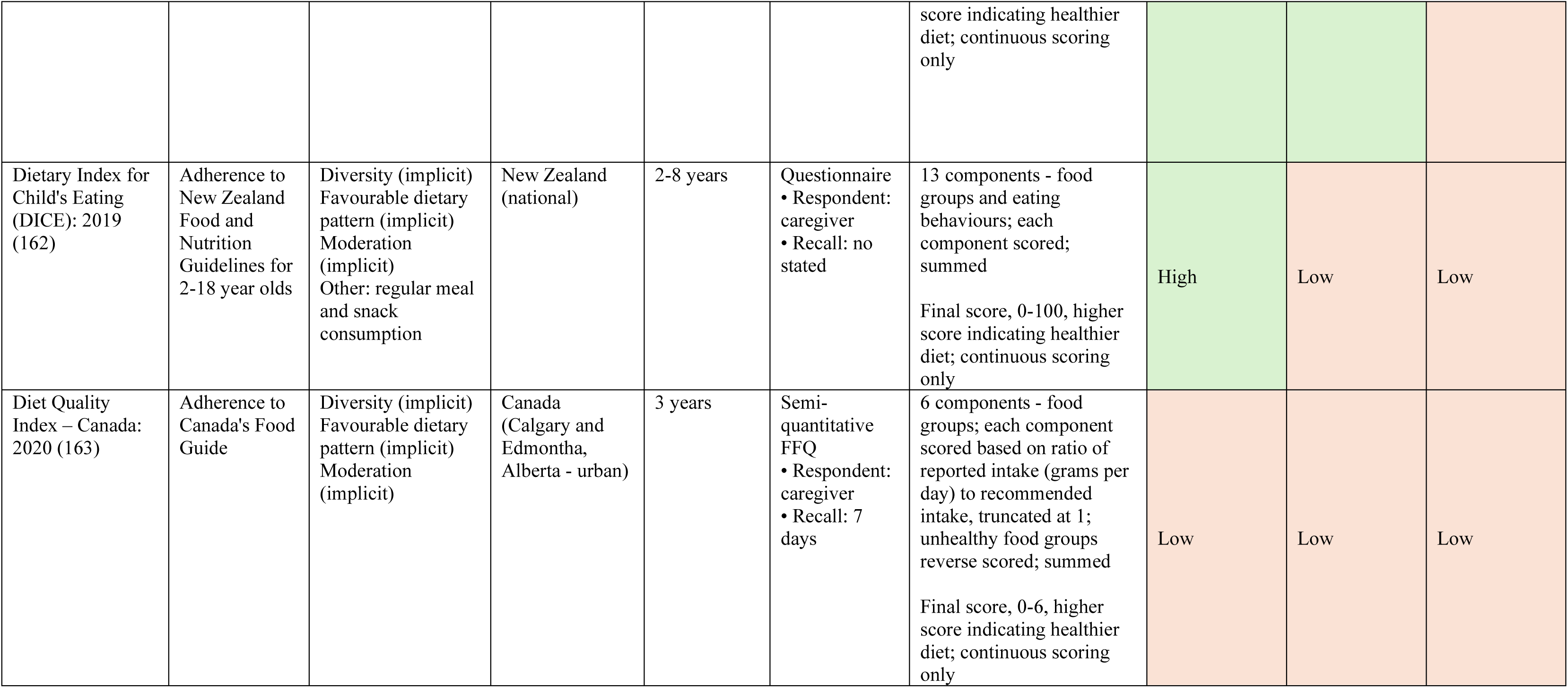
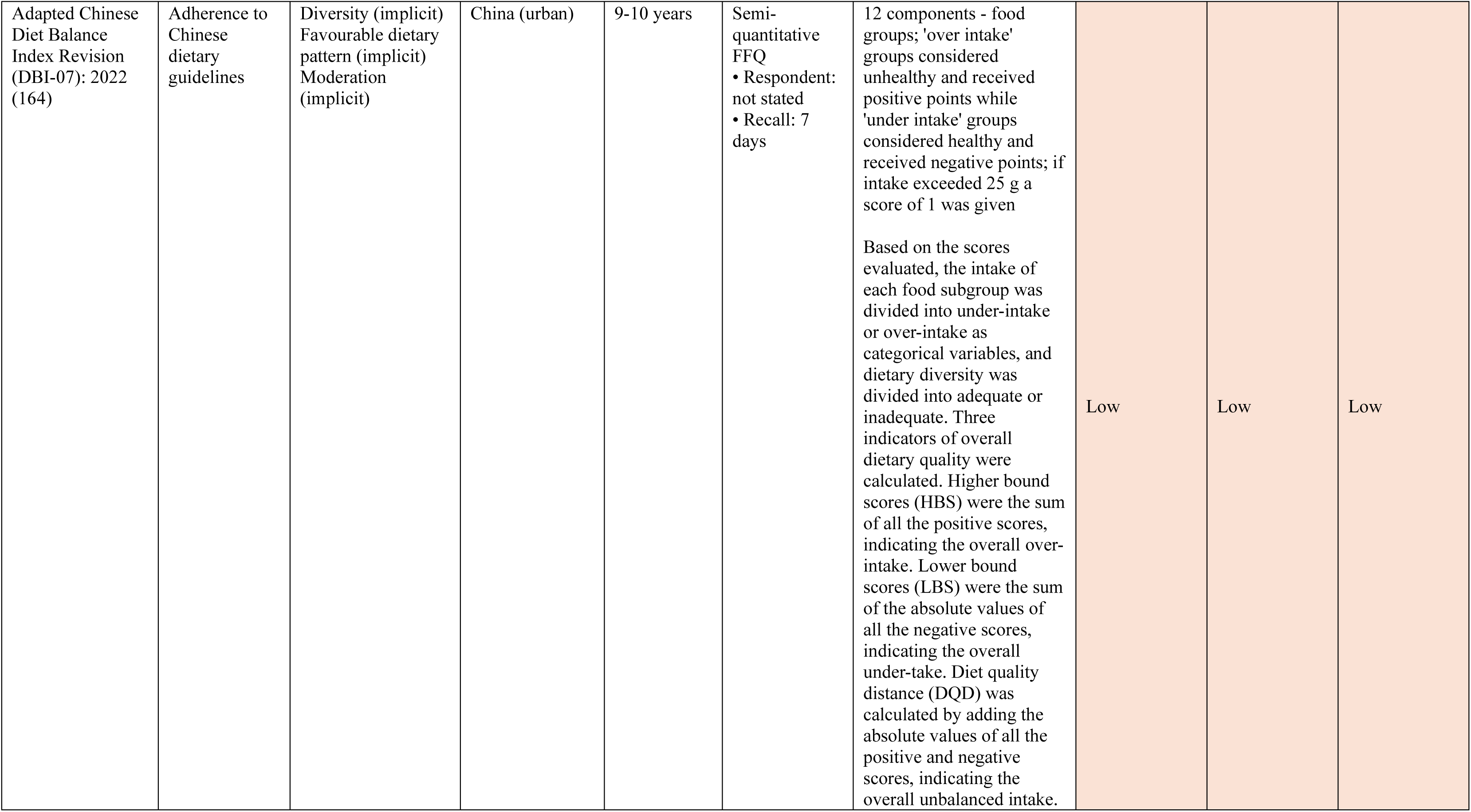
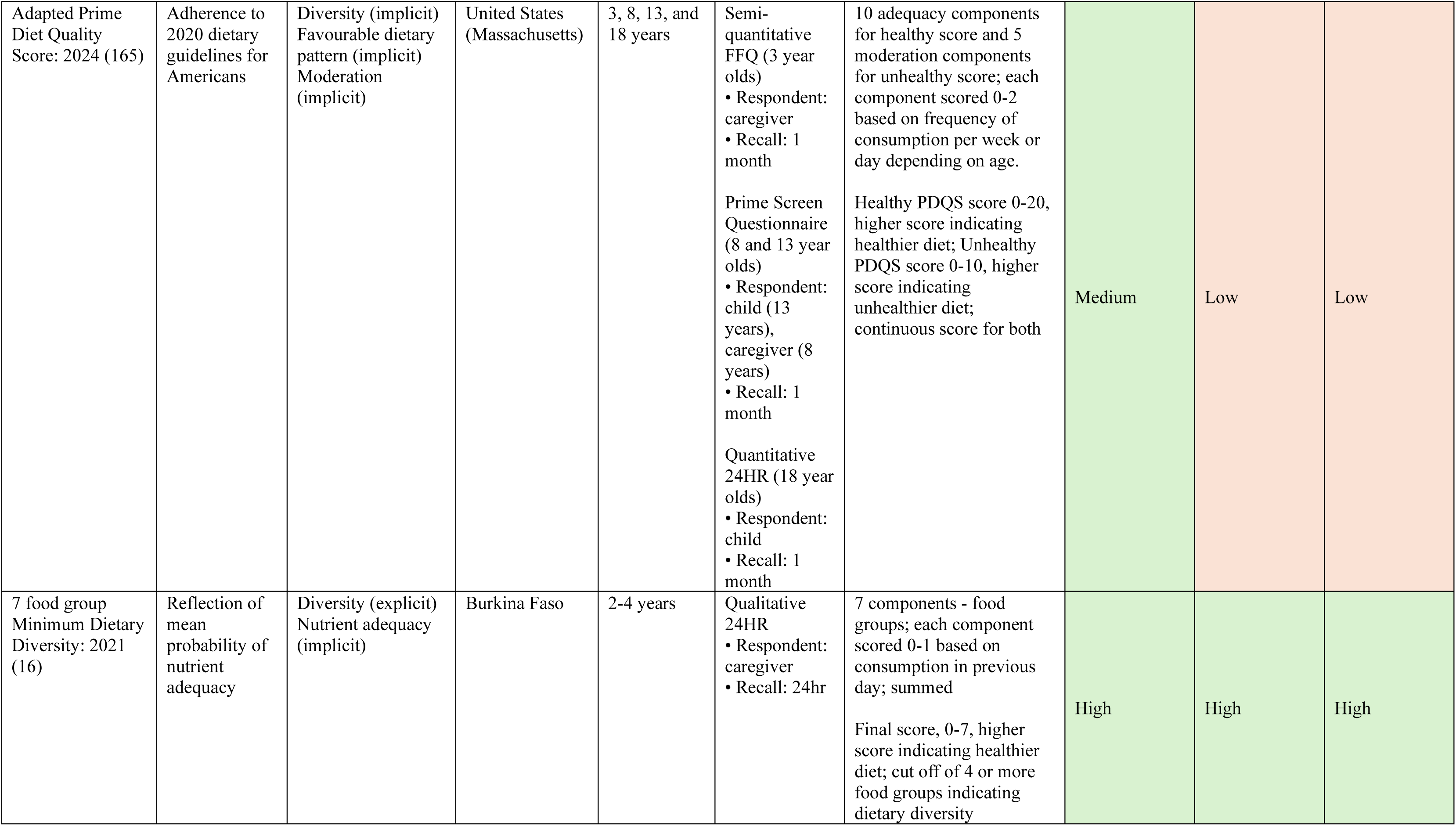
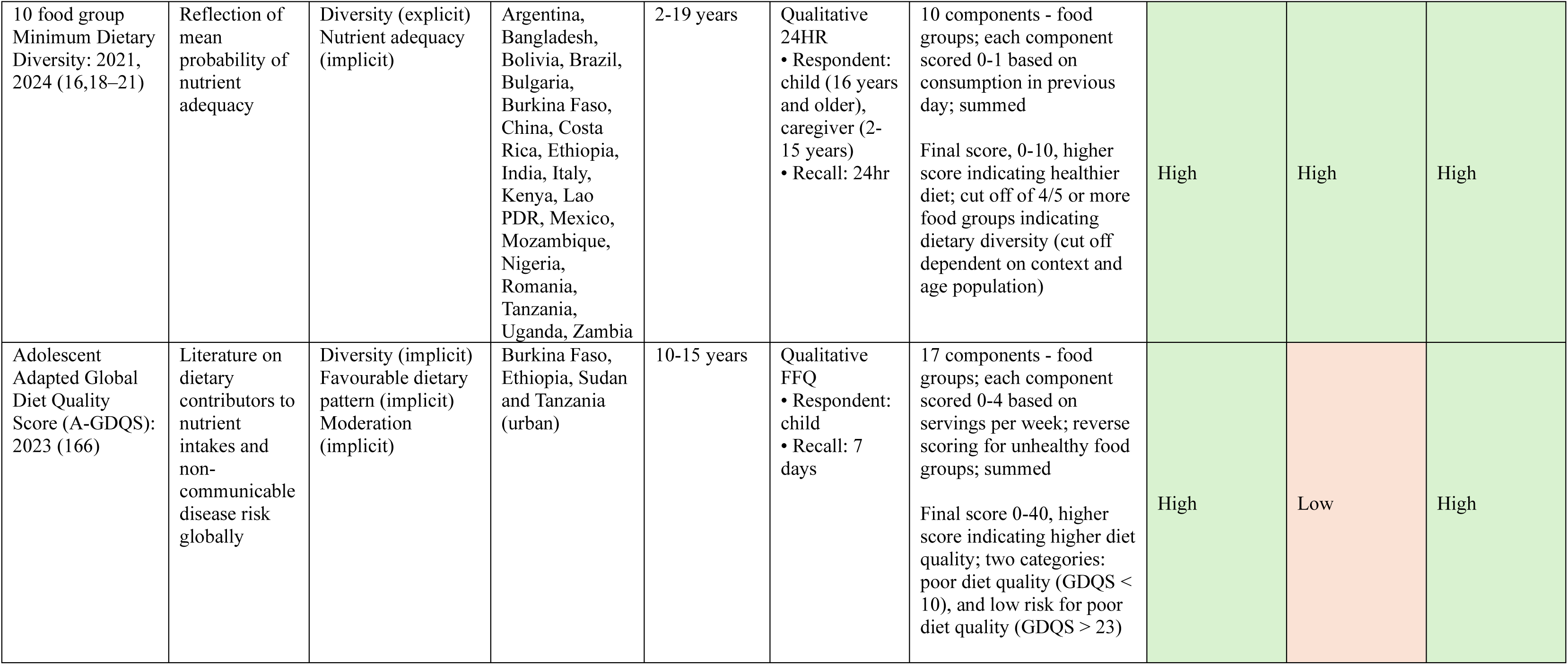
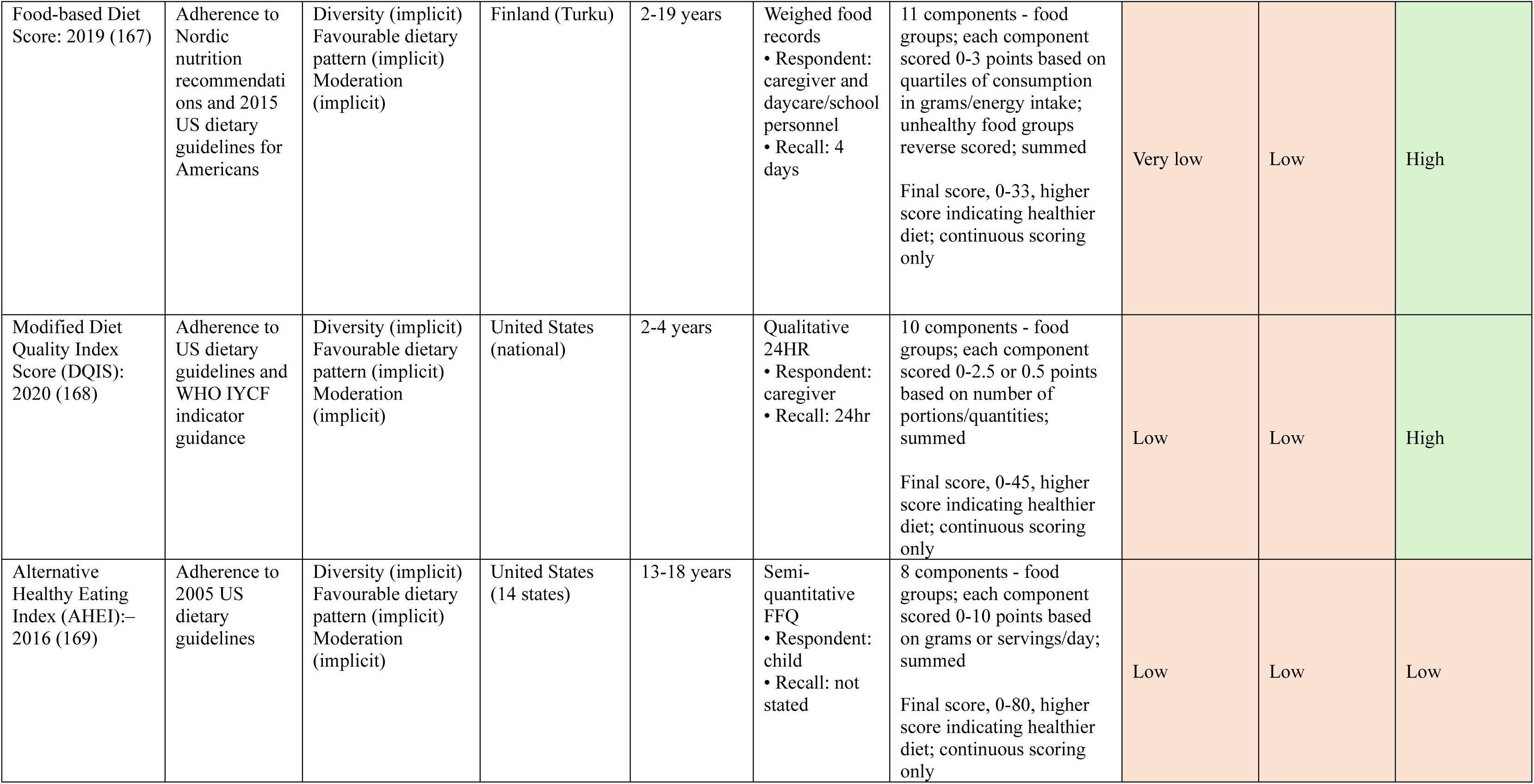
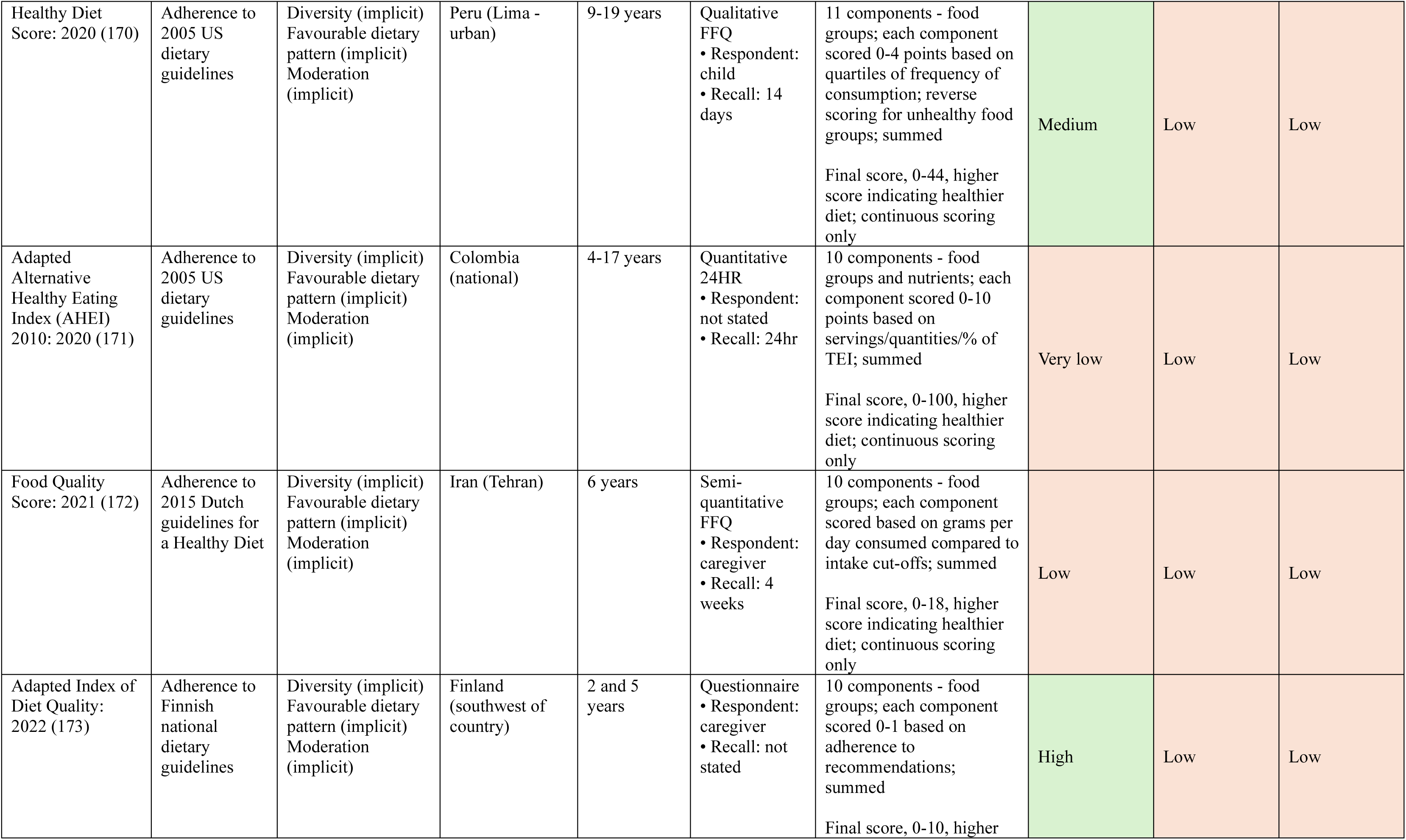
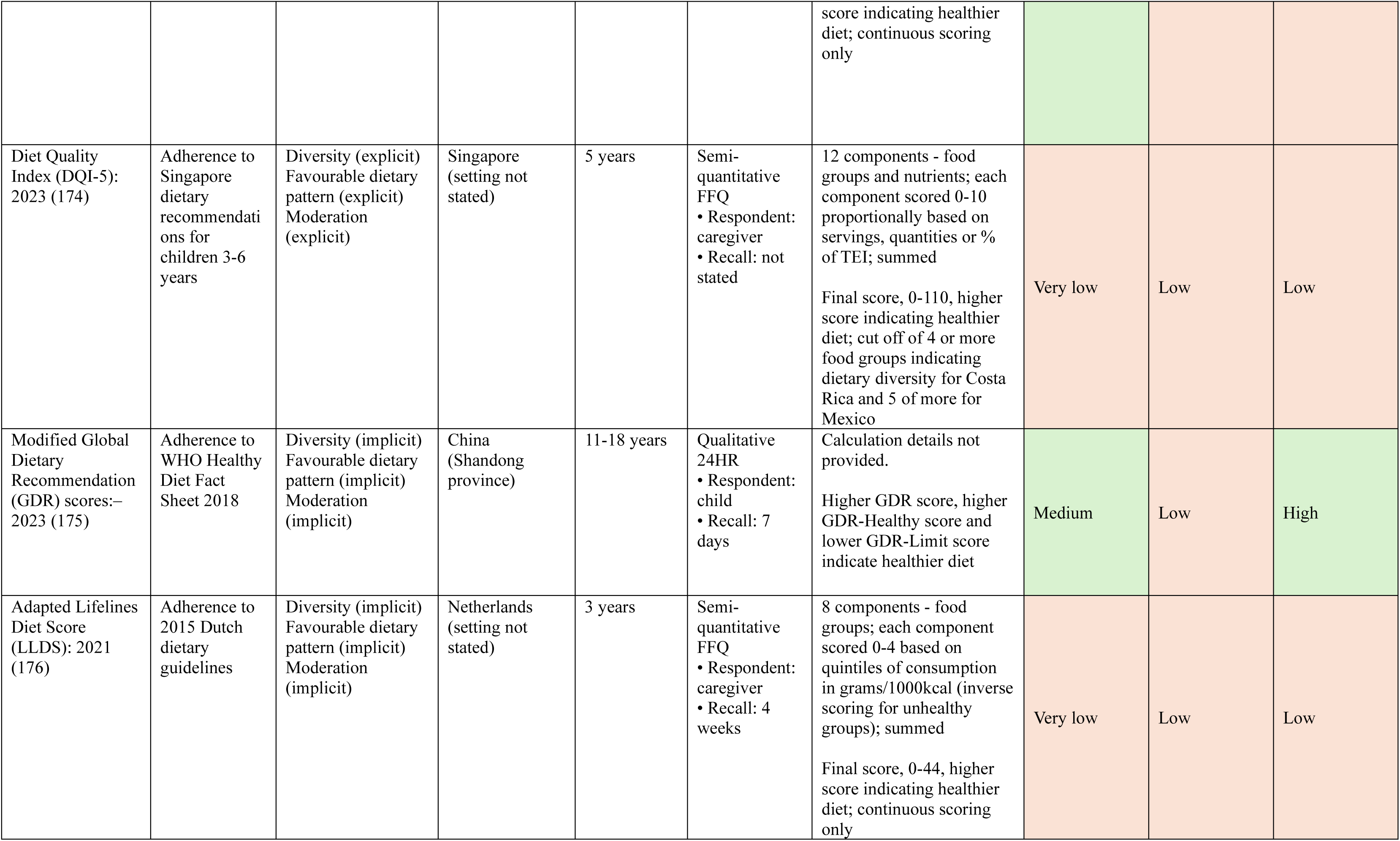
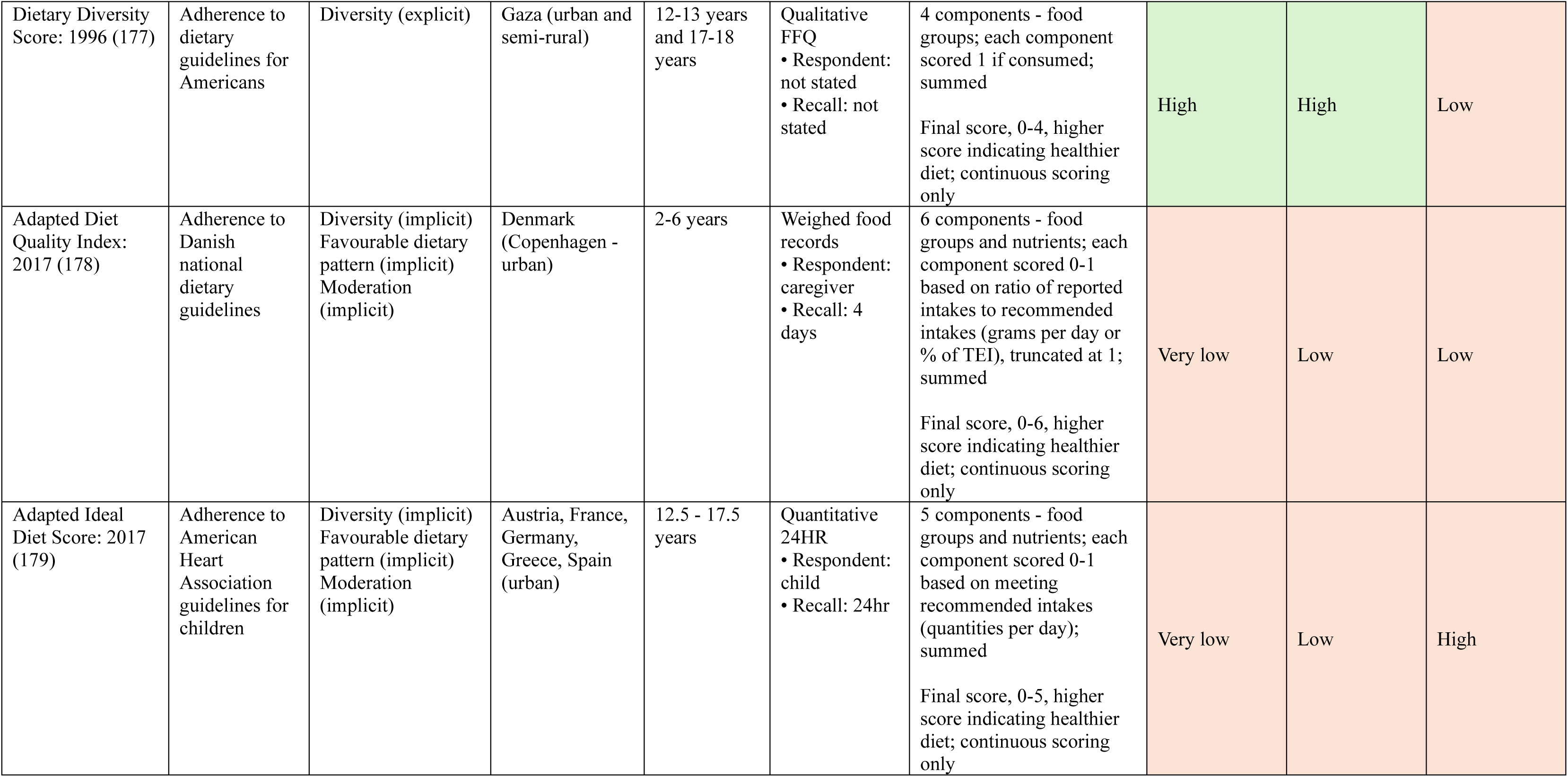
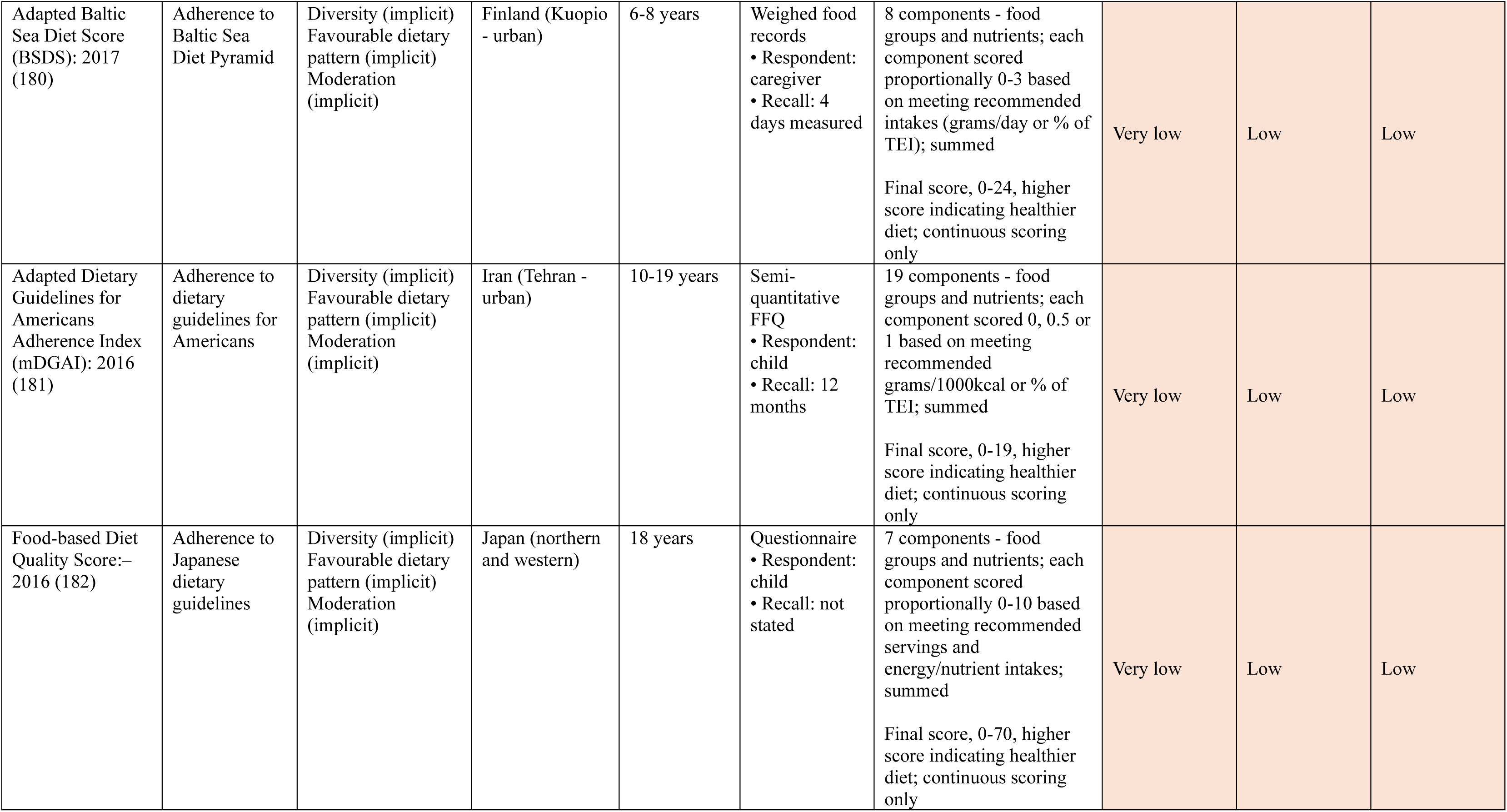
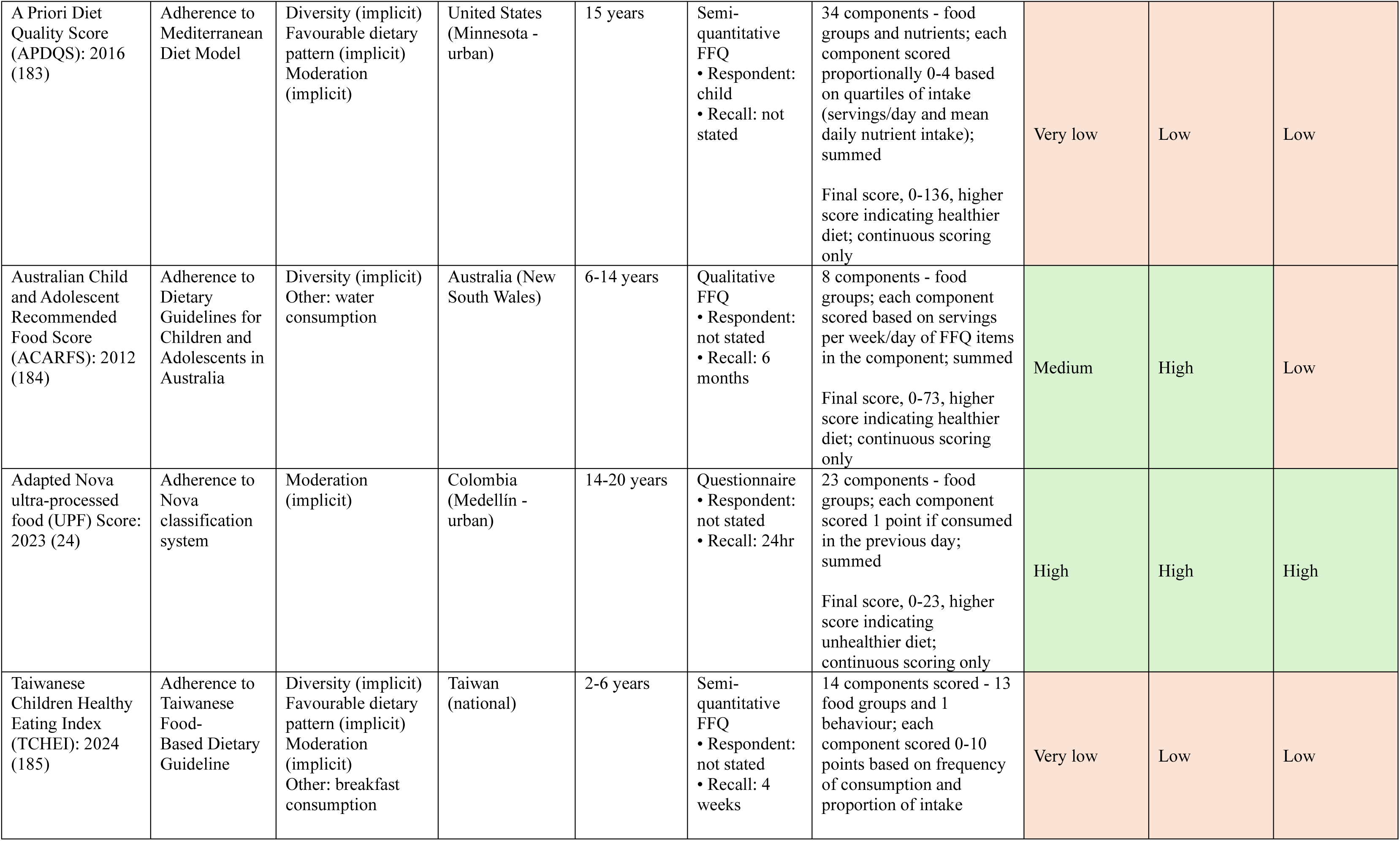
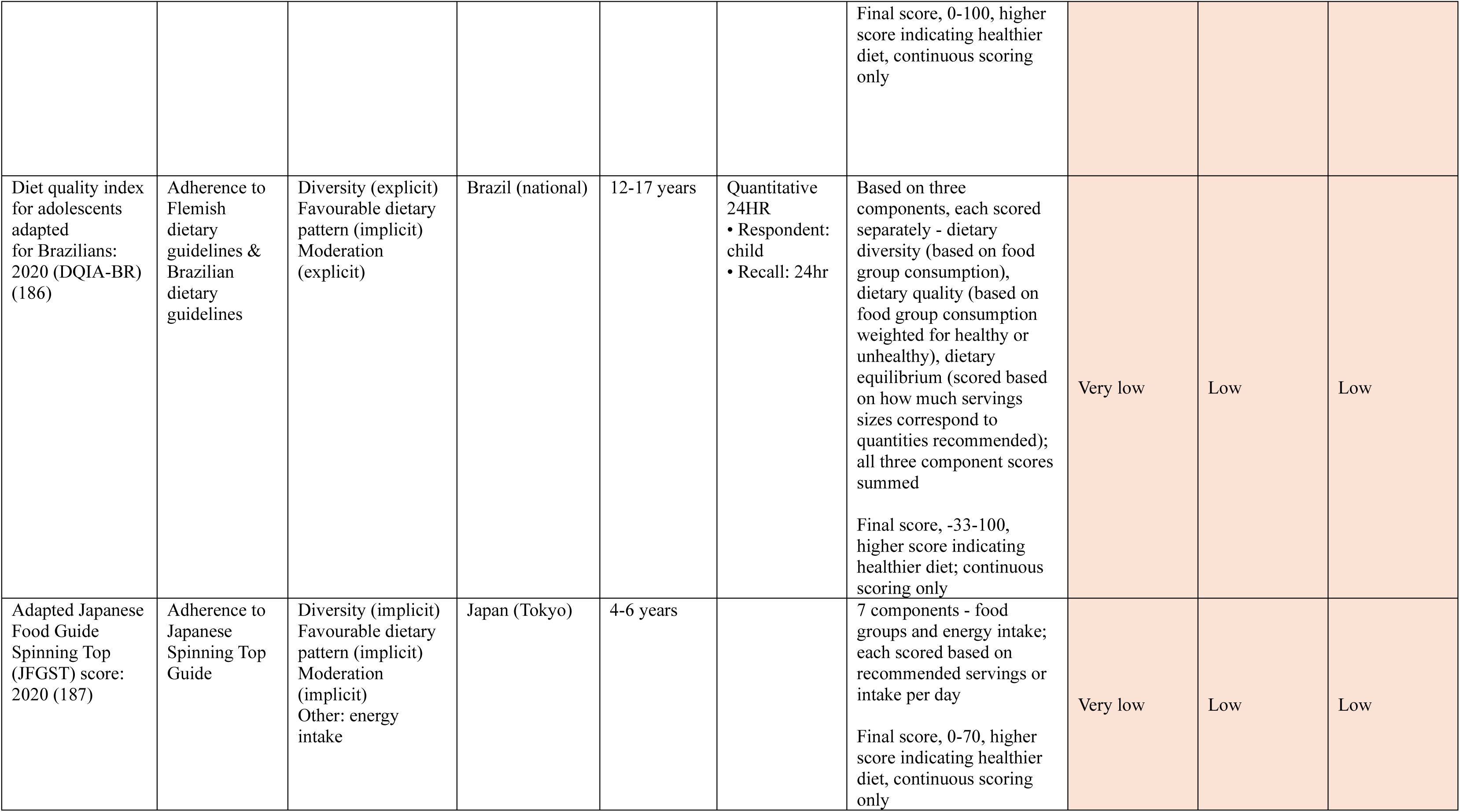
Identified developed or adapted healthy diet metrics for children and adolescents.

Over one-third of metrics (n=48, 37.8%) had been developed or adapted among pre-school age children (2-4 years), 55.9% (n=71) among primary school children (5-9 years), 66.9% (n=85) among younger adolescents (10-14 years), and 48.0% (n=61) among older adolescents (15-19 years). The 127 identified metrics were developed or adapted across 58 countries, including 30 nations classified as low-, lower-middle, or upper-middle –income by the World Bank (FY24), but most of these metrics were more frequently developed or adapted for high-income countries. Metrics were most commonly developed or adapted for the United States (n=18), Australia (n=11), Germany (n=10), and Greece (n=9).

### Dietary assessment methods

A range of dietary assessment methods were used to collect data for the identified metrics. The most common method was quantitative 24-hour dietary recalls with portion size estimation (n=46, 36.2%), followed by non-quantitative food frequency questionnaires (only frequency of consumption collected) (n=29, 22.8%) and semi-quantitative food frequency questionnaires (FFQ) (additionally including portion size estimation) (n=29, 22.8%). A variety of recall periods were noted across metrics utilizing food frequency questionnaires, ranging from 7 days to 12 months. Eleven metrics (8.7%) used metric-specific diet history questionnaires. Just over one-third of the metrics (n=46, 36.2%) relied on the child’s primary caregiver as the respondent, while 42.5% (n=54) used the child or adolescent as the respondent (with the remaining metrics not specifying the respondent). Of the 46 metrics where the primary caregiver served as the respondent, 78.3% (n=36) of these were for children aged 10 years or younger. Ten metrics involved the caregiver providing dietary intake information for younger children, with the child being the respondent once they reached a certain age. Across these ten metrics, the age cut-off for when caregivers versus children and adolescents served as the respondent ranged from 7 to 16 years, with a median of 10 years.

### Suitability of identified metrics for global monitoring

The feasibility of computation and interpretation varied among the identified healthy diet metrics for children and adolescents (**Table 4**). Overall, for ease of computation, only 35 (27.6%) and 22 metrics (17.3%) were rated as medium or high, respectively. More than half (n=70, 55.1%) required quantitative dietary intake data, such as the grams of foods or beverages consumed or energy/nutrient intakes. Most metrics (n=103, 81.1%) were rated low for ease of interpretation. This was primarily due to inclusion of both the sub-constructs of diversity and moderation within one composite metric, obscuring the ability to discern whether directionality of scoring was related to high diversity or low consumption of foods/nutrients to moderate. Adaptability of the metrics was generally low, with only 28 metrics (22.0%) rated as high, either being based on regional or global dietary guidance or having been developed or adapted for use in more than one country.

Of the 127 metrics, only five were identified as suitable for high-frequency global monitoring based on their feasibility and adaptability (**Table 5**). All five were determined to have low or medium interviewer and respondent burden according to discussions with child and adolescent diet assessment experts. These were i) the Individual Dietary Diversity Score (IDDS) (15), a metric measuring diversity of diets based on a simple count of consumption of 9 food groups across the previous day (scored as a discrete count); ii) 7 food group Minimum Dietary Diversity (MDD-7) (16), an adapted version of a WHO/UNICEF indicator for children 6-23 months of age (17) measuring diversity of diets across the previous day based on a 7-point food group diversity score (FGDS) (dichotomously scored with a cut-off of at least 4 food groups); iii) 10 food group Minimum Dietary Diversity (MDD-10) (16,18–21), an adapted version of a FAO indicator for women 15-49 years of age (22) measuring diversity of diets across the previous day based on a 10-point FGDS (dichotomously scored with a cut-off of at least 4 or 5 food groups based on the country and age sub-groups); iv) the Healthy Plate Variety Score (23), a metric measuring diversity of diets based on daily frequency of five food groups (continuously scored); and v) the Adapted Nova ultra-processed food (UPF) score (24), an adapted version of a 2021 indicator (25) measuring moderation of diets based on consumption of 23 UPF sub-groups in the previous day (continuously scored).

**Table 5.** Healthy diet metrics identified as suitable for global monitoring among children and adolescents.

| **Metric** | **Construction** | **Scoring** |
| --- | --- | --- |
| Individual Dietary Diversity Score (15) | 10 components - food groups; each component receiving 1 point if consumed in previous day; summed<br>Food groups:<br>1) cereals and tubers; 2) meat, poultry and fish; 3) dairy; 4) eggs; 5) pulses and nuts; 6) vitamin A-rich fruits and vegetables; 7) other fruit; 8) other vegetables; and 9) oils and fats. Excluded a 10<sup>th</sup> food group of 'other foods'. | Final score: 0-9, higher score indicating healthier diet; continuous scoring only |
| 7 food group Minimum Dietary Diversity (16) | 7 components - food groups; each component scored 0-1 based on consumption in previous day; summed<br>Food groups:<br>1) grains, roots, and tubers; 2) legumes and nuts; 3) dairy products; 4) flesh foods; 5) eggs; 6) vitamin A-rich fruits and vegetables; and 7) other fruits and vegetables | Final score: 0-7, higher score indicating healthier diet<br>Cut off of $\geq 4$ food groups indicating adequate dietary diversity |
| 10 food group Minimum Dietary Diversity (16,18–21) | 10 components - food groups; each component scored 0-1 based on consumption in previous day; summed<br>Food groups:<br>1) grains, white roots and tubers, and plantains; 2) pulses; 3) nuts and seeds; 4) dairy; 5) flesh foods; 6) eggs; 7) dark-green leafy vegetables; 8) vitamin A-rich fruits and vegetables; 9) other vegetables; and 10) other fruits | Final score: 0-10, higher score indicating healthier diet<br>Cut off of $\geq 4$ or $\geq 5$ food groups indicating adequate dietary diversity, depending on context |
| Healthy Plate Variety Score (23) | 5 components scored - food groups; each component scored based on ratio of recommended servings to consumed servings and truncated at 1; summed<br>Food groups:<br>1) starchy foods (including potatoes); 2) fruits; 3) vegetables; 4) meat, fish and alternatives; and 5) dairy foods | Final score; 0-5, higher score indicating healthier diet; continuous scoring only |
| Adapted Nova ultra-processed food (UPF) score (24) | 23 components – UPF sub-groups; each component scored 1 point if consumed in the previous day; summed<br>UPF sub-groups:<br>1) regular or noncaloric, light, or zero calorie sodas; 2) juice in a box or in a bottle; 3) powdered mixes to prepare soft drinks; 4) powdered mixes for chocolate drinks (powdered and ready-to-drink); 5) tea drinks (powdered and ready-to-drink); 6) fruit-flavored milk drinks; 7) sausage, hamburger meat, or chicken nuggets; 8) ham or mortadella; 9) sliced bread, hot dog bun, or hamburger bun; 10) table and cooking margarines; 11) frozen french fries or fast-food-chain fries; 12) ketchup, mayonnaise, or mustard sauce; 13) salad dressings or vinaigrettes (ready-to-eat) 14) instant noodles or packaged soups; 15) frozen or fast-food restaurant pizza; 16) | Final score: 0-23, higher score indicating unhealthier diet; continuous scoring only |
|  | lasagna or other frozen meals to heat or prepare at home; 17) salted packaged products (chips or crackers); 18) sweet biscuits with or without filling; 19) cake or packaged cake; 20) cereal bars; 21) ice cream or frozen popsicles; 22) chocolates and chocolate bonbons; 23) commercial breakfast cereal |  |

### Validity and reliability of metrics suitable for global monitoring

A total of 21 papers were identified with results among children and adolescents aged 2-19 years pertaining to the validity and reliability of the four of five metrics identified as suitable for global monitoring. Thirteen papers provided results for MDD-10 (eight with positive and five with neutral study quality), five papers pertained to the MDD-7 (one also covered MDD-10; three with positive and two with neutral study quality, three papers pertained to the IDDS developed by Kennedy et al. (15) (all with positive study quality), and one paper pertained to the Adapted Nova UPF score (neutral study quality) (**Table 6**). No papers containing validation or reliability results were identified for the Healthy Plate Variety Score. Validation and reliability evidence was primarily for construct validity, cross-context equivalence, and predictive capacity for health or nutrition outcomes; no results on test-retest reliability or sensitivity to change over time were identified.

**Table 6.**
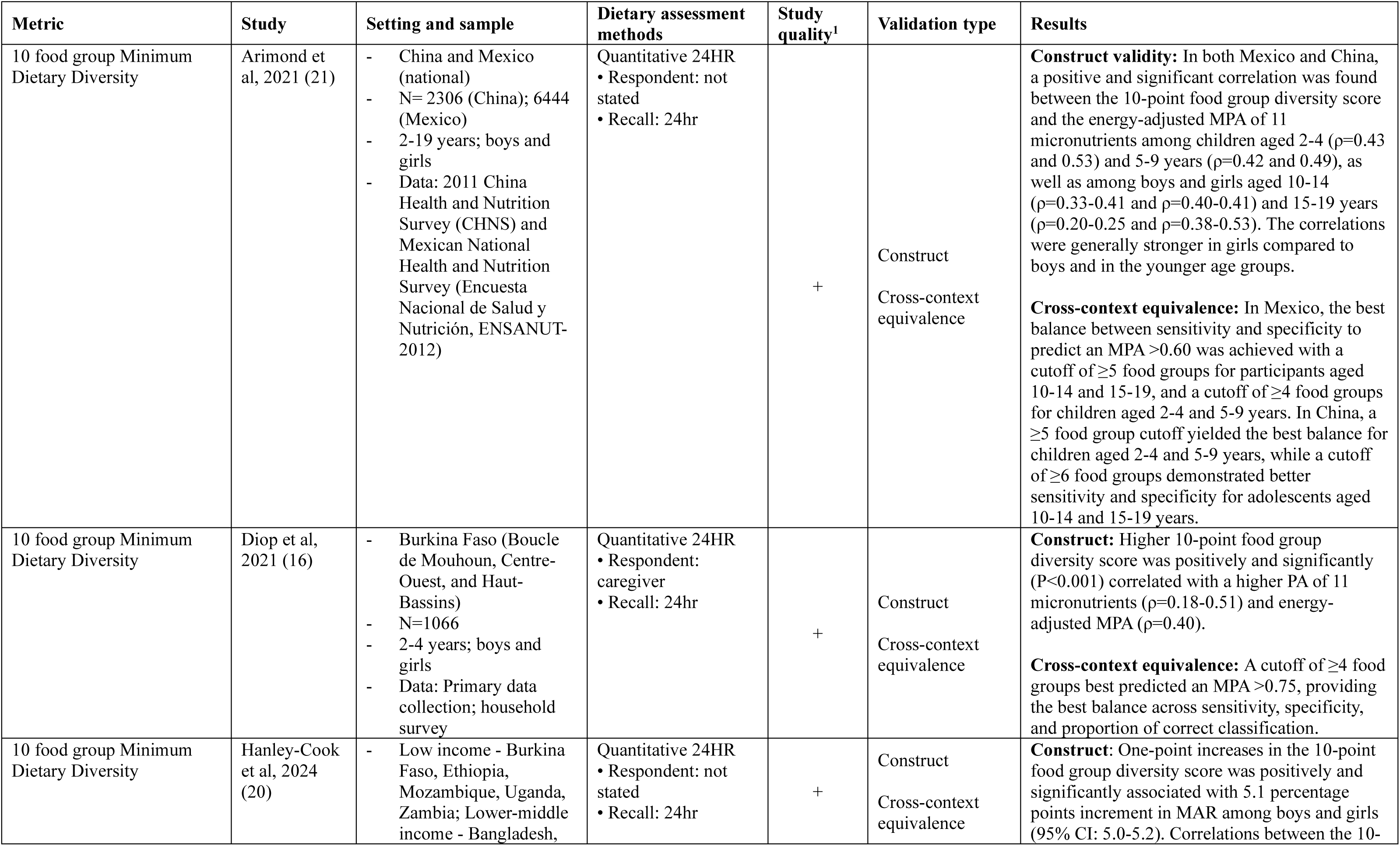
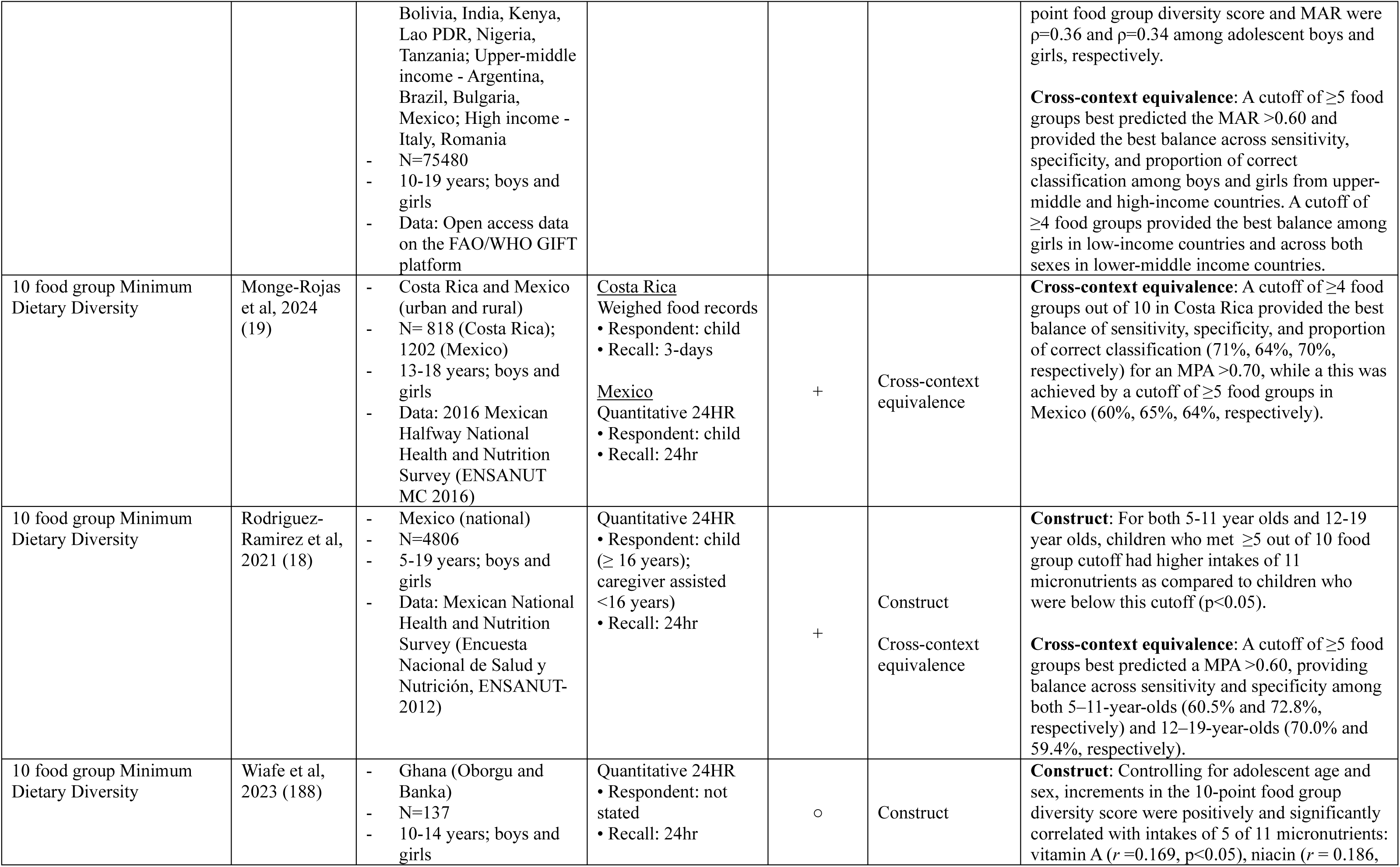
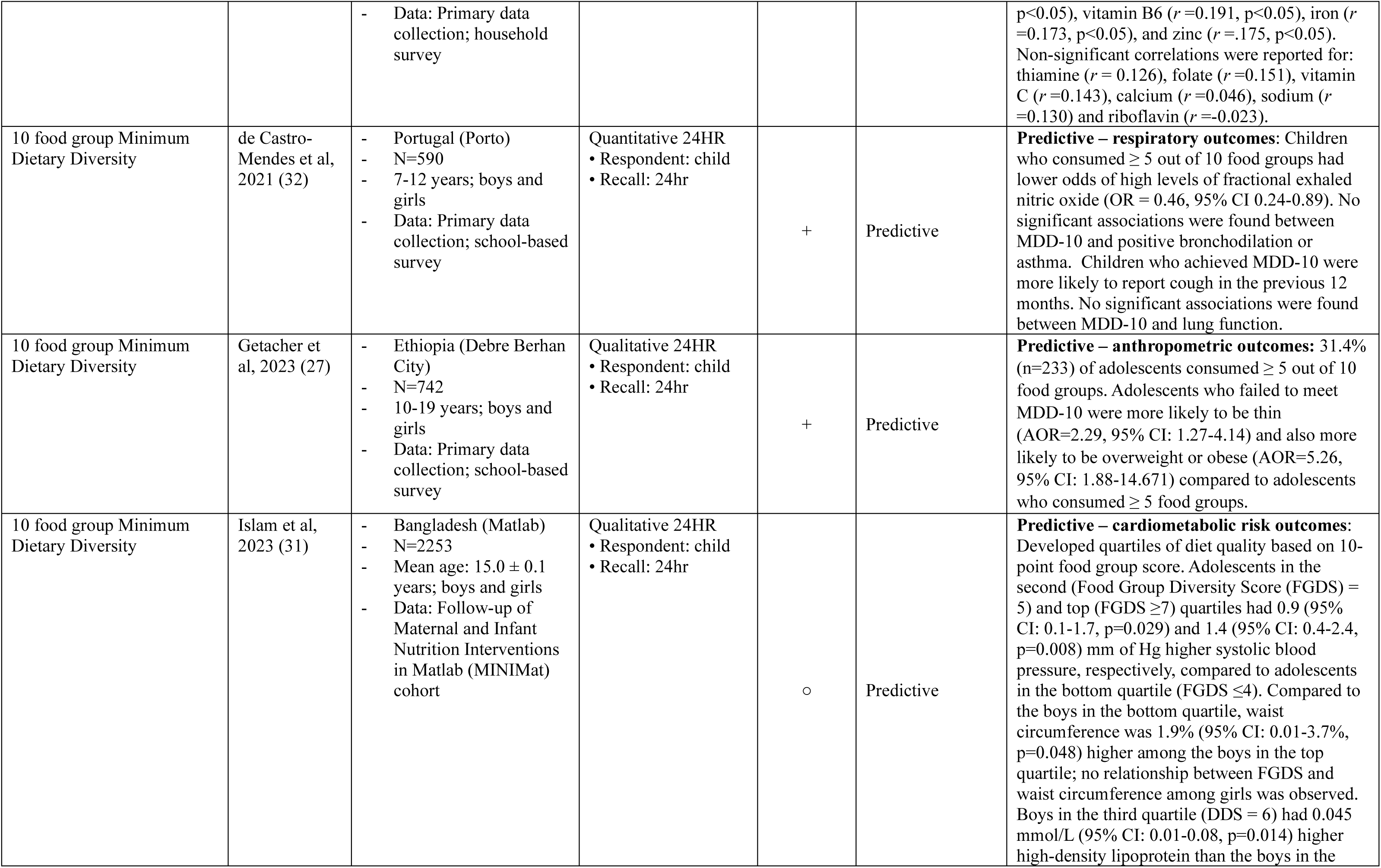
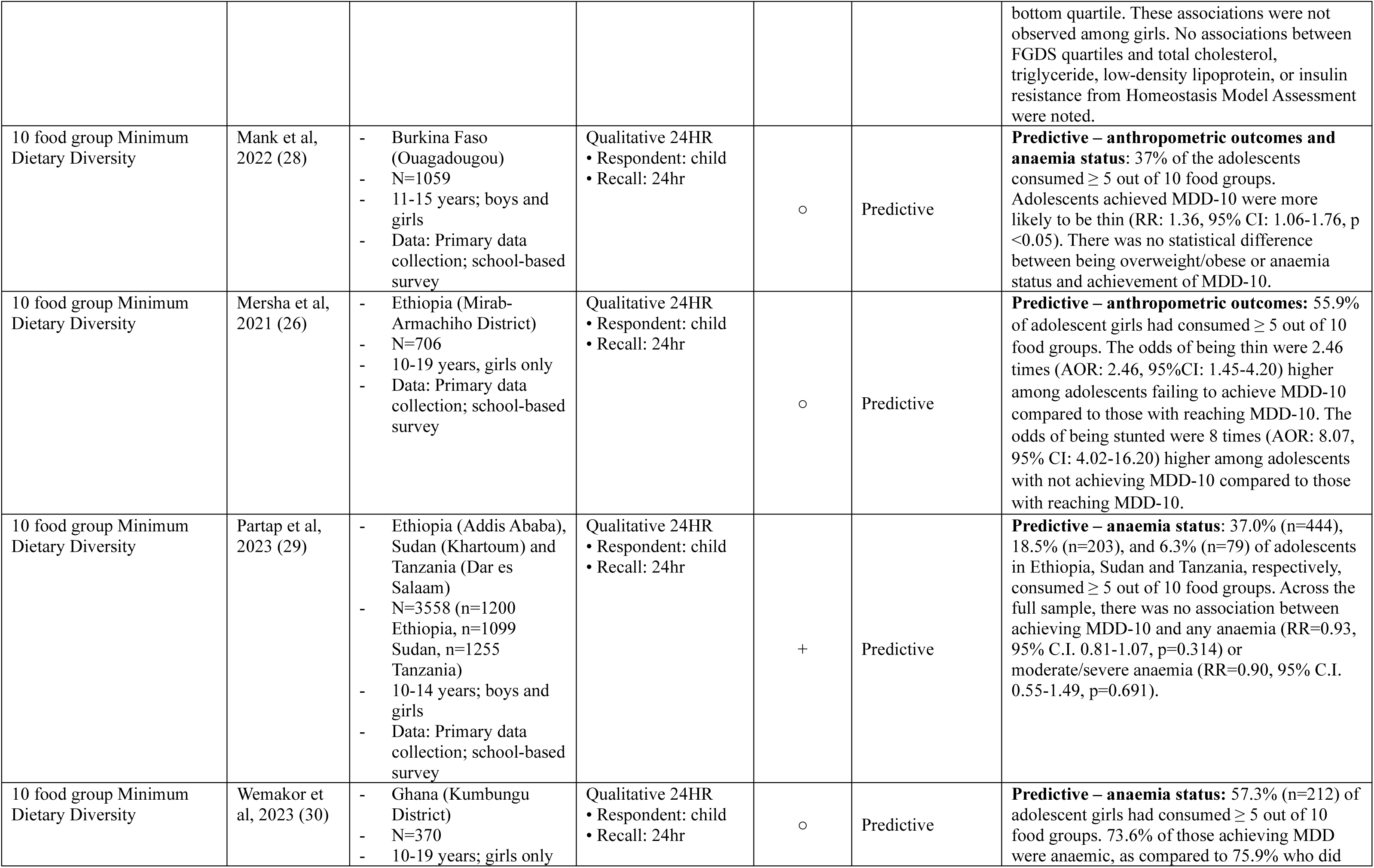
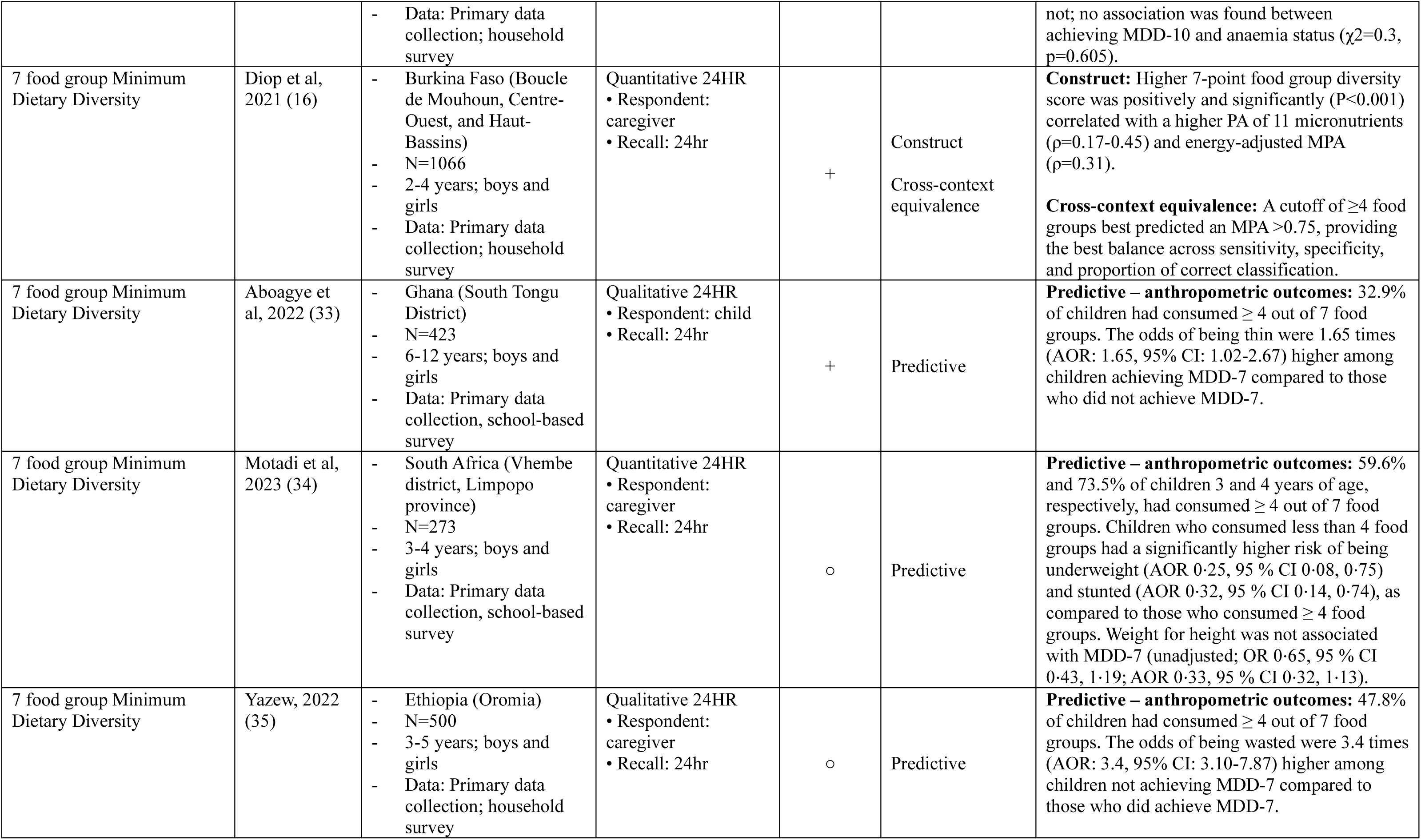
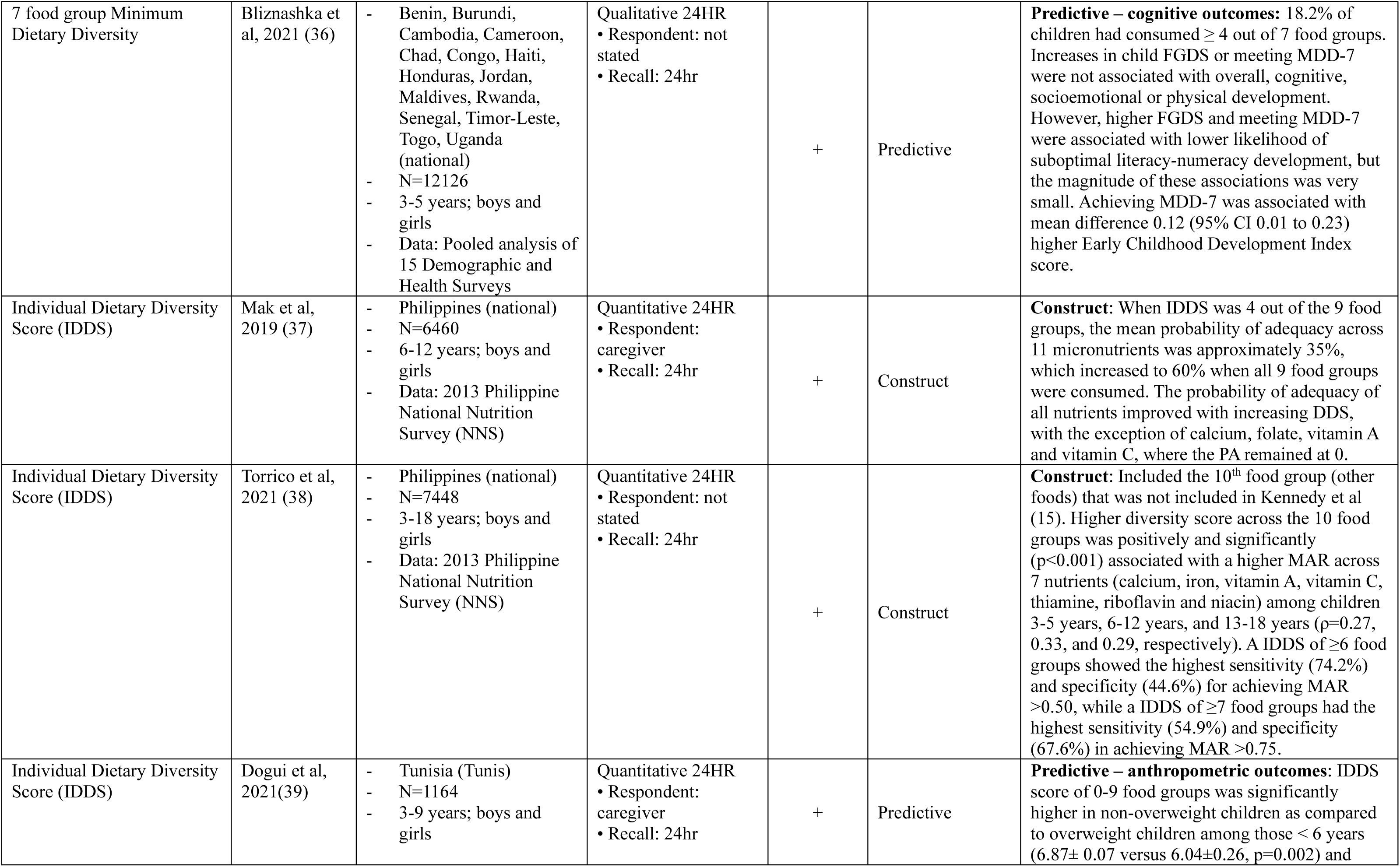
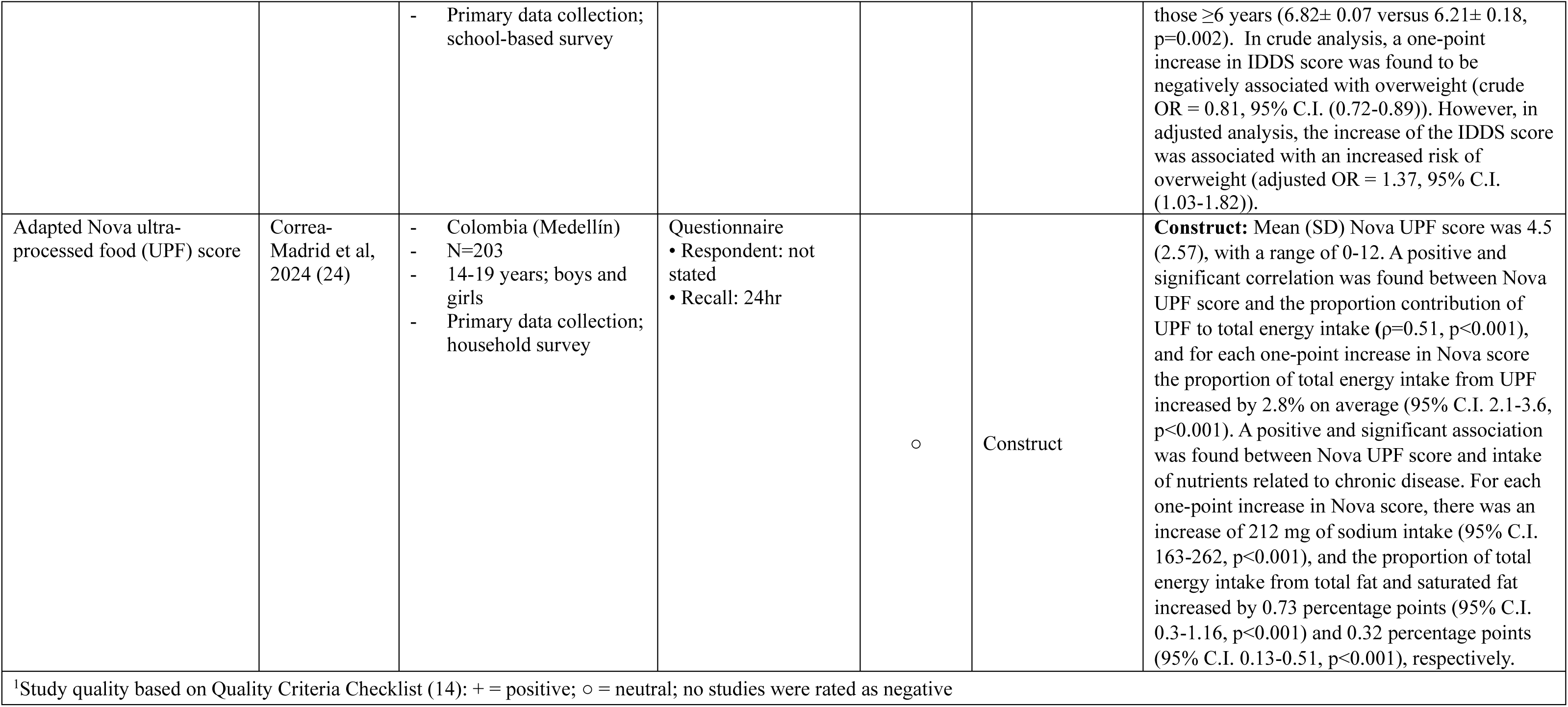
Validity of healthy diet metrics suitable for global monitoring among children and adolescents.

### 10 food groups Minimum Dietary Diversity (MDD-10)

The MDD-10 metric demonstrated relatively consistent construct validity across diverse settings, with positive associations reported between food group diversity and micronutrient adequacy among children and adolescents in China, Mexico, Burkina Faso, and across 18 countries globally (16,20,21). In Mexico and China, these associations were generally stronger among younger children and girls (21).

The cross-context equivalence of the food group cutoff for MDD-10 has been explored across a range of countries and age groups, with a cutoff of ≥4 or ≥5 food groups out of ten performing most consistently (16,20,21). (20)

The predictive capacity of the MDD-10 metric for nutrition/health outcomes varied across contexts. Not meeting the MDD-10 cutoff of ≥5 food groups was associated with undernutrition in some adolescent populations (26,27), while meeting this cutoff was associated with an increased risk of thinness in other adolescent populations (28). No associations were found between MDD-10 and anaemia status (28–30). One study noted an association between MDD-10 and cardiometabolic markers, with higher MDD-10 scores correlating with higher waist circumference, blood pressure and high-density lipoprotein (HDL) (31), while another found a positive association between achieving MDD-10 and lower odds of respiratory inflammation, (32).

### 7 food groups Minimum Dietary Diversity (MDD-7)

Evidence on the construct validity of MDD-7 is limited. One study from Burkina Faso found moderate correlations between MDD-7 and micronutrient adequacy among children aged 2–4 years, with a cutoff of ≥4 food groups offering the best balance between sensitivity and specificity (16). MDD-10 performed slightly better than MDD-7 in predicting nutrient adequacy in this setting.(16).

The predictive validity of the MDD-7 metric for nutrition and health outcomes varied across contexts and ages. Achieving a cutoff of ≥4 food groups was associated with both increased and decreased risk of undernutrition depending on the country and age group (33–35). (34)(35)One multi-country study found no overall associations with child development outcomes, though small positive associations were reported for early literacy and socioemotional development (36).

### Individual Dietary Diversity Score (IDDS)

Two studies assessing the construct validity of the IDDS developed by Kennedy et al. (15) reported positive associations with nutrient adequacy among children and adolescents in the Philippines(37,38). Associations were consistent across age groups, though some nutrients (e.g., calcium, folate, vitamin A, and vitamin C) showed weaker relationships in one study (37). (38).

Limited evidence exists on predictive capacity, with one study reporting a higher risk of overweight among 3–9-year-olds with higher diversity scores in urban Tunisia a (39). No studies assessing the cross-context equivalence of IDDS were identified.

### Adapted Nova UPF score

The construct validity of the Adapted Nova UPF score was supported by one study among Colombian adolescents, which found that higher scores were associated with increased energy intake from ultra-processed foods and elevated intakes of sodium, total fat, and saturated fat

(24). No studies were identified assessing the cross-context equivalence or predictive capacity of the Adapted Nova UPF score for nutrition or health outcomes.

## Discussion

Despite the global urgency to improve the diets of children and adolescents, data on the healthiness of their diets remains scarce (40,41). This gap is partly due to the lack of universally recognized low-burden metrics for monitoring healthy diets within this age group. Previous systematic reviews, including Marshall et al. (2014) (7) and Dalwood et al. (2020) (8), have catalogued existing diet quality indices used for children and adolescents, identifying a large and growing number of metrics. These reviews, however, did not evaluate which metrics are suitable for global monitoring purposes - a critical need given growing international efforts to track progress on nutrition and health targets. Findings from this review build on this prior work by identifying 127 distinct healthy diet metrics and critically assessing their feasibility, adaptability, and evidence base for global monitoring among children and adolescents 2-19 years of age.

Only five healthy diet metrics designed to capture healthiness of child and adolescent diets demonstrated sufficient feasibility and adaptability to be considered suitable for global monitoring. This reinforces the concern that, although many metrics exist, few are practical for wide-scale application across diverse contexts. Most remaining metrics relied on quantitative dietary intake data that require substantial time and resources for data collection and processing, measure multiple sub-constructs of healthy diets that obscures clear understanding of dietary challenges across populations (42), or were developed based on national dietary guidelines, limiting cross-country comparability. The strength of evidence supporting construct validity and cross-context equivalence varied across the five suitable metrics, with some metrics more extensively studied than others. Prior research has also underscored limited evidence on cross-context equivalence of healthy diet metrics in adult populations (42). Other gaps exist for all metrics, specifically regarding their sensitivity to change and test-retest reliability, highlighting the need for further research to support their use for global monitoring.

Of the five metrics identified, MDD-10 was found to be a low-burden metric with high ease of computation and interpretation, and has the greatest body of evidence supporting its construct validity and cross-context equivalence in over 20 countries across Africa, Asia, Europe and Latin America. A cutoff of ≥5 food groups provided optimal sensitivity and specificity in higher-income settings, while a cutoff of ≥4 food groups was more appropriate in lower-income contexts, effectively predicting mean adequacy ratio or the mean probability of adequacy of essential nutrients across different populations (16,20,21). More recent validation of MDD-10 among 4–15-year-old children and adolescents across seven low- and middle-income countries found that the optimal cutoff for predicting micronutrient adequacy varied between 4 to 6 food groups by country, but a cutoff of ≥5 demonstrated the highest performance in pooled analyses among this age group (43). This variation underscores that no single cutoff is universally optimal—an insight consistent with previous findings from MDD-W validation studies among women, where the most appropriate cutoff ranged from 4 to 6 depending on the population/dietary pattern(21,44). Despite this, a global cutoff of ≥5 has been proposed to support standardized monitoring and cross-country comparison (20). Both the construct validity and cross-context equivalence evidence for MDD-10 was predominantly assessed among children and adolescents 5-19 years of age, and further research among pre-school children aged 2-4 years is needed.

A limitation of the five metrics identified in this review is their narrow focus on dietary diversity. The predominant focus of healthy diet metrics on dietary diversity, which can serve as a proxy for nutrient adequacy, has been noted in other reviews of diet indices for populations of all ages (45). While diversity is a critical aspect of a healthy diet, facilitating the nutrient adequacy needed for growing children and adolescents, the limited attention to other important sub-constructs, particularly moderation, is concerning. Moderation is particularly relevant for this population given the rising consumption of unhealthy foods and growing global burden of childhood obesity (46). Among the four metrics, only the Adapted Nova UPF score measured moderation, specifically targeting the intake of UPF. Although UPF are an important health concern, particularly due to their association with obesity (47,48), metabolic disorders (49), and inadequate nutrient intakes (50), this metric does not fully capture the broader spectrum of unhealthy foods that are prevalent in children and adolescents’ diets. Foods high in added sugars, unhealthy fats, and sodium, which may not be classified as ultra-processed, still pose substantial risks to children and adolescents’ health and nutritional status (51). Additionally, some evidence suggests that not all UPF have the same impact on health outcomes, with certain foods—such as processed meats and sugar-sweetened beverages—posing a higher health risk than other UPF (52,53). It is important to note that the composition of UPF sub-groups can vary by context when the Nova UPF score metric is adapted (54), posing challenges for cross-country comparisons and complicating global monitoring efforts. The lack of comprehensive metrics to assess moderation highlights the need for the development of more holistic methods that can provide a fuller picture of children and adolescents’ dietary risks and help guide more effective policies and interventions.

An additional gap for these five identified metrics is the limited validation evidence pertinent to global monitoring, specifically in terms of test-retest reliability and sensitivity to change. These aspects are crucial for ensuring that metrics can reliably measure healthy diet sub-constructs over time and accurately reflect changes in these sub-constructs (12), whether due to interventions or natural variations in eating patterns across seasons, time and contexts. Test-retest reliability means a metric will consistently produce the same results under similar conditions, reinforcing the confidence in their use for global monitoring. Sensitivity to change is equally important, as it determines a metric’s ability to detect meaningful changes in dietary intake, which is critical for evaluating the impact of nutrition actions. The absence of this robust evidence for the four child- and adolescent-focused metrics identified in this review raises concerns about their overall measurement accuracy and reliability. The reliability and sensitivity of the MDD-W, the basis for MDD-10, has been more rigorously evaluated among women of reproductive age, with several studies showing MDD-W to accurately reflect when changes in diet diversity did or did not occur in populations, in alignment with changes in nutrient intakes, across seasons or in response to interventions (55–58). The lack of similar evidence for child-focused diet metrics underscores a critical need for further research to validate these methods and metrics, particularly in terms of their consistency and responsiveness to dietary changes. Without this validation, the full utility of these metrics for monitoring healthiness of diets among this age group remains uncertain.

Furthermore, the studies of the five metrics only investigated relationships among individuals within populations whereas the primary interests for global monitoring are differences among, and change over time in, populations and sub-populations. Explicit evidence is needed to determine whether metrics feasible for global monitoring can adequately differentiate healthy diets of children and adolescents across countries and how child and adolescent diets within countries change over time.

Of the five metrics identified as suitable for global monitoring, two stem from prior existing metrics for adult populations: MDD-10 and the Adapted Nova UPF score. The Global Diet Quality Score (GDQS) is also a healthy diet metric for adults currently being adapted and validated for use among children and younger adolescents (personal communication), which may be another promising option for global monitoring. Studies on the construct validity of both MDD-10 and the Adapted Nova UPF score suggest that metrics designed for assessing adult diets can be effectively adapted for use among children and adolescents, and this adaptation offers potential logistical and feasibility benefits for global monitoring efforts. For both MDD-10 and the Adapted Nova UPF score, the same food groups are included in the metrics for both adults and younger populations, facilitating measurement across different age groups within single platforms of data collection. Furthermore, the questionnaire used for collecting MDD-10 data is the same as that used for MDD-W and is already integrated into widely implemented national population-based survey platforms, such as the Demographic and Health Surveys (DHS) and Gallup World Poll (including older adolescents aged 15-19 years), as well as various national nutrition and health surveillance systems. These platforms would need to include younger age groups in their sampling, and potentially increase their sample sizes, to generate measurements of dietary diversity for children and adolescents. While MDD-10 shows promise as an effective adaptation for assessing dietary diversity among children and adolescents, future research should focus on adapting metrics that capture other sub-constructs of a healthy diet, such as moderation, to better support comprehensive monitoring.

While this review primarily focused on identifying metrics for assessing healthy diets among children and adolescents, the methods used to measure these metrics are equally critical when considering their suitability for global monitoring, as the choice of method can significantly impact the accuracy and reliability of the data collected. One of the key methodological questions for dietary assessment among children and adolescents is the accuracy and reliability of child-reported versus guardian-reported consumption information. Determining the appropriate age at which children and adolescents can reliably report their own dietary intake versus when a parent or guardian should be the respondent is crucial (59). This present review highlighted variations in age cutoffs among studies; studies among younger children and adolescents typically relied on parental reporting, the age at which children and adolescents became the primary respondent varied from as young as 7 years (60) to as old as 16 years (18). Parental recall may also be subject to bias and inaccuracies, particularly as parents may not always be present during all meals or eating occasions outside the home (61) or may have difficulty estimating portion sizes consumed by children and adolescents.

Recent advancements in dietary assessment methods may offer solutions to enhance the accuracy of dietary data collection in children and adolescents. Technology-based methods, including smartphone applications, provide innovative ways to assess dietary intake using visual aids and web-based platforms that can ease dietary data collection (62,63). Several of these methods have been shown to improve portion size estimation and reduce participant burden (64,65), making them potentially valuable for large-scale monitoring efforts among children and adolescents. More recently, the integration of artificial intelligence and machine learning into dietary assessment tools has opened up new possibilities for collecting dietary data with greater precision and regularity. These developments hold promise for increasing the feasibility of collecting more granular, quantitative dietary data at scale, an endeavour historically constrained by resource intensity and respondent fatigue. While such technologies are advancing rapidly, their validation in diverse child and adolescent populations is still needed to confirm appropriateness for global use. The effectiveness of these technological methods, alongside traditional methods like 24-hour recalls and food frequency questionnaires, must be carefully considered when selecting metrics and methods for assessing children and adolescents’ diets on a global scale.

Among the strengths of this review is the systematic method employed in the literature search, which ensured a comprehensive identification of existing metrics and methods for assessing healthy diets among children and adolescents. Additionally, prior to title and abstract screening, two reviewers independently evaluated a 5% sample of papers to ensure consistency in their assessments. This process helped identify and resolve discrepancies in title and abstract screening decisions early, leading to a more consistent and reliable review process. Additionally, evidence captured through this search was augmented by interviews with key experts in the field, which not only facilitated the sourcing of additional grey literature but also provided valuable insights into the logistical feasibility of data collection for the four identified metrics. These expert interviews added a practical dimension to the review and aided the assessment of real-world feasibility of the identified metrics, which is often not presented in published literature. Nonetheless, this review has several limitations related to its methodology. One significant limitation is the exclusion of non-English literature, which may have resulted in the omission of relevant studies published in other languages, potentially biasing the findings towards English-speaking regions and authors. In addition, while the review followed a systematic and structured method to searching and evaluating the literature, the review was not registered in a protocol repository (e.g. PROSPERO), and duplicate screening was not possible for all papers. Despite these limitations, this review provides a valuable synthesis of current knowledge and identifies key gaps for future research.

Regardless of the increasing number of metrics available to assess the healthiness of diets among children and adolescents, few demonstrate the feasibility and adaptability required for global monitoring. Furthermore, those metrics that offer a lightweight, feasible, universal indicator primarily focus on dietary diversity, and metrics development and validation for other critical sub-constructs of a healthy diet have lagged, such as the moderation of foods and nutrients associated with increased risk of non-communicable diseases, a growing concern worldwide. There are also critical gaps in understanding the reliability and sensitivity of these metrics, and explicit investigation of validity to differentiate populations and change in populations, which are essential for ensuring that these indicators can reliably track healthiness of diets over time, accurately reflect the impact of interventions, and compare populations to one another.

It is imperative for research to address these gaps, with a focus on developing or adapting additional metrics to encompass a broader range of sub-constructs of a healthy diet. Additionally, the exploration and validation of innovative, technology-based dietary assessment methods tailored for children and adolescents should be prioritized to enhance data accuracy and feasibility in global monitoring efforts. By strengthening the evidence base and expanding the scope of these methods, we can better support global initiatives aimed at providing guidance on monitoring to countries, improving child and adolescent diets, promoting healthier eating habits in the future, and enabling long-term well-being worldwide.

## Acknowledgements

The members of the Healthy Diets Monitoring Initiative are Víctor Aguayo (UNICEF), Francesco Branca (WHO), Elaine Borghi (WHO), Jennifer Coates (Tufts University Friedman School of Nutrition Science and Policy), Edward Frongillo (University of South Carolina), Giles Hanley-Cook (FAO), Karoline Hassfurter (UNICEF), Chika Hayashi (UNICEF), Bridget Holmes (FAO), Luc Ingenbleek (WHO), Pragya Mathema (UNICEF), Vrinda Mehra (UNICEF), Lynnette Neufeld (FAO), Kuntal Saha (WHO), and Isabela Fleury Sattamini (WHO). AMP, EAF, and JCC designed the research with input from all co-authors; AMP and HC conducted research; AMP analyzed data; and AMP and HC wrote the paper. AMP had primary responsibility for final content. All authors read and approved the final manuscript.

## Data availability

Data described in the manuscript will be made available upon reasonable request to the corresponding author.

## Abbreviations

DHS: Demographic and Health Surveys
FAO: Food and Agriculture Organization of the United Nations
FGDS: food group diversity score
FFQ: food frequency questionnaires
HDMI: Healthy Diets Monitoring Initiative
IDDS: individual dietary diversity score
MAR: mean adequacy ratio
MDD: minimum dietary diversity
MPA: mean probability of adequacy
QCC: Quality Criteria Checklist
UNICEF: United Nations Children’s Fund
UPF: ultra-processed foods
WHO: World Health Organization

