## Supplemental Table 1 for "Healthy diet metrics for children and adolescents and their suitability for global monitoring: a critical review"

| **Supplemental Table 1.** Definitions of subconstructs of a healthy diet | | |
| --- | --- | --- |
| **Sub-construct** | **Definition** | **Sources** |
| Nutrient adequacy | “Sufficient quantity and quality of nutrient intake compared with nutrient requirements to meet dietary needs, without excess”   - Ex. Nutrient intake meets reference value | Verger et al 2023 (1) |
| Nutrient density | “Amount or relative proportion of nutrients per weight of food, per unit of energy (often kcal), or per serving”   - Ex. Quantity of nutrients consumed per quantity of food/energy intake | Verger et al 2023 (1) |
| Macronutrient balance | “Balance of energy-yielding macronutrients: carbohydrates, proteins and fats”   - Ex. Proportionality within range of reference value | Verger et al 2023 (1) |
| Diversity | “Diets composed of a variety of foods derived from diverse food groups”   - Ex. Variety of foods within and across food groups | Verger et al 2023 |
| Moderation | “Limited intake of foods related to chronic diseases, including refined grains, red and processed meats, and sugar-sweetened foods and beverages”   - Ex. Limiting intake of unhealthy dietary components or total energy intake | Arimond & Deitchler 2019(2) |
| Favourable dietary pattern | Metric evaluates both healthy and unhealthy food consumption; composite of diversity and moderation sub-constructs. "Common characteristics of dietary patterns associated with positive health outcomes include relatively higher intake of vegetables, fruits, legumes, whole grains, dairy, lean meats and poultry, seafood, nuts, and unsaturated vegetables oils, and relatively lower consumption of red and processed meats, sugar-sweetened foods and beverages, and refined grains” | USDA & USHHS 2020 (3); Verger et al 2023 (1) |
| Food safety | “Foods are free of microbial pathogens, foodborne macroparasites, toxins and chemicals”   - Metric evaluates contamination - Distinct from food hygiene behaviors (ex. handwashing; food storage practices) | Verger et al 2023 (1) |
